# Comprehensive genetic analysis of the human lipidome identifies novel loci controlling lipid homeostasis with links to coronary artery disease

**DOI:** 10.1101/2021.08.20.21261814

**Authors:** Gemma Cadby, Corey Giles, Phillip E Melton, Kevin Huynh, Natalie A Mellett, Thy Duong, Anh Nguyen, Michelle Cinel, Alex Smith, Gavriel Olshansky, Tingting Wang, Marta Brozynska, Mike Inouye, Nina S McCarthy, Amir Ariff, Joseph Hung, Jennie Hui, John Beilby, Marie-Pierre Dubé, Gerald F Watts, Sonia Shah, Naomi R Wray, Wei Ling Florence Lim, Pratishtha Chatterjee, Ian Martins, Simon M Laws, Tenielle Porter, Michael Vacher, Ashley I Bush, Christopher C Rowe, Victor L Villemagne, David Ames, Colin L Masters, Kevin Taddei, Matthias Arnold, Gabi Kastenmüller, Kwangsik Nho, Andrew J Saykin, Xianlin Han, Rima Kaddurah-Daouk, Ralph N Martins, John Blangero, Peter J Meikle, Eric K Moses

## Abstract

We integrated lipidomics and genomics to unravel the genetic architecture of lipid metabolism and identify genetic variants associated with lipid species that are putatively in the mechanistic pathway to coronary artery disease (CAD). We quantified 596 lipid species in serum from 4,492 phenotyped individuals from the Busselton Health Study. In our discovery GWAS we identified 667 independent loci associations with these lipid species (479 novel), followed by meta-analysis and validation in two independent cohorts. Lipid endophenotypes (134) identified for CAD were associated with variation at 186 genomic loci. Associations between independent lipid-loci with coronary atherosclerosis were assessed in ∼456,000 individuals from the UK Biobank. Of the 53 lipid-loci that showed evidence of association (P<1×10^−3^), 43 loci were associated with at least one of the 134 lipid endophenotypes. The findings of this study illustrate the value of integrative biology to investigate the genetics and lipid metabolism in the aetiology of atherosclerosis and CAD, with implications for other complex diseases.

## Introduction

Lipids comprise thousands of individual species, spanning many classes and subclasses. Genome-wide association studies (GWAS) of lipid species can provide novel insights into human physiology, inborn errors of metabolism and mechanisms for complex traits and diseases. Dyslipidaemia, a broad term for disordered lipid and lipoprotein, is a major risk factor for atherosclerotic cardiovascular disease and a therapeutic target for the primary and secondary prevention of coronary artery disease (CAD)^1,2^. Defined by elevated low-density lipoprotein cholesterol (LDL-C) and triglycerides with decreased high-density lipoprotein cholesterol (HDL-C) – these ‘clinical lipid’ measures provide only a partial view of the complex lipoprotein structures and their metabolism. Lipidomic technologies can now measure hundreds of individual molecular lipid species that make up the human lipidome, providing a more complete snapshot of the underlying lipid metabolism occurring within an individual.

Genome-wide association studies have uncovered thousands of genetic variants linked to traditional clinical lipids (LDL-cholesterol, HDL-cholesterol, triglycerides)^3,4^. Genes implicated at these loci show functional links between lipid levels and CAD^5^. The human lipidome is heritable and predictive of CAD, furthering our understanding of the biology of CAD^6^. The individual lipid species that make up the lipidome are biologically simpler measures that may reside closer to the causal action of genes, making them valuable endophenotypes for gene identification. Genetic interrogation of the human lipidome may therefore reveal further genetic variants that play a role in lipid metabolism and CAD.

Compared with other complex traits, relatively few genomic loci have been associated with lipid species in GWAS of the human serum/plasma lipidome^7-17^, although these studies have generally interrogated a restricted subset of lipid species. The serum lipidome is complex and consists of many isobaric and isomeric species that share elemental composition but are structurally distinct. Existing lipidomic studies often employ techniques that provide poor resolution of these species, limiting their biological interpretation. We have recently expanded our lipidomic platform to better characterise isomeric lipid species, now measuring 596 lipids from 33 classes^18^. Our methodology focuses on the precise measurement of a broad number of lipid and lipid-like compounds, utilising extensive chromatographic separation.

Here, we report a GWAS of 596 targeted lipid species (across 33 lipid classes) in an Australian population-based cohort of 4,492 individuals, validation of significant loci in two independent cohorts and meta-analysis of all results. Using robust procedures, we disentangle genetic effects of lipid species from lipoproteins. Integration of multiple datasets, including expression quantitative trait loci (eQTL), methylation QTL (meQTL), and protein QTL (pQTL), and in-depth analysis of significant loci highlights putative susceptibility genes for CAD. We demonstrate robust associations between lipid species and CAD using genetic correlations, polygenic risk scores and phenotypic associations. Many lipid-associated loci show pleiotropy with CAD in colocalization analysis. Assessment of loci with coronary atherosclerosis in 456,486 UK Biobank participants reveals novel associations, independent of clinical lipid measures.

## Results

### Lipidomic profiling

We measured 596 individual lipid species within 33 lipid classes, covering the major glycerophospholipid, sphingolipid, glycerolipid, sterol and fatty acyl classes in serum and plasma samples from three independent cohorts (Supplementary Tables 1-3). Assay performance was monitored using pooled plasma quality control samples, enabling determination of coefficient of variation (%CV) values for each lipid class and species. In the Busselton Health Study (BHS) discovery cohort, the median %CV was 8.6% with 570 (95.6%) lipid species showing a %CV less than 20%. All lipids were measured in every individual, with the exception of three values which were below the limit of detection. The lipidomic analysis of the Australian Imaging, Biomarker, and Lifestyle (AIBL) and Alzheimer’s Disease Neuroimaging Initiative (ADNI) validation cohorts showed similar assay performance^19^.

### GWAS of the human serum lipidome

We performed a GWAS of the human serum lipidome (Figure 1), in the BHS discovery cohort of 4,492 individuals of European ancestry (Supplementary Tables 4-7 and Figure 2) and a meta-analysis of the two validation cohorts, consisting of 670 and 895 individuals of European ancestry (Supplementary Table 8). We further performed a discovery meta-analysis of all three studies (Supplementary Table 9). All summary-level statistics are available at our data portal (https://metabolomics.baker.edu.au/).

**Figure 1.**
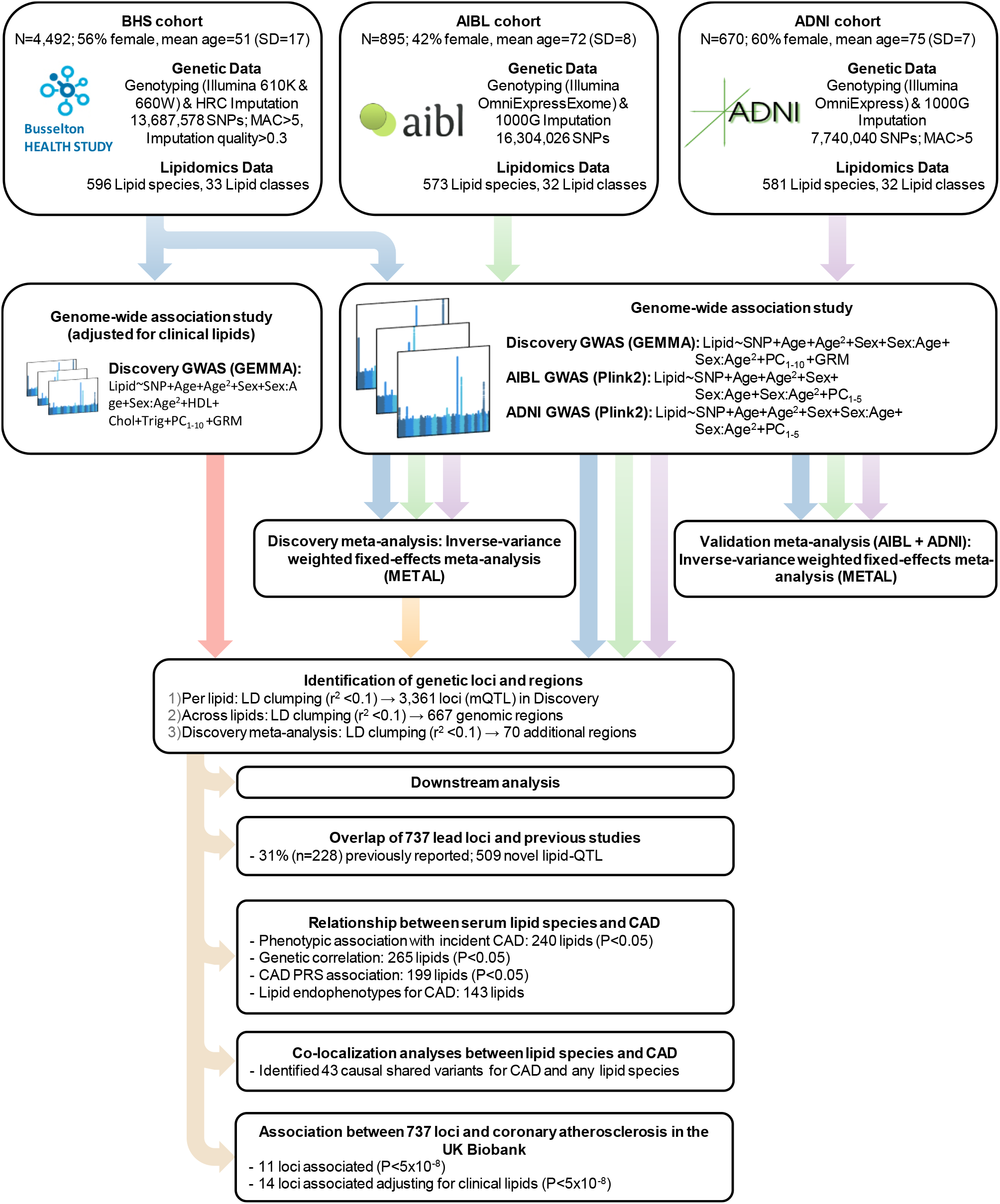
Study design for the genetic analysis of the human lipidome. Representation of genome-wide association studies of the lipidome in the BHS discovery sample, validation and meta-analysis of ADNI and AIBL studies, and downstream analyses.

**Fig. 2.**
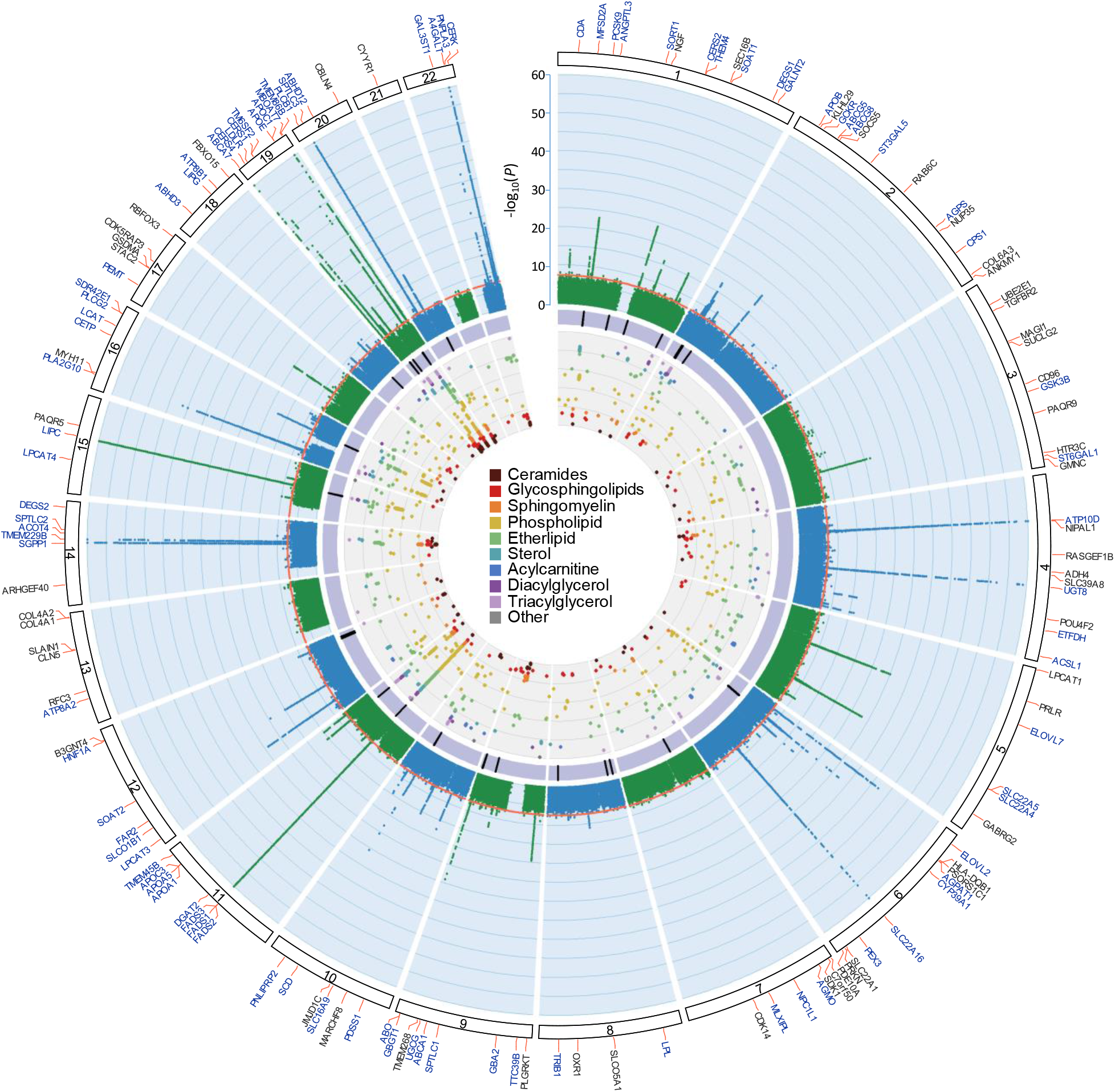
Circular presentation of loci associated with circulating lipid species identified in our Discovery GWAS. The -log_10_(*P*) for genetic association with lipid species are arranged by chromosomal position, indicated by alternating blue and green points. Association *P*-values are truncated at *P*<1×10^−60^. Genome-wide significance (*P*<5×10^−8^) is indicated by the red line. For details about significant associations, see Supplementary Tables 3 and 4. Genes identified in our candidate gene analysis are highlighted in blue, otherwise the closest gene is indicated in black. The purple band indicates lipid loci that colocalize with coronary artery disease (CAD) or show association with CAD after adjusting for clinical lipids. The inner circle shows a Fuji plot of SNP-lipid associations, colored by broad lipid category. Color keys representing broad lipid categories are indicated in the plot center. Chromosomes are indicated by numbered panels 1–22.

The discovery GWAS identified 2,279 independent SNP-lipid species associations, and 132 independent SNP-lipid class associations at a genome-wide significance (P<5.0×10^−8^; r^2^<0.1; Figure 2; Supplementary Table 8). All lipid classes and 543 (of 596; 91.1%) lipid species had at least one significant association. All significantly associated SNPs were in Hardy-Weinberg Equilibrium (HWE; all P≥1.53×10^−4^), and were relatively common (minor allele frequency; MAF<0.01: 4%; MAF>0.05: 91%, Supplementary Table 6). Overall, 667 independent SNPs were significantly associated across lipid outcomes (Supplementary Table 10).

Each SNP was associated with between 1 and 222 lipids (Extended data Fig. 1). SNPs associated with a large number of lipids were in regions known to be involved in lipid regulation, including *FADS1/FADS2/FADS3, APOE*, and *LIPC*. The most significant associations were observed between PC(18:0_20:4) and rs174564 (*FADS2*; P=4.63×10^−220^) and between Cer(d19:1/22:0) and the intergenic SNP rs364585 (flanking *SPTLC3*; P=7.81×10^−185^). In fact, the most significant 26 SNP-lipid species associations were with SNPs in these two regions.

The median genomic inflation factors were 1.01 (range: 0.99-1.03), and 1.02 (range: 1.00-1.03) for lipid species and class analyses, respectively. SNP-based heritability estimates were moderately correlated (r=0.45) with lambda estimates, for each of the lipid species and classes (Extended data Fig. 2a), as expected^20^.

### SNP-lipid species associations are largely independent of clinical lipid measures

We performed additional analyses, adjusting for clinical lipids (total cholesterol, HDL-cholesterol, triglycerides), to identify SNP-lipid species associations independent of clinical lipid traits. The median genomic inflation factors were 1.01 (range: 0.99-1.03), and 1.01 (range: 1.00-1.03) for lipid species and classes, respectively; with heritability estimates moderately correlated (r=0.51) with lambda estimates, for each of the lipid species and classes (Extended data Fig 2b). Adjustment for clinical lipids identified 2,424 independent SNP-lipid species associations, and 124 independent SNP-lipid class associations (Supplementary Table 9). There were 1,545 SNP-lipid species and 72 SNP-lipid class associations that were significant in both the unadjusted and the adjusted analyses, with an r^2^ between beta coefficients of 0.93 (Figure 3; Supplementary Table 4 and 5). Adjustment for clinical lipids identified an additional 879 significant SNP-lipid species associations, for 387 lipid species. However, 726 SNP-lipid species associations previously associated in the unadjusted analysis, fell below our significance threshold. Approximately 24% of these were lipid species in the classes cholesteryl ester (n=93), and phosphatidylcholine (n=81) (Supplementary Table 9). We also identified an additional 52 significant SNP-lipid class associations, particularly for trihexosylceramide (6 associations) and hexosylceramide (6 associations) classes. However, 60 SNP-lipid class associations, fell below our significance threshold, with the classes diacylglycerol, G_M3_ ganglioside, lysophosphatidylcholine, lysoalkenylphosphatidylethanolamine, phosphatidylcholine, alkylphosphatidylethanolamine, alkenylphosphatidylethanolamine, phosphatidylserine, sphingomyelin, and triacylglycerol no longer associated (P<5.0×10^−8^) with any genetic variants.

**Fig. 3.**
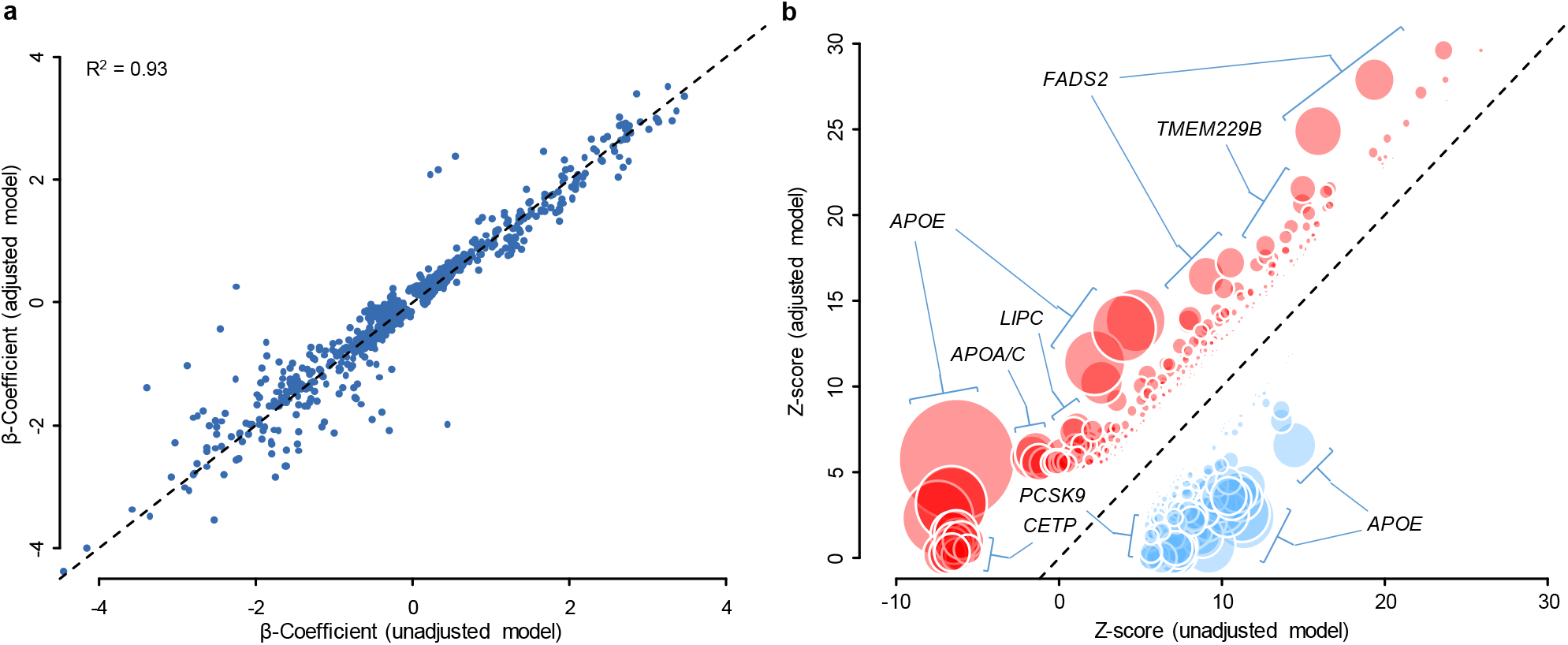
Comparison of estimated lipidomic effect sizes between clinical lipid adjusted and unadjusted models. **a**, Beta coefficients for independent unadjusted SNP-lipid associations (*x* axis) are plotted against clinical lipid adjusted SNP-lipid associations (*y* axis). **b**, Z-scores for unadjusted SNP-lipid associations (*x* axis) are plotted against clinical lipid adjusted SNP-lipid associations (*y* axis). Z-scores for SNP associations reaching genome-wide significance (P<5×10^−8^) in either the clinical lipid adjusted or unadjusted models. Variant effect signs are fixed so adjusted associations are positive. Variants showed greater (positive) associations in clinical lipid adjusted analysis are shown in red, and variants showing reduced associations are shown in blue. Circle diameter is proportional of -log_10_(P) t-test of effect differences.

Results from multi-trait conditional and joint (mtCOJO; Supplementary Tables 4 and 5) analyses using clinical lipid traits (total cholesterol, HDL-cholesterol, triglycerides) GWAS results from the UK Biobank, to minimise the risk of pleiotropy/collider bias introduced by heritable covariates, were largely consistent with those of the clinical lipid adjusted analysis (r^2^ of beta coefficients=0.91, Extended data Fig. 3). Comparison of the clinical lipid adjusted Z-scores and mtCOJO Z-scores identified three regions (*APOE, FADS1*/*FADS2*/*FADS3, TMEM229B*/*PLEKHH1*) with substantial differences (P<1.0×10^−4^) indicating the possibility of biased effect measures for the adjusted analyses in these regions. Overall, results were overwhelmingly consistent between mtCOJO and clinical lipid-adjusted analyses.

Conditional analysis (sequentially conditioning on the lead SNP) identified 386 secondary signals (across both funadjusted and clinical lipid-adjusted analyses), associated with 163 lipid species/classes (Supplementary Table 7). Two regions, *LIPC* and *ATP10D*, each contained five independent signals (P_CONDITIONAL_<5.0×10^−8^). The *LIPC* genomic region was strongly associated with phosphatidylethanolamine species and class, while *ATP10D* was associated with hexosylceramide species and class. The *SPTLC3* region harboured four independent signals, strongly associating with sphingolipids containing a d19:1 sphingoid base.

### Associations validated in independent cohorts

For each lipid, significantly associated SNPs were linkage disequilibrium (LD)-clumped to remove variants in LD(r^2^>0.1). We assessed whether the 2,411 independent lipid species/class associations identified in the BHS discovery cohort (unadjusted analysis) were validated within a combined ADNI and AIBL validation cohort meta-analysis. There were 273 SNP-lipid associations not available for validation in the meta-analysis, either due to lipids not available in the ADNI and AIBL cohorts; missing SNPs (and proxies) on the imputation panel; or monomorphic/very low frequency MAF in ADNI/AIBL. Therefore, we attempted to validate the remaining 2,137 significant SNP-lipid associations (Supplementary Table 8). We considered a SNP-lipid association to be validated if i) the SNP was significantly associated (P<5×10^−8^) in the unadjusted BHS discovery GWAS; ii) the direction of effect was concordant between the validation meta-analysis and the BHS discovery analysis; and iii) the association was nominally significant (P<0.05; less conservative) or reached the Bonferroni significance threshold (P<2.34×10^−5^) in the validation meta-analysis. We identified 1,474 (69.2%) SNP-lipid associations that reached nominal significance (P<0.05), and 644 (30.1%) reaching Bonferroni-corrected significance. Almost all associations (>99%) had the same direction of effect, with a very strong correlation between validation meta-analysis and significant (P<5×10^−8^) discovery effect sizes (r^2^=0.53 overall, and r^2^=0.80 for SNPs with MAF > 0.05 in the BHS; Extended data Fig. 4).

### Discovery meta-analysis

At a stringent significance threshold of P<3.47×10^−10^ (5×10^−8^/144 effective lipid dimensions), the meta-analysis of all three studies identified 65,563 significant SNP-lipid associations (Supplementary Table 9), involving 499 lipid species/classes and 7,600 SNPs. We identified 5,658 new associations not observed in the BHS discovery GWAS alone, involving 352 lipids and 2,914 SNPs. The majority of these (n=5,543; 98%) showed some evidence of association in the BHS discovery GWAS (5×10^−8^< P <5×10^−4^). However, 89 associations were not nominally significant (P>0.05) in the BHS discovery GWAS, indicating that the effects observed in the meta-analysis were largely due to the AIBL and ADNI samples.

### Defining independent loci and genes controlling lipid homeostasis

For each lipid, significantly associated SNPs were LD-clumped to remove variants in LD (r^2^>0.1). Lead variants from the individual analyses (clinical lipid adjusted and unadjusted), including conditional analyses, were clumped if the index SNPs were in linkage disequilibrium (r^2^>0.1). We identified 3,361 independent loci-lipid associations, involving 610 lipid species/classes, each associated with between 1 and 30 independent SNPs. To identify genomic regions associated with lipid metabolism, a single dataset was produced by identifying the smallest P-value for each SNP, across all lipids and analyses. LD-clumping of this dataset resulted in 667 independent genomic regions (Supplementary Table 10). This procedure was repeated, including SNP-lipid associations passing our discovery meta-analysis significance threshold (P<3.47×10^−10^), resulting in 682 independent genomic regions, 612 of which overlap with those identified in BHS alone (737 in total). The variants within a genomic region and the lipids associated with those variants are collectively termed a genetically influenced lipotype.

### Identification of candidate genes within loci

Using the **Pr**ioritization **o**f candidate causal **Ge**nesat **M**olecular QTLs (ProGeM) framework^21^ to prioritize candidate causal genes, biologically plausible genes were identified in 573 of the 737 genomic regions (Supplementary Tables 10-12), with an overlap of 498 genomic regions between genetic-based (bottom-up) and biological knowledge (top-down) based approaches. A total of 2,321 SNP-gene pairs were identified, where the gene has previously been implicated in the regulation of metabolism or a molecular phenotype (Figure 4a). Of these genes, 970 (41.8%) are present in lipid-metabolism specific databases.

**Fig. 4.**
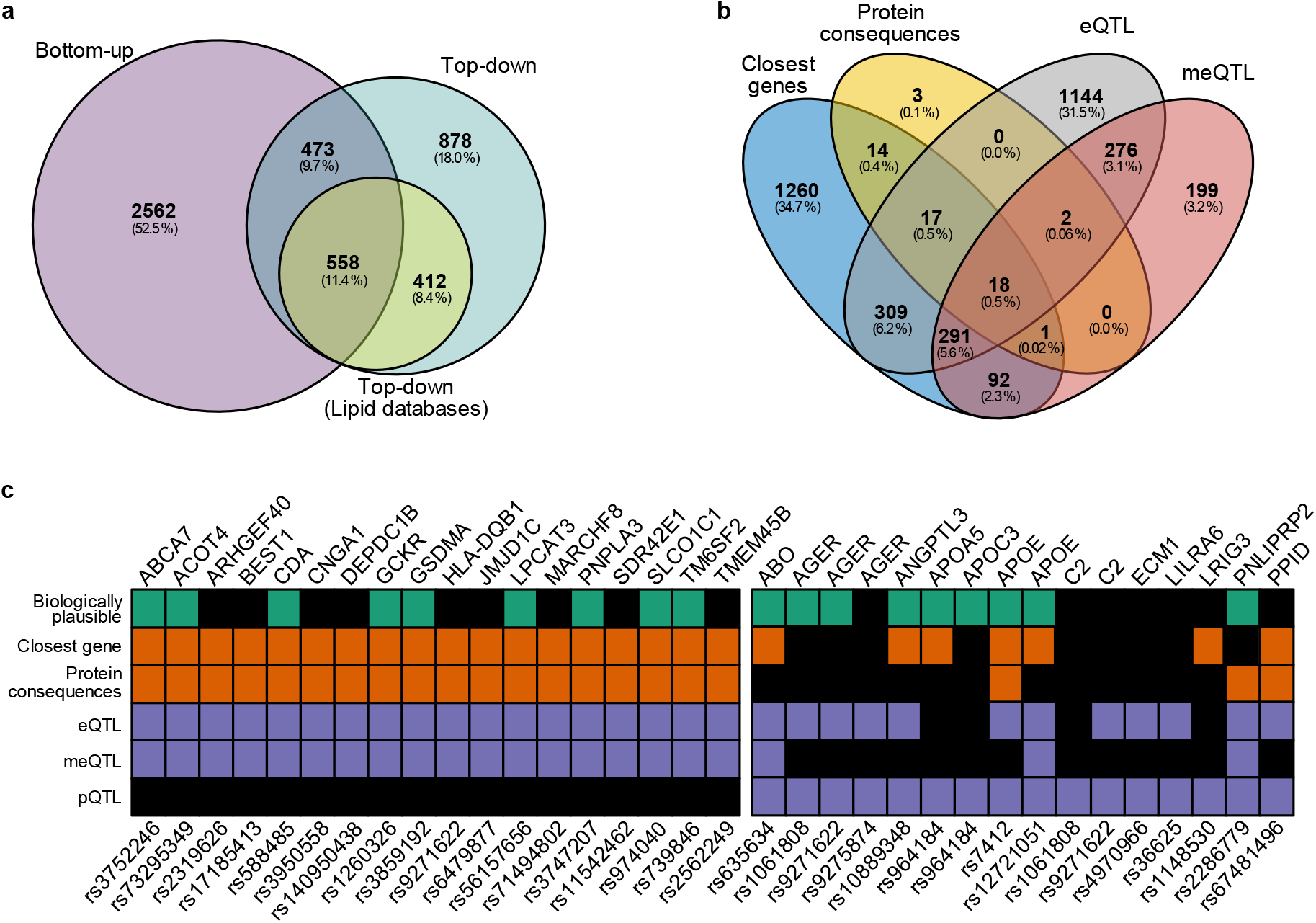
Identification of putative causal genes using genetic prioritization and knowledge-based approaches. Assignment of putative causal genes was performed using the ProGeM framework, incorporating genetic-based prioritization (bottom-up) and biological knowledge-based approaches (top-down). **a**, Venn diagram showing the number of loci with annotations for causal genes using the distinct approaches and the overlap. Top-down annotations were divided into lipid-specific databases and generic databases. **b**, Venn diagram of distinct genes identified in genetic-based prioritization analysis. **c**, summary of putative causal genes with overlapping annotations for closest gene, protein consequences, eQTL and meQTL (left). Summary of putative causal SNP-gene pairs for which pQTL evidence was identified (right).

A total of 62 SNPs were annotated as either missense (n=59), stop gain (n=2), structural interaction (n=1), start loss (n=1), or splice donor (n=1) mutations. Of these, three were annotated as having a putative ‘high’ impact, and the remaining as ‘moderate’ impact. These SNPs are linked to 55 protein products (Figure 4b).

Comparing our lead SNPs and proxies against previously published eQTL associations, 2,058 SNP-gene pairs were identified (Figure 4b). Published meQTL associations revealed 879 SNP-gene pairs, 587 (66.8%) of which replicated eQTL associations. In contrast to eQTL and meQTL, overlap of published pQTL associations were much less evident, with only 16 SNP-gene pairs identified (Figure 4c). In total, 18 SNP-gene pairs were identified with evidence from closest gene, protein consequences, eQTL and meQTL. The overlap of top-down and bottom-up candidates supported the annotation of 1,031 SNP-gene pairs.

### Most SNP-lipid species associations were nove

Of the 737 lead variants (and their proxies), 228 (31%) had been reported in at least one of 35 previous metabolomic/lipidomic studies (Supplementary Note 1), resulting in 509 putatively novel genetically influenced lipotypes (Supplementary Table 13).

### Genetically influenced lipotypes overlap with coronary artery disease and cardiovascular disease related loci

We looked at overlap between 10 hard cardiovascular disease (CVD) points from the GWAS catalog and the lead SNP (or proxy) from each of the 737 regions, identifying a total of 23 lead SNPs, or their proxies, associated (P<5×10^−8^) with 10 hard CVD endpoints (Supplementary Table 14). The most frequently overlapping GWAS catalog hard CVD endpoints were CAD (n=14 SNPs), CVD (n=10 SNPs), coronary artery calcification (n=8 SNPs), and myocardial infarction (n=8 SNPs). Three additional lead SNPs were associated with CAD in the CARDIoGRAMplusC4D and UK Biobank meta-analysis. Eighty-four lead SNPs were associated with 101 CVD-related traits, including chronic kidney disease (n=18,) C-reactive protein (n=14), metabolic syndrome (n=12), body mass index (n=8), and systolic blood pressure (n=4). As expected, lead SNPs frequently overlapped with 186 lipid-related traits, with 99 lead SNPs or proxies observed in the GWAS catalog.

### Serum lipid species/classes are phenotypically and genetically associated with coronary artery disease

Using nominal significance (P<0.05), we identified 240 lipid species/classes phenotypically associated with incident CAD in the BHS (Figure 5a; Supplementary Table 15), with 11% in the positive direction. The strongest association was between TG(50:2)[NL-18:2] and incident CAD (0.311±0.046, P=1.74×10^−11^, FDR q=1.09×10^−8^). Overall, the most strongly associated lipid species were those in the triacylglycerol, diacylglycerol, phosphatidylethanolamine, and cholesteryl ester classes.

**Fig. 5.**
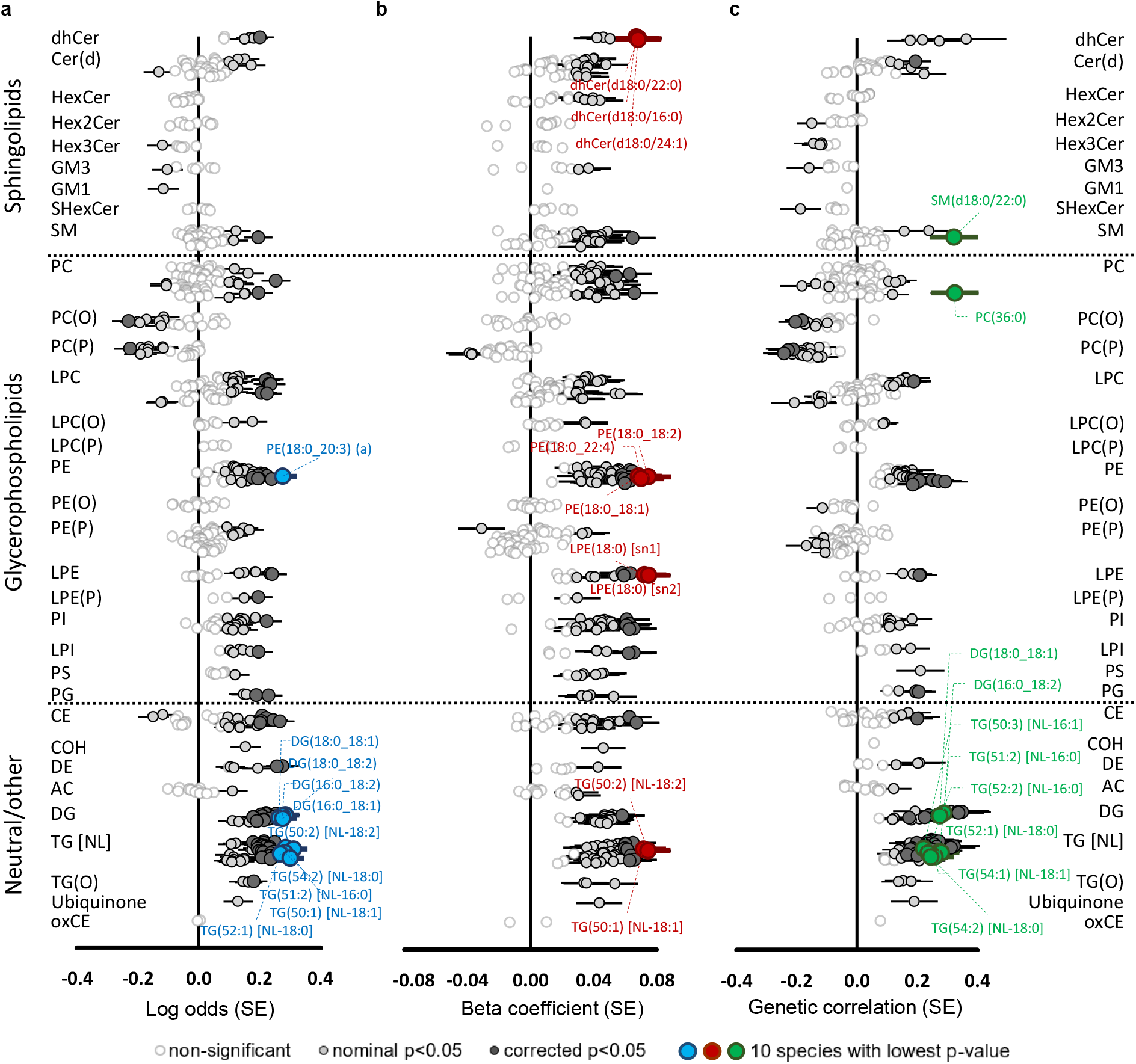
Genetic and phenotypic associations of the lipidome with coronary artery disease. Forest plots of lipid-coronary artery disease effect sizes and standard errors. **a**, phenotypic associations between lipid species and incident coronary artery disease in the BHS cohort (551 cases and 3,703 controls), adjusted for age, sex, and the first 10 genomic principal components. **b**, association of lipid species with polygenic risk for coronary artery disease. Individuals in the discovery cohort (n=4,492) were assessed for risk using the metaGRS polygenic score, consisting of approximately 1.7 million genetic variants. Linear regressions were performed to test the association between an individual’s polygenic score and lipid species concentrations, adjusting for age, sex and the 10 first principal components. **c**, genetic correlations of lipid species against coronary artery disease (meta-analysis of CARDIoGRAMplusC4D and UK Biobank; 122,733 cases and 424,528 controls), performed with Linkage Disequilibrium Score Regression (LDSC; v1.0.1). The 10 most significant lipid species are highlighted in blue, red, or green.

**Fig. 6.**
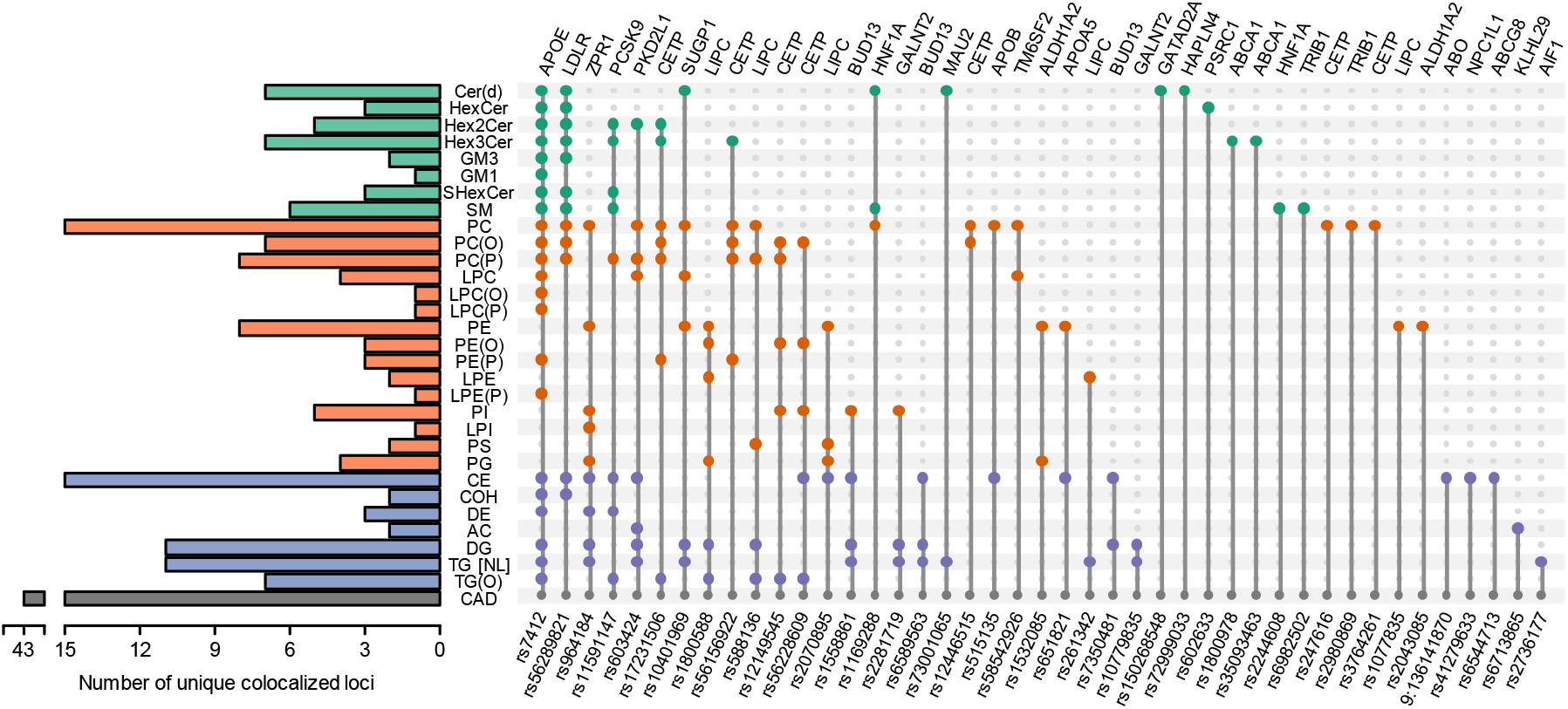
Colocalization of lipid-loci with coronary artery disease. Summary of lipid classes which contain at least one lipid species that colocalizes with coronary artery disease. Colors indicate broad lipid categories. Indicated variants were identified as the most likely causal variant for each of the colocalization analyses. Genetic variants are ordered according to the number of colocalizations across lipid classes. Evidence of colocalization included H3+H4 > 0.8 and H4/H3 > 10. Variants were annotated to the closest gene.

We identified 265 lipid species/classes that showed a nominally significant (P<0.05) association with the CAD polygenic risk score^22^ in the BHS (Figure 5b; Supplementary Table 15). These were positive associations except for lipids in the alkenyl-phosphatidylcholine and alkenyl-phosphatidylethanolamine classes. The strongest association was observed for LPE(18:0) [sn2] (0.075±0.014, P=8.9×10^−8^, FDR q=5.59×10^−5^).

Next, we estimated the genetic correlation between lipid species/classes and CAD. Using linkage disequilibrium score regression, we identified nominally significant genetic correlations (P<0.05) between 199 lipid species/classes and CAD, with 50 of these negatively correlated (Figure 5c; Supplementary Table 14). The strongest genetic correlations were between TG(51:2) [NL-16:0] (0.275±0.058, P=2.22×10^−6^, FDR q=8.94×10^−4^) and CAD.

Overall, using a significance threshold of P<0.05, we identified 134 lipid species/classes that were significantly associated in each of the three analyses - association with incident CVD (phenotypic), CAD polygenic risk (PRS), and genetic correlation. Importantly, these lipid species/classes showed concordant directions of effects in all three analyses, defining these lipid species/classes as lipid endophenotypes for CAD.

### Colocalization analysis identified shared causal variants for coronary artery disease

We performed pairwise colocalization analysis, within each QTL, between lipid species and CAD to assess whether they share common causal variants (Supplementary Table 16). We identified evidence of 43 shared causal variants for CAD and any lipid species (Table 1; Supplementary Note 2). The strongest evidence was between CE(18:1) and CAD at the *APOE* rs7412 loci (H3+H4=1.00; H4/H3=1.17×10^11^). There was strong evidence for the sharing of this causal variant between CAD and 184 lipid species from 23 lipid classes (with and without clinical lipid adjustment). There was also strong evidence for rs603424, near a likely candidate *SCD* (Stearoyl-CoA desaturase), and 24 lipid species/classes (0.936<H3+H4<0.998; 16<H4/H3<1.8×10^3^).

**Table 1.**
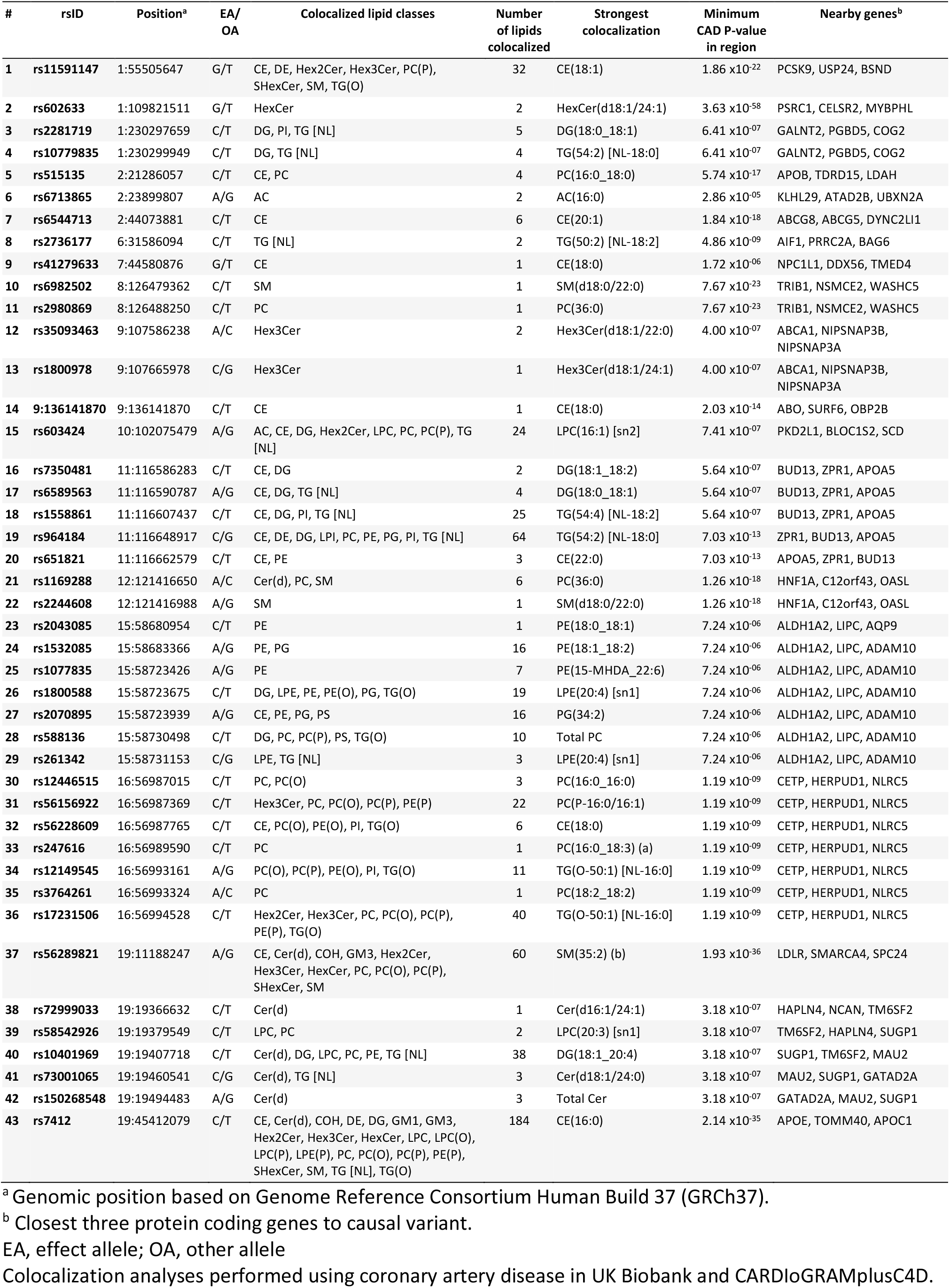
Genomic regions showing colocalization with lipid species and coronary artery disease.

### Genetically influenced lipotypes were associated with coronary atherosclerosis in the UK Biobank

To further define pleiotropic effects between lipid species and CAD, we performed association analysis of 737 lead SNPs and coronary atherosclerosis in 456,486 participants of the UK Biobank (Supplementary Table 17). Eleven of the lipid-associated SNPs had genome-wide significant (P<5×10^−8^) associations with coronary atherosclerosis. Adjustment for clinical lipids (total cholesterol, HDL cholesterol, triglycerides) increased this number to 17; however, adjustment for clinical lipids using mtCOJO, which is free of the bias introduced by heritable covariates, resulted in only 14 associations with coronary atherosclerosis. Importantly, 11 of these associations were sub-genome wide significant in the initial analysis, suggesting the presence of strong pleiotropy in these regions. After comparing effect estimates between the standard GWAS and mtCOJO clinical lipid adjusted analysis, eight lead SNPs (with P<5×10^−8^ in the standard GWAS) showed opposite direction of associations. These regions contain prototypical lipid/lipoprotein regulating genes, such as *APOE, CETP, LDLR*, and *PCSK9*. Interestingly, for all lead SNPs with marginal association with coronary atherosclerosis (P<1.0×10^−3^; with and without conditioning on clinical lipids), 43 (81%) were associated with lipid endophenotypes for CAD.

## Discussion

By integrative analysis of the human lipidome and CAD phenotypes, we have identified putative causal genes for CAD, providing evidence for a causal role of these lipid species in the development of CAD. Our high resolution genome-wide association analyses of the human lipidome has identified 737 independent genomic regions associated with lipid metabolism, of which 509 represent novel genetic loci. This is a substantial increase over previous studies with similar or larger sample sizes^7,10,23^. Our expanded lipidomic platform utilises extensive chromatographic separation to increase the diversity of measured lipid species and distinguish lipid isomers and isotopes over those measured in previous studies. Combined with the extended pedigree study design of the BHS, we identify many rare/low-frequency variants with large effect sizes.

The majority (69.2%) of the 2,137 SNP-lipid associations identified in our discovery GWAS were validated in a meta-analysis of two independent cohorts. Adjustment for clinical lipids (both as standard covariates and mtCOJO analysis), confirmed that the majority of SNP-lipid associations observed were not acting directly through clinical lipids (i.e. associations were not the result of mediated pleiotropy). Meta-analysis of all three studies identified an additional 5,658 SNP-lipid associations (from 122 loci) - involving 352 lipid species - that were not identified in the BHS discovery GWAS alone. Overall, nearly all lipid species (95%) had at least one genome-wide significant SNP association, highlighting the genetic contribution to lipid metabolism and homeostasis.

We identified 134 lipid species/classes showing consistent and significant associations with CAD when assessed with genetic correlation, phenotypic association, and PRS association. These lipids are potential endophenotypes for CAD, which can facilitate the identification of susceptibility genes. Of those loci associated with this subset of lipids, we identified 32 regions with evidence of shared causal genetic effects (colocalization) with lipids and CAD. We assessed the association of lipid-loci with coronary atherosclerosis in ∼456,000 individuals of the UK Biobank, considering independence of clinical lipid traits. A total of 53 loci showed evidence of association (P<1×10^−3^) in at least one analysis. Of these, 43 loci were associated with at least one of the 134 lipid species identified above.

Our lipidomic profiling provided improved resolution and precision in measurement of lipid species. Prior studies examined lipid phenotypes that were mixtures of similar, but distinct species; lacked structural characterization of lipid species; or were contaminated through isotopic overlap. Many of the associations between lipid species and prototypical lipid regulating genes observed in our study - such as *FADS1/FADS2, APOE* and *LDLR* - have been reported in earlier GWAS^7-15,17,23^. With our expanded lipidomic profile, we have built on these earlier studies, identifying many new loci associated with lipid species and classes. Previous studies, containing mis-annotation of lipid species, report associations between SNPs in the *FADS* region and sphingomyelin species as containing a mono-unsaturated (16:1, 18:1 or 20:1) n-acyl chain^8,12^. Here, we show the associations of sphingomyelins with SNPs in the *FADS* region is disproportionally with species containing the d18:2 sphingoid base. This is supported by recent experimental evidence, suggesting FADS3 is a ceramide specific desaturase, targeting the sphingoid bases^24,25^. Early dogma suggested the dominant isoform of sphingomyelins was d18:1 leading to the aforementioned annotations (i.e. SM(d18:1/16:1)). However, chromatographic separation and characterisation identifies the predominant species as SM(d18:2/16:0)^18^. While these associations are not novel *per se*, the additional specificity of our lipidomics methodology extends across all lipid species and classes, leading to greater confidence in defining true relationships.

We also observed strong associations between specific sphingolipid isoforms and variants in the *SPTLC3* gene region. Serine palmitoyltransferase long chain base subunits (SPTLC) are a series of enzymes responsible for the *de novo* synthesis of sphingolipids through condensation of serine with palmitoyl-CoA. Three mammalian isoforms have been identified (SPTLC1-3), which form a heterodimer *in situ*, of which SPTLC1 is requisite for function^26^. The subunit SPTLC3 was discovered more recently and was thought to facilitate the synthesis of shorter-chain sphingolipids^27^. However, we identify strong associations of SNPs in the *SPTLC3* region with atypical sphingolipids, containing a d19:1 sphingoid base (Supplementary Table 5). This supports the recent report that SPTLC3 has broader substrate specificity, with capacity to metabolise branched isomers of palmitate (anteiso-branched-C16)^26^ leading to the synthesis of d19:1 sphingoid bases. The atypical structure of these sphingolipids has previously led to mis-annotation resulting in reported associations of *SPTLC3* with hydroxylated sphingomyelins^10,13,14^, when hydroxylated sphingomyelins in the n-acyl chain are unlikely to exist in human plasma^28^.

Many genes associated with CAD risk were identified as also associated with lipid species and classes, including *HMGCR, PCSK9* and *LDLR* (Table 1), thereby providing new avenues for investigation into causal pathways. We also provide new evidence to support causal roles for genes not reaching genome wide significance, and identify possible mechanisms linking these genes to CAD; we identified strong associations between ten independent signals in the *LIPC/ALDH1A2/AQP9* region with phosphatidylethanolamine, lyso-phosphatidylethanolamine, and phosphatidylglycerol lipid species independent of clinical lipids. Two lead variants were associated with functional consequences, including a start loss for *ALDH1A2* and a missense variant for *LIPC*. The *LIPC* gene on chromosome 15 encodes hepatic lipase, which is functionally described as a triglyceride lipase and as possessing phospholipase A1 activity (hydrolyses sn-1 fatty acid from phospholipids). The role of hepatic lipase in lipoprotein remodelling is complex, being intimately involved in HDL-, IDL-, and chylomicron remnant-metabolism^29^. Consequently, the role of hepatic lipase in cardiovascular disease risk has been controversial, with both pro- and anti-atherogenic mechanisms identified^29,30^. These mechanisms are often viewed through the lens of lipoprotein kinetics. However, the associations of variants in the *LIPC* region with phosphatidylethanolamine species are independent of lipoprotein metabolism (Supplementary Tables 4 and 5) – notionally as these lipids are direct substrates for hepatic lipase. Interestingly, the strength of association of *LIPC* variants with coronary atherosclerosis is considerably increased when conditioned on clinical lipids (both standard adjustment and mtCOJO analyses; Figure 7c, Supplementary Table 17) further supporting a direct mechanistic link. Phenotypically, phosphatidylethanolamine species are associated with incident CAD (Supplementary Table 15), with a direction of effect concordant with the SNP associations (Figure 7a). Visual comparison of regional association plots and SNP effect scatter plot supports consistent effects (Figure 7b and 7d). We selected independent SNPs (r^2^<0.05) in the *LIPC* region associated with the phosphatidylethanolamine class and assessed the similarity of effects with CAD (Figure 7d). Inverse-variance weighted meta-analysis of SNP effects using Generalised Summary-data-based Mendelian Randomisation (GSMR) support strong pleiotropy consistent with a causal relationship (Figure 7e).

**Fig. 7.**
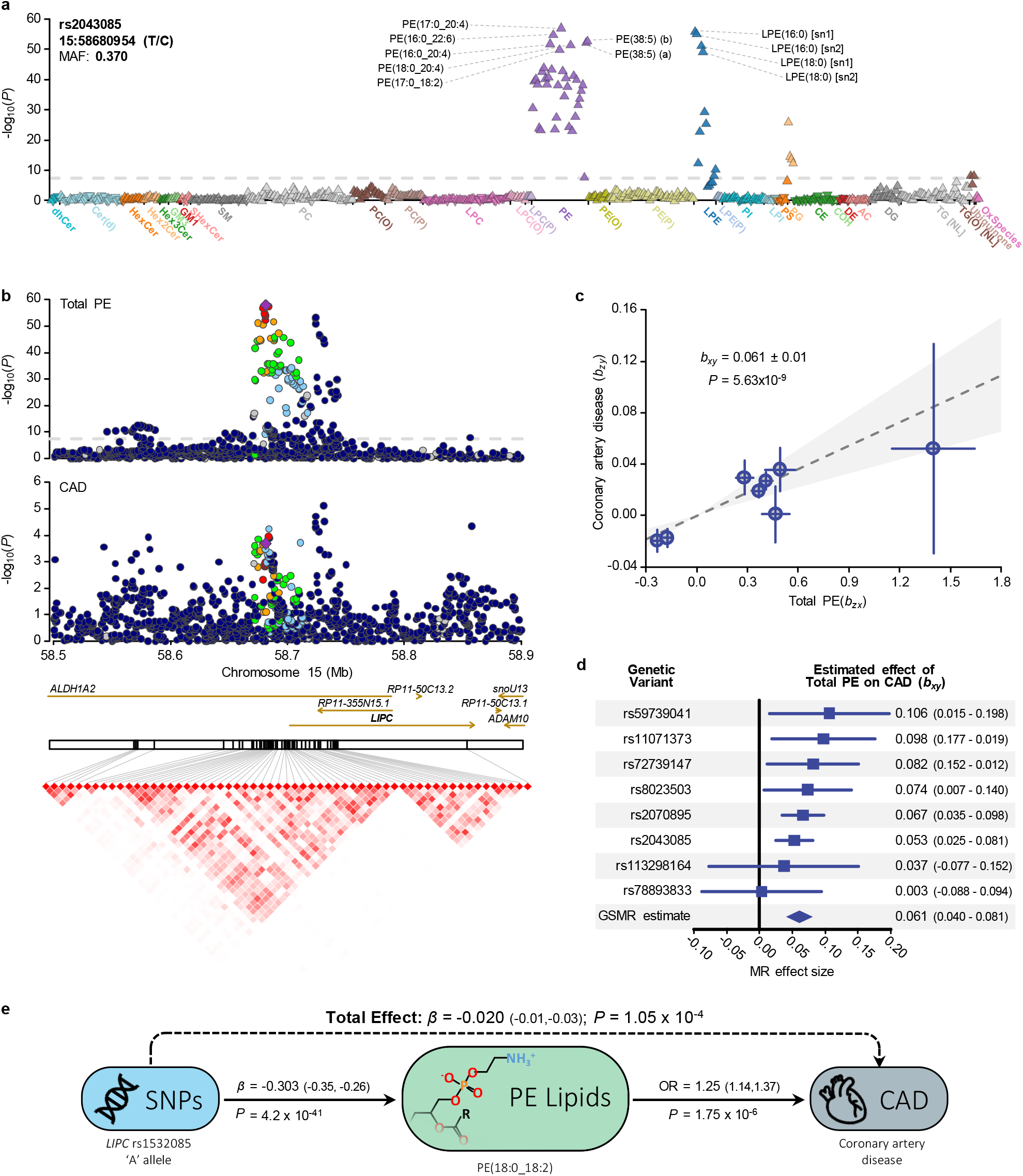
Genetic analysis of the LIPC gene region and circulating levels of phosphatidylethanolamine. **a**, lipid-wide association with the genetic variant, rs2043085, in the BHS cohort (n=4,492). Symbol color is used to distinguish lipid classes. The symbol orientation indicates the effect sign, inverted triangles indicate negative associations, while regular triangles indicate positive associations. The dashed line indicates genome-wide significance (P<5×10^−8^). **b**, regional association plots for Total PE and coronary artery disease (van der Harst & Verweij 2018), focusing on the LIPC region. Variants are colored based on LD with the lead variant, rs2043085. Linkage disequilibrium plot showing correlation between variants following clumping (R^2^>0.8; *P*<5×10^−8^). Variant correlations were obtained from 10,000 unrelated individuals from the UK Biobank. **c**, plot of genetic instrument effect sizes against Total PE and coronary artery disease. Variants were selected based on association with Total PE from within the LIPC region. Eight approximately independent variants were left following clumping (R^2^>0.05; *P*<5×10^−8^). Generalised summary-data based Mendelian randomisation (GSMR) was used to estimate effect of Total PE on coronary artery disease, accounting for the variant correlations and uncertainty in both b_zx_ and b_zy_. **d**, forest plot of single variant tests and GSMR estimate. **e**, diagram of mediated pleiotropy, showing effect sizes estimated across multiple datasets. Exposure modifying variant effect sizes were estimated in the BHS cohort, as well as odds-ratio of phosphatidylethanolamine lipid species against incident cardiovascular disease. Total effect represents the sum of genetics effects on coronary artery disease, whether mediated through phosphatidylethanolamine or not. Coronary artery disease effect size was obtained from van der Harst & Verweij 2018.

Angiopoietin-like 3 (ANGPTL3) has been implicated in CAD risk, with a deficiency being associated with cardioprotective effects^31-33^. ANGPTL3 acts as an inhibitor to two other lipases, lipoprotein lipase (LPL) and endothelial lipase (LIPG); loss of function mutations in *ANGPTL3* have been linked to hypolipidemia^33^. We recently identified a rare frameshift deletion (rs398122988) associated with decreased ANGPTL3 protein levels in extended Mexican American families^34^; the variant was also associated with a ∼1.3 standard deviation decrease in phosphatidylinositol species. In this study, we validate this observation, with SNPs in the *ANGPTL3* region associated with a decrease in phosphatidylinositol species, again these associations persisted even after adjustment for clinical lipids (total cholesterol, HDL-C, triglycerides). Interestingly, we also observe associations of phosphatidylinositol species with SNPs in the *LIPG* region. Commonly, phosphatidylinositol species have been studied for their intracellular messaging roles following phosphorylation of the inositol ring by kinases, including PI-3-kinase, which lead to downstream cardio-metabolic effects^35^. However, the role of phosphatidylinositol species in CVD risk is still largely unknown; we have previously observed the change in the ratio of phosphatidylinositol to phosphatidylcholine species as a predictor of CVD risk reduction from statin treatment^36^. Further work is now required to unravel the role on phosphatidylinositol in mediating the effect of these genes on CVD risk.

In summary, using our expanded lipidomic profiling platform, we have investigated the largest number of targeted lipid species in a GWAS, and have reported significant genetic associations with lipid species that have not previously been reported in any genetic association studies to date. Our strategy to use lipid species as endophenotypes in the search for CVD genes is the ‘tip of the iceberg’. We have previously reported phenotypic associations of lipid species with other complex traits, including diabetes^37^, Alzheimer’s disease^19^, and atrial fibrillation^38^; we believe the same integrative genomics approach may now be used to elucidate the mechanistic underpinnings of lipid metabolism in these and other complex diseases. These data now represent a valuable resource for the future exploration of the genetic analysis of the lipidome to identify lipid metabolic pathways and regulatory genes associated with complex disease and identify new therapeutic targets. To this end we provide all summary statistics and an online searchable resource of association plots of lipid species and classes with genetic variants and regional association plots with individual lipid species and classes (https://metabolomics.baker.edu.au/).

## Methods

### Study populations

Participants in the discovery cohort (n=4,492) were all participants of the 1994/95 survey of the long-running epidemiological study, the BHS, for whom genome-wide SNP data, extensive longitudinal phenotype data, and blood serum were available. The BHS is a community-based study in Western Australia that includes both related and unrelated individuals (predominantly of European ancestry), and has been described in more detail elsewhere^39-41^. Informed consent was obtained from all participants and the 1994/95 health survey was approved by the University of Western Australia Human Research Ethics Committee (UWA HREC). The current study was also approved by UWA HREC (RA/4/1/7894) and the Western Australian Department of Health HREC (RGS03656).

The two validation cohorts used in this study were the AIBL study^42^ and the ADNI study^43^; both of which were established to discover biomarkers, health and lifestyle factors for the development, early detection, and tracking of Alzheimer’s disease. The AIBL study is a longitudinal study which recruited 1,112 individuals aged over 60 years within Australia. Time points for blood/data collection were every 18 months from baseline. For each individual, lipidomic data obtained from the earliest blood collection was used. At baseline, 768 individuals were characterized as cognitively normal, 133 with mild cognitive impairment and 211 with Alzheimer’s disease. The ADNI study is a longitudinal study, starting in 2004 and recruited 800 individuals at baseline, from sites across the United States of America and Canada. Serum samples obtained at baseline were analysed. Study data analysed here were obtained from the ADNI database, which is available online (http://adni.loni.usc.edu/). For the lipidomics analysis, the AIBL study was deemed low risk (The Alfred Ethics Committee; Project 183/19), and the ADNI study was deemed ‘research not involving human subjects’ (Duke Institute review board; ID:Pro00053208).

### Lipidomic profiling

Targeted lipidomic profiling was performed using liquid chromatography coupled electrospray ionization-tandem mass spectrometry to quantify 596 lipid species from 33 lipid classes, from non-fasting blood serum (BHS discovery) and non-fasting blood plasma (ADNI and AIBL validation). Lipidomic profiling of each cohort was performed using the methodology described by Huynh *et al*. and has been described previously^18,44^. Briefly, 10μL of serum was spiked with an internal standard mix (Supplementary Table 2) and lipid species were isolated using a single phase butanol:methanol (1:1; BuOH:MeOH) extraction^45^. Analysis of serum extracts was performed on an Agilent 6490 QqQ mass spectrometer with an Agilent 1290 series HPLC, as previously described. Mass spectrometry settings and transitions for each lipid class are shown in Supplementary Table 2. A total of 497 transitions, representing 596 lipid species, were measured using dynamic multiple reaction monitoring (dMRM), where data was collected during a retention time window specific to each lipid species. Raw mass spectrometry data was analysed using MassHunter Quant B08 (Agilent Technologies).

### Data integration and cleaning

Lipid concentrations were calculated by relating the area under the chromatographic peak, for each lipid species, to the corresponding internal standard. Correction factors were applied to adjust for differences in response factors, where these were known^18^. In-house pipelines were used for quality control and filtering of lipid concentrations. Across the entire dataset, only three missing values were evident. Lipids below the limit of detection (missing values) were imputed to half the minimum observed value. To remove technical batch variation, the lipid data in each analytical batch (approximately 486 samples per batch; 11 batches in total) was aligned to the median value in pooled plasma quality control samples included in each analytical run. Unwanted variation was identified using a modified remove unwanted variation-2 (RUV-2) approach^46^. In brief, lipid data were residualized in a linear mixed model, against age, sex, body mass index (BMI), clinical lipids and the genetic relatedness matrix (described below) as the random effects. Principal component analysis was performed on the residualized data. The first two components showed clear trends along samples in collection order. Therefore, variation associated with these first two principal components was removed from the original data set. Lipid class totals were generated by summing the concentration of the individual species within each class. Validation cohorts were processed in a similar manner.

### Phenotypic variables

Details of the BHS data collection have been published previously^47^. Serum cholesterol and triglycerides were calculated by standard enzymatic methods on a Hitachi 747 (Roche Diagnostics, Sydney, Australia) from fasting blood collected in 1994/95. HDL-C was determined on a serum supernatant after polyethylene glycol precipitation using an enzymatic cholesterol assay and LDL-C was estimated using the Friedewald formula^48^. Height and weight (used to calculate BMI) were collected from participants at time of interview (1994/95). Use of lipid-lowering medication was recorded at the time of interview (1994/95). Diagnosis of incident CAD was defined as either hospitalisation or death due to CAD (ICD9: 410-414; ICD10: I20-I25) after blood collection date (and until June 2015). Hospitalisations and deaths were identified from the Western Australian Department of Health Hospital Morbidity Data Collection and Death Registrations.

### Medication usage adjustment

For individuals taking lipid-lowering medication (BHS, n=108; AIBL, n=366; ADNI, n=382), lipid species and clinical lipid concentrations were adjusted using previously identified effects of lipid-lowering medication. Changes in lipid species and clinical lipids following one year of statin use were calculated from a placebo randomised controlled trial (LIPID study; n=4991)^36^. To calculate correction factors, lipid measures were centred and scaled by the mean and standard deviation of baseline measures (prior to statin usage), and the change in lipid abundance was calculated and regressed on age, sex, BMI and statin usage. Statin usage beta coefficients (effect of the lipid-lowering medication) was added to standardised lipid species concentrations of the individuals taking lipid-lowering medication in the current study. For lipid species present in both this study and the LIPID study (overlap of 314 lipid species), species-specific correction factors were calculated. For those lipid species not measured in the LIPID study (n=282), class-specific corrections were calculated.

### Genotyping and Imputation

For the BHS discovery cohort, genotyping was performed on the Illumina Human 610K Quad-Bead Chip (Illumina Inc., San Diego, CA, USA) at the Centre National de Genotypage in Paris, France (n=1468), and on the Illumina 660W Quad Array Bead Chip (Illumina Inc., San Diego, CA, USA) at the PathWest Laboratory Medicine WA (Nedlands, WA, Australia (n=3428). Complete linkage clustering based on pairwise identity by state distance in PLINK^49^ showed no batch effects, therefore the batches were merged. Standard genotype data quality control was performed as described previously^41^. Briefly, individuals were excluded if: >3% of SNP data were missing (*n* = 11), reported sex did not match genotyped sex (*n* = 48), duplicates (*n* = 123), missing phenotype data (*n* = 11), or >5 standard deviations above/below mean heterozygosity (*n*=28). Individuals with non-European ancestry (*n*=4) were also excluded. To prepare genotype data for imputation, SNPs were excluded if: call rates < 95%, minor allele count < 10, deviations from HWE (P<5.0×10^™4^), no matching Haplotype Reference Consortium (HRC) reference panel SNP, palindromic (A/T, G/C) SNPs with MAF greater than 0.4 from the HRC (*n*=5), and SNPs with >0.2 MAF difference compared to HRC (*n*=150). After quality control, SNP data was available for 513,634 SNPs. Imputation was performed to the HRC reference panel using the Michigan Imputation Server^50^. Following imputation, 39,117,105 SNPs were available for analysis. We excluded variants if the number of copies of the minor allele <5 or if imputation quality (r^2^) <0.3. This resulted in 13,887,524 variants available for analysis.

Genotyping in ADNI was performed on the Human 610-Quad BeadChip (Illumina, Inc., San Diego, CA). Following standard quality control procedures performed in Plink^49^ (minimum SNP and individual call rate > 95%, MAF>0.05, HWE test P>1×10^™6^), the sample was imputed to the 1000 Genomes Phase 3 reference panel using Impute2^51^, with pre-phasing using ShapeIT^52^.

Genotyping in AIBL was performed on the Infinium OmniExpressExome array (Illumina, Inc., San Diego, CA)^53^. Quality control procedures were performed in Plink^49^. After removing individuals with ambiguous sex, Plink was used to remove individuals with call rate <0.90; SNPs were removed if call rate<0.95, HWE test P<1.0×10^−4^, or MAF<0.05. SNPs were flipped to the positive strand before imputation to the 1000 Genomes Phase 3 reference panel using the Michigan Imputation Server^50^ (using Minimac 4). Both the AIBL and ADNI validation cohorts were restricted to individuals of non-Hispanic European ancestry, based on projection onto the 1000 genomes reference panel.

### Genetic relatedness matrix

The discovery sample, BHS, used in this study consisted of related and unrelated individuals; therefore, all analyses included a genetic relatedness matrix. Twenty-two genetic relatedness matrices were calculated. First, a hard-call set of imputed SNPs was created in Plink (i.e. SNP genotypes were called if SNP imputation quality r^2^>0.8 and if genotype probability >0.9). The *HLA* region on chromosome 6 was also excluded. SNPs were then pruned in Plink using ‘indep-pairwise 500 50 0.3’ [window of size 500, moving 50 SNPs along each time, removing variants with r^2^>0.3] to create a set of 486,553 independent SNPs. Twenty-two genetic relatedness matrices were created (using the option ‘gk 1’ which specifies a centred relatedness matrix), with each omitting one chromosome, in GEMMA^54^.

### Statistical analysis

Genome-wide association analyses for the 596 lipid species and 33 lipid classes in the discovery cohort were performed using imputed genotype dosages in linear mixed models, as implemented in GEMMA^54^. To avoid proximal contamination, analyses were performed using genetic relatedness matrices implementing a leave-one-chromosome out scheme. Analyses were performed using rank-based inverse normal transformed residuals, after adjustment by age, sex, age^2^, age*sex, age^2*^sex and the first 10 principal components (generated from Eigenstrat)^55,56^.

Validation cohorts, ADNI and AIBL, were analysed using an additive linear model, as implemented in Plink^49^. Analyses were performed using rank-based inverse normal transformed residuals, after adjustment by age, sex, age^2^, age*sex, age^2*^sex, study-specific covariates and a number of principal components deemed sufficient to capture population structure. Meta-analysis between all three studies was performed using an inverse-variance weighted fixed-effects model, as implemented in METAL^57^. Due to the correlation between lipid species, the effective number of tests was calculated as the number of principal components required to explain at least 95% variance of the lipidome (144 components).

Statistical significance was defined using the standard genome-wide significance (P<5×10^−8^) in the BHS discovery analysis, P<0.05 in AIBL/ADNI validation, and P<3.47×10^−10^ in the three-study meta-analysis (5×10^−8^/144 lipid dimensions; Bonferroni correction using the effective number of tests). A more stringent threshold was used for the meta-analysis due to the lack of validation samples available.

For each lipid, significantly associated SNPs were LD-clumped (r^2^>0.1) using correlation measures obtained from 10,000 unrelated individuals from the UK Biobank, the 1000 Genomes, or the BHS. A singular dataset was created by retrieving the smallest P-value across all analyses. This dataset was LD-clumped (r^2^>0.1) to determine the number of independent genomic regions. For each locus, a regional association plot was produced using LocusZoom^58^.

### Detection of distinct association signals

Conditional analysis was performed to detect independent association signals at each genome-wide significant loci, using GEMMA. For each lipid, we iteratively clumped regions within a 2Mb window centered on the lead SNP until no more genome-wide significant associations were left. Regions with overlapping windows were merged. Conditional analysis was iteratively performed, including the lead variant as a covariate until no more conditionally independent signals (P<5×10^−8^) remained.

### Assessment of effects of clinical lipid trait adjustment

Within the discovery cohort, to determine whether SNP-lipid associations were independent of clinical lipid traits (total cholesterol, HDL-C, triglycerides), all SNPs were tested with and without adjustment for clinical lipid traits. We compared loci effect sizes between analyses run with and without clinical lipid adjustment using a pooled standard deviation t-test (Supplementary Note 3). Bonferroni adjustment (0.05/number of loci) was used to identify loci which differed substantially following adjustment. As adjusting for heritable covariates can introduce collider bias^59^, we further validated these using multi-trait conditional and joint analysis (mtCOJO)^60^, conditioning on GWAS summary-level data for clinical lipids obtained from the UK Biobank^61^.

### Annotation

Proxies for lead SNPs were found by identifying those in high LD (r^2^>0.8) within the BHS dataset; in an unrelated subset of white, British individuals from the UK Biobank^62^; or in the 1000 Genomes. Lead SNPs and their proxies were annotated using SNPEff^63^. SNiPA database v3.3^64^ was used to retrieve combined annotation dependent depletion (CADD) score. Expression QTL associations (cis-eQTL) were obtained from GTEx^65^ (release v8) and eQTLGen^66^ (release 2019-12-20). SNiPA metabolite QTL (mQTL) associations were supplemented with mQTL associations reported in PhenoScanner^67,68^ and recently published lipidomic GWAS^7,17^. SNiPA protein QTL (pQTL) associations were supplemented with cis-pQTL associations from Emilsson *et al*. 2018^69^. Methylation QTL (meQTL) associations were obtained from Huan *et al*. 2019^70^. A locus was defined as novel if the lead SNP or its proxies were not previously reported as an mQTL or lipid related trait loci.

Putative causal genes, for each loci, were identified using a slightly modified approach to that previously described (ProGeM)^21^. For the bottom-up approach, the three closest protein coding genes (within a 1Mb window) were identified, for each lead SNP. Genes were noted if a lead SNP or its proxies were annotated by SNPEff as missense, start loss, stop gain, or with an annotation impact as High. As performed by ProGeM, the top-down analysis reports genes within 500kb of the lead SNP that are present in a curated database of known metabolic-related genes. A list of primary candidates was generated based on the overlap of top-down and bottom-up genes.

### Overlap of lead variants with cardiovascular disease-related loci

To assess whether our lead SNPs were previously associated with CVD-related traits, we performed a look-up within the GWAS catalog v1.02 (release 2020-08-26)^71^ of 10 hard CVD endpoints, 72 CVD-related traits, and 141 lipid-related traits. We also performed a look up against a meta-analysis of CAD between CARDIoGRAMplusC4D and UK Biobank^72^.

### Associations of lipid species with coronary artery disease and coronary artery disease polygenic risk

Within the discovery cohort, the association of lipid species with incident CAD was assessed using logistic regression, adjusting for age, sex, and the first 10 genomic principal components. Prevalent CAD cases were removed prior to analysis; defined as individuals hospitalised with CAD between the start of the Hospital Morbidity Data Collection (1970), and an individual’s serum collection date. Incident CAD events (CAD hospitalisations or death) were included up to the end of follow-up (July 2015). Results are displayed as log-odds ratios.

Polygenic risk for CAD was calculated for each individual in the discovery cohort using the metaGRS polygenic score, consisting of approximately 1.7 million genetic variants^22^. Linear regression in R was performed to test the association between an individual’s polygenic score and lipid species concentrations, adjusting for age, sex and the 10 first principal components.

### Genetic correlations

Genetic correlations of lipid species against CAD was assessed using Linkage Disequilibrium Score Regression (v1.0.1)^73^. Regression weights and scores were obtained from 1000 Genomes European data, as previously described^74^. Summary statistics from all datasets were restricted to SNPs from the HapMap 3 panel, with 1000 Genomes European MAF greater than 5%. Where available, SNPs were filtered to an imputation quality r^2^ > 0.9. Similarly, SNPs were removed if the reported MAF deviated from 1000 Genomes European MAF by greater than 0.1. Summary statistics for CAD were obtained from the meta-analysis of CARDIoGRAMplusC4D and UK Biobank by van der Harst and Verweij^72^. Due to no overlapping samples between BHS and other summary results, the genetic covariance intercept was constrained to 0.

### Colocalization analysis

Colocalization between lipid species genome-wide significant loci and CAD was performed using the R package COLOC^75^. For each loci, all variants within a 400kb window centered on the lead SNP were selected. Priors were kept at default settings. Evidence for shared causal variants was determined as the posterior probability of both traits containing causal variants in the region (H3+H4>0.8) and a larger probability of a shared causal variant (H4/H3>10). Sensitivity analysis for regions with causal variants are shown in Supplementary Note 2.

### Association of loci with coronary atherosclerosis in the UK Biobank

Lead SNPs (or proxies) were tested for association with coronary atherosclerosis in the UK Biobank. In a subset of white, British individuals (n=456,486), electronic health records (updated 14^th^ December 2020) were converted into PheCodes^76,77^ using the R package PheWAS^78^. Coronary atherosclerosis (phecode 411.4) was exported for genome-wide association analysis. FastGWA^79^ was used to assess the association of lipid-loci with these phenotypes, adjusting for age, sex, age^2^, age*sex, age^2*^sex, the first 20 principal components as provided by the UK Biobank, and the genetic relatedness matrix as the random effect. The analysis was repeated, additionally adjusting for clinical lipids (total cholesterol, HDL-cholesterol, triglycerides; measurements obtained from the first available blood collection). Individuals with missing values were excluded from the analysis. As clinical lipids are heritable, mtCOJO analysis was also performed using GWAS summary statistics obtained above.

## Data Availability

Complete summary statistics of all lipid species and classes will be available via the NHGRI-EBI GWAS catalog (https://www.ebi.ac.uk/gwas), GCP ID: GCP000197; study accession nos. GCST90023981-GCST90025848. In addition, summary-level statistics are available at our data portal (https://metabolomics.baker.edu.au/).
Individual-level data for the BHS are accessible through applications to the Busselton Population Medical Research Institute (http://bpmri.org.au/research/database-access.html). Individual-level data for the ADNI and AIBL studies are available through applications to the LONI Image and Data Archive (http://adni.loni.usc.edu/data-samples/access-data/). Individual-level data for AIBL are also available through applications to the AIBL management committee (https://aibl.csiro.au/research/support/).
Publically available datasets used within the study are available via UK Biobank (http://www.ukbiobank.ac.uk/register-apply/), HRC (http://www.haplotype-reference-consortium.org/home), 1000 Genomes (https://www.internationalgenome.org/), SNiPA (https://snipa.helmholtz-muenchen.de/snipa3/), GTEx (https://gtexportal.org/home/), and eQTLGen (https://www.eqtlgen.org/).

http://bpmri.org.au/research/database-access.html

http://adni.loni.usc.edu/data-samples/access-data/

https://aibl.csiro.au/research/support/

## Data availability

Complete summary statistics of all lipid species and classes will be available via the NHGRI-EBI GWAS catalog (https://www.ebi.ac.uk/gwas), GCP ID: GCP000197; study accession nos. GCST90023981– GCST90025848. In addition, summary-level statistics are available at our data portal (https://metabolomics.baker.edu.au/).

Individual-level data for the BHS are accessible through applications to the Busselton Population Medical Research Institute (http://bpmri.org.au/research/database-access.html). Individual-level data for the ADNI and AIBL studies are available through applications to the LONI Image and Data Archive (http://adni.loni.usc.edu/data-samples/access-data/). Individual-level data for AIBL are also available through applications to the AIBL management committee (https://aibl.csiro.au/research/support/).

Publically available datasets used within the study are available via UK Biobank (http://www.ukbiobank.ac.uk/register-apply/), HRC (http://www.haplotype-reference-consortium.org/home), 1000 Genomes (https://www.internationalgenome.org/), SNiPA (https://snipa.helmholtz-muenchen.de/snipa3/), GTEx (https://gtexportal.org/home/), and eQTLGen (https://www.eqtlgen.org/).

## Code availability

All software and bioinformatic tools used in the present study are publicly available.

## Acknowledgements

Support was provided by the National Health and Medical Research Council of Australia (#1101320 and 1157607). K.H. was supported by a Dementia Australia Research Foundation Scholarship. This work was also supported in part by the Victorian Government’s Operational Infrastructure Support Program, and the Royal Perth Hospital Research Foundation.

The BHS acknowledges the generous support for the 1994/95 Busselton follow-up studies from HealthWay, the Department of Health, PathWest Laboratory Medicine of WA, The Great Wine Estates of the Margaret River region of Western Australia, the Busselton community volunteers who assisted with data collection, and the study participants from the Shire of Busselton.

Statistical analyses performed in this work were supported by resources provided by The Pawsey Supercomputing Centre with funding from the Australian Government and the Government of Western Australia. The authors wish to thank the staff at the Western Australian Data Linkage Branch and Death Registrations and Hospital Morbidity Data Collection for the provision of linked health data.

Funding for the AIBL study was provided in part by the study partners [Commonwealth. Scientific Industrial and research Organization (CSIRO), Edith Cowan University (ECU), Mental Health Research institute (MHRI), National Ageing Research Institute (NARI), Austin Health, CogState Ltd]. The AIBL study has also received support from the National Health and Medical Research Council (NHMRC) and the Dementia Collaborative Research Centres program (DCRC2), as well as funding from the Science and Industry Endowment Fund (SIEF) and the Cooperative Research Centre (CRC) for Mental Health—funded through the CRC Program (Grant ID:20100104), an Australian Government Initiative. Support for AIBL genetic data acquisition and analysis was provided by a grant from the NHMRC (APP1161706) awarded to S.M.L and through the CRC for Mental Health (Grant ID:20100104). T.P. is supported by ECU strategic research funding.

Support for the metabolomics sample processing, assays and analytics reported here was provided by grants from the National Institute on Aging (NIA); NIA supported the Alzheimer’s Disease Metabolomics Consortium which is a part of NIA’s national initiatives AMP-AD and M2OVE-AD (R01 AG046171, RF1 AG051550, RF1 AG057452 and 3U01 AG024904-09S4). Additional NIH support from the NIA, NLM and NCI for analysis includes P30 AG10133, R01 AG19771, R01 LM012535, R03 AG054936, R01 AG061788, K01 AG049050 and R01 CA129769. M.A. is supported by National Institute on Aging grants RF1 AG057452, RF1 AG058942, RF1 AG059093, 1U19AG063744 and U01 AG061359. K.N. is supported by NLM R01 LM012535 and NIA R03AG054936. Data collection and sharing for the ADNI was supported by National Institutes of Health Grant U01 AG024904. ADNI is funded by the National Institute on Aging, the National Institute of Biomedical Imaging and Bioengineering, and through generous contributions from the following: AbbVie, Alzheimer’s Association; Alzheimer’s Drug Discovery Foundation; Araclon Biotech; BioClinica, Inc.; Biogen; Bristol-Myers Squibb Company; CereSpir, Inc.; Cogstate; Eisai Inc.; Elan Pharmaceuticals, Inc.; Eli Lilly and Company; EuroImmun; F. Hoffmann-La Roche Ltd and its affiliated company Genentech, Inc.; Fujirebio; GE Healthcare; IXICO Ltd; Janssen Alzheimer Immunotherapy Research & Development, LLC; Johnson & Johnson Pharmaceutical Research & Development LLC; Lumosity; Lundbeck; Merck & Co., Inc.; Meso Scale Diagnostics, LLC; NeuroRx Research; Neurotrack Technologies; Novartis Pharmaceuticals Corporation; Pfizer Inc.; Piramal Imaging; Servier; Takeda Pharmaceutical Company; and Transition Therapeutics. The Canadian Institutes of Health Research is providing funds to support ADNI clinical sites in Canada. Private sector contributions are facilitated by the Foundation for the National Institutes of Health (www.fnih.org). The grantee organization is the Northern California Institute for Research and Education, and the study is coordinated by the Alzheimer’s Therapeutic Research Institute at the University of Southern California. ADNI data are disseminated by the Laboratory for Neuro Imaging at the University of Southern California. This study was only possible with the help of the AIBL research group. The authors who made direct contribution to this study have been listed as authors in this article. Members of the AIBL group who did not participate in the analysis or writing of this report are listed here: https://aibl.csiro.au/about/aibl-research-team/. Part of the data used in preparation of this article were obtained from the Alzheimer’s Disease Neuroimaging Initiative (ADNI) database (adni.loni.usc.edu). The authors who made direct contribution to this study have been listed as authors in this article. As such, the investigators within the ADNI contributed to the design and implementation of ADNI and/or provided data but did not participate in analysis or writing of this report. A complete listing of ADNI investigators can be found at: http://adni.loni.usc.edu/wpcontent/uploads/how_to_apply/ADNI_Acknowledgement_List.pdf. Part of the data used in preparation of this article were generated by the Alzheimer’s Disease Metabolomics Consortium (ADMC). The authors who made direct contribution to this study have been listed as authors in this article. Investigators within the ADMC provided data but did not participate in analysis or writing of this report can be found at https://sites.duke.edu/adnimetab/team/.

Metabolomics data and results from the ADNI study have been made accessible through the AMP-AD Knowledge Portal (https://ampadportal.org). The AMP-AD Knowledge Portal is the distribution site for data, analysis results, analytical methodology and research tools generated by the AMP-AD Target Discovery and Preclinical Validation Consortium and multiple Consortia and research programs supported by the National Institute on Aging.

## Author contributions

Design of study and interpretation of results: GC, CG, PEM, KH, MI, NSM, JHung, JBeilby, MPD, GFW, SS, NRW, JBlangero, PJM, EKM. Statistical and bioinformatic analyses: GC, CG, PEM, MB, AA. Lipidomic analysis: KH, NAM, TD, AN, MC, AS, GO, TW. Cohort oversight, phenotyping or genotyping: JHung, JHui, JBeilby, WLFL, PC, IM, SML, TP, MV, AIB, CRC, VLV, DA, CLM, KT, MA, GK, KN, AJS, XH, RKD, RNM, PJM, EKM. Drafted the manuscript: GC, CG, PEM, KH, PM, EKM, PJM. All authors read, edited and approved the final version of the manuscript.

## EXTENDED DATA

**Extended data Fig. 1.**
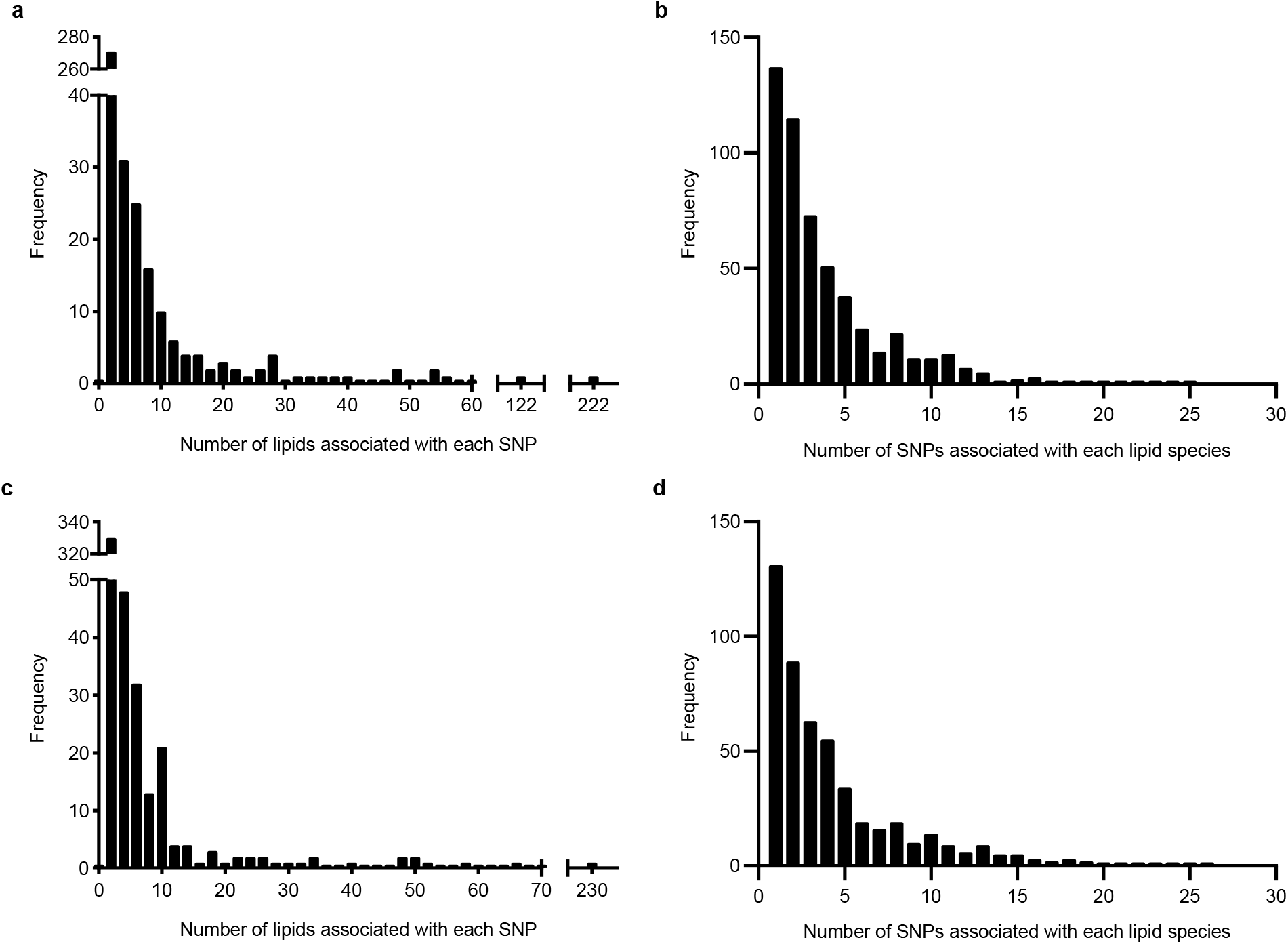
Distribution of genome-wide significant associations for independent SNPs and lipid species. **a**, the number of lipid species associated with independent SNPs in the BHS discovery cohort. **b**, the number of independent SNPs associated with each lipid species in the BHS discovery cohort. **c**, the number of lipid species associated with independent SNPs in the BHS discovery cohort following adjustment for clinical lipid traits. **d**, the number of independent SNPs associated with each lipid species in the BHS discovery cohort following adjustment for clinical lipid traits.

**Extended data Fig. 2.**
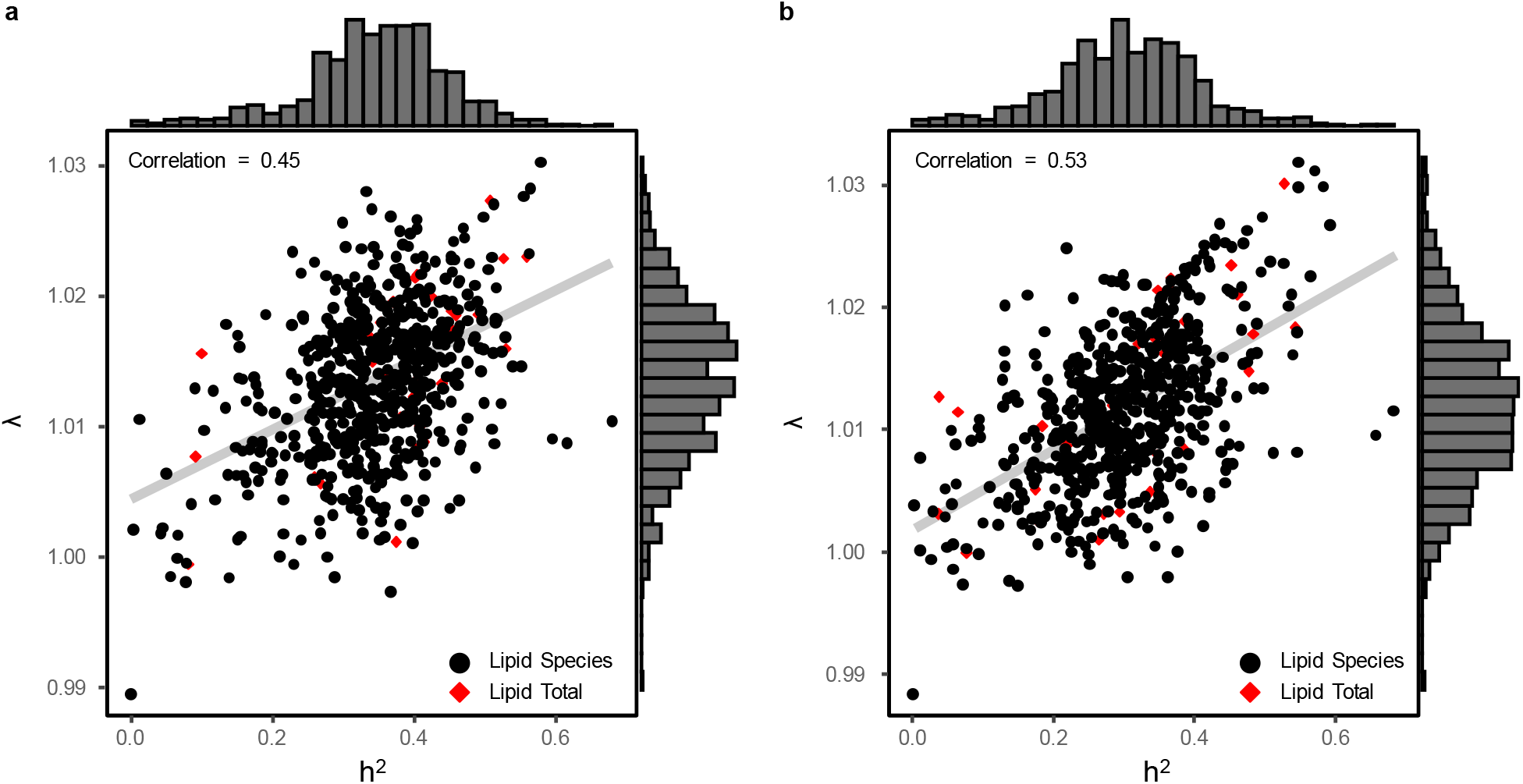
Scatterplot of lipid heritabilities (h^2^) vs GWAS genomic inflation factors (λ) for lipid species and classes. **a**, lipid heritability and genomic inflation factors for genome-wide association analysis in the BHS cohort. **b**, lipid heritability and genomic inflation factors for genome-wide association analysis, adjusting for clinical lipids, in the BHS cohort. Red diamonds indicate lipid classes and black circles indicate lipid species. The correlation between the heritabilities and genomic inflation factors are also shown, with a line of best fit. The right and top axes show histograms of the distribution of the genomic inflation factors from each GWAS, and heritability estimates, respectively. Heritability estimates were calculated in GCTA; using the genetic related matrix (GRM) and adjusted by age, sex, age2, age*sex, age2*sex.

**Extended data Fig. 3.**
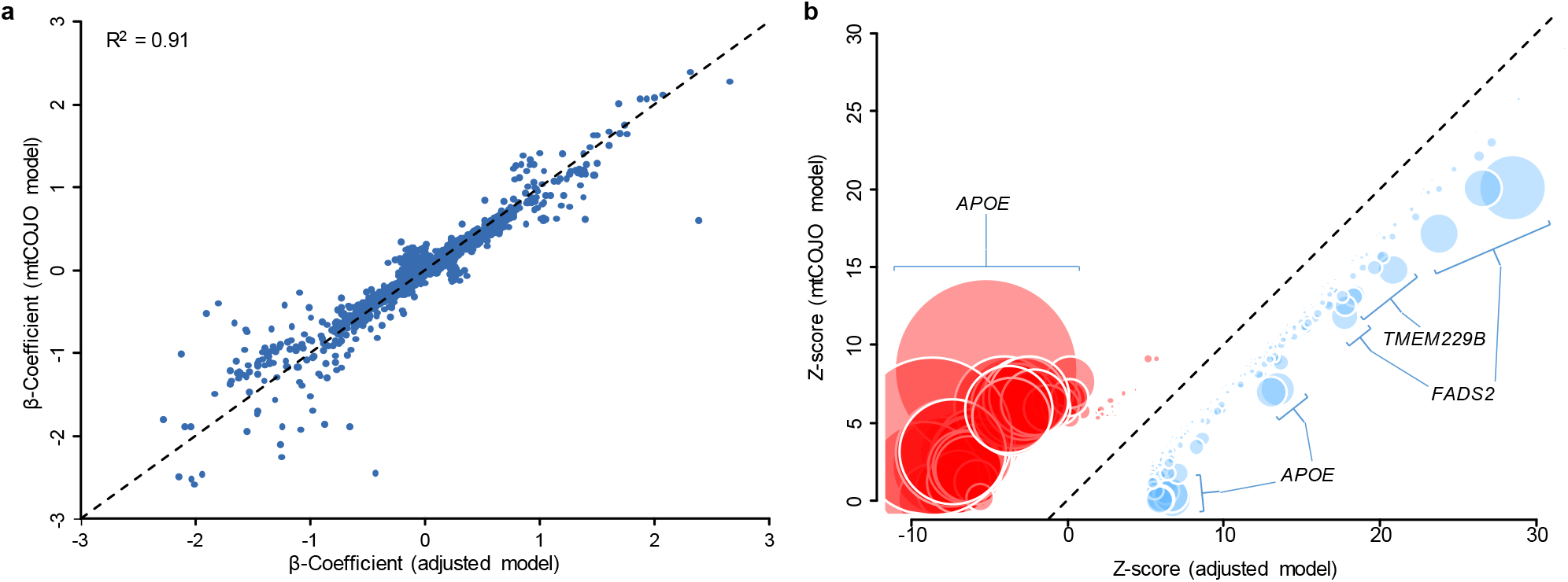
Comparison of estimated lipidomic effect sizes between clinical lipid adjusted and mtCOJO adjusted models. **a**, Beta coefficients for clinical lipid adjusted SNP-lipid associations (*x* axis) are plotted against mtCOJO adjusted SNP-lipid associations (*y* axis). **b**, Z-scores (Beta coefficient divided by standard error) for clinical lipid adjusted SNP-lipid associations (*x* axis) are plotted against mtCOJO adjusted SNP-lipid associations (*y* axis). Variant effect signs are fixed so mtCOJO adjusted associations are positive. Variants showed greater (positive) associations in mtCOJO adjusted analysis are shown in red, and variants showing reduced associations are shown in blue. Circle diameter is proportional of -log_10_(*P*) t-test of effect differences.

**Extended data Fig. 4.**
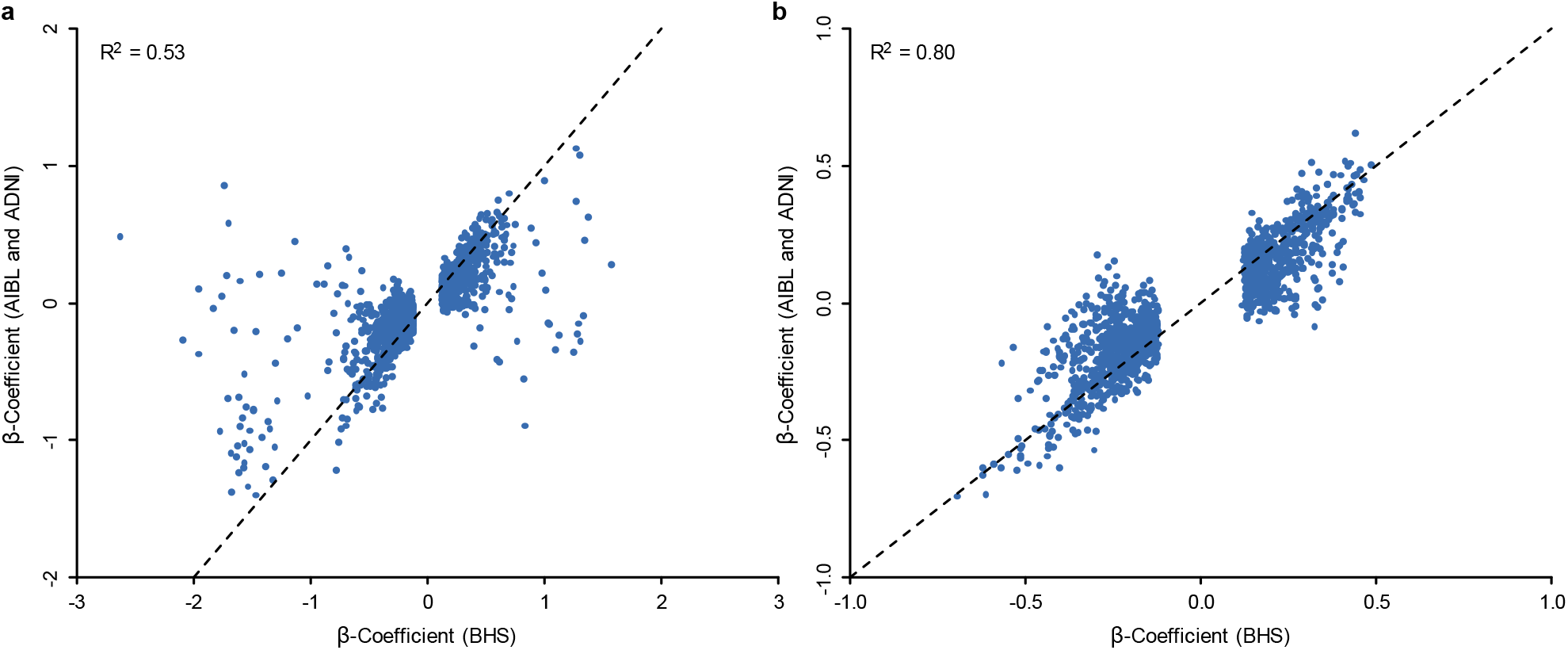
Comparison of estimated lipidomic effect sizes between the discovery BHS GWAS and the meta-analysis (ADNI and AIBL). **a**, Beta coefficients were estimated from linear regression models for lipid species using the Busselton Health Study discovery GWAS (*x*-axis) and the ADNI and AIBL validation meta-analysis (*y*-axis). **b**, Beta coefficients for only common SNPs (MAF>=0.05) in the Busselton Health Study discovery GWAS (*x*-axis) and the ADNI and AIBL validation meta-analysis (*y*-axis). Only significantly associated SNPs (P<5×10^−8^) in the Busselton Health Study discovery GWAS are shown.

