## Supplementary Information for "Comprehensive genetic analysis of the human lipidome identifies novel loci controlling lipid homeostasis with links to coronary artery disease"

### Table of Contents


| | | |
|---|---|---|
| Supplementary Note 1. | **Associations between 737 lead SNPs with lipid species and metabolites in previous studies.** | 2 |
| Supplementary Note 2. | **Regional association plots for lipid loci that colocalize with coronary artery disease.** | 4 |
| Supplementary Note 3. | **Comparison of estimated lipidomic effect sizes** | 48 |
| Supplementary References | | 49 |

**Supplementary Note 1: Associations between 737 lead SNPs with lipid species and metabolites in previous studies**. We identified 35 previous studies that had reported one or more of our 737 lead SNPs or a proxy ($R^2$>0.8) (Supplementary Table 13); details below.

| Author or database | PMID or DOI | Sample size | Method | # lipids/metabolites | Ethnicity | Reference |
|---|---|---|---|---|---|---|
| Burkhardt, R. | 26401656 | 2,107 | MS | 96 AA, AC | Central European | 1 |
| Chai, J.F. | 31628463 | 1,954 | MS/MS | 136 AA, AC | Chinese | 2 |
| Chasman, D.I. | 19936222 | 17,296 | NMR | 17 lipoproteins | European | 3 |
| Davis, J. | 29084231 | 8,372 | NMR | 68 lipid and lipoprotein subclasses | Finnish | 4 |
| Demirkan, A. | 22359512 | 4,034 | ESI-MS/MS | 153 lipids | European | 5 |
| Draisma, H. | 26068415 | 7,478 | ESI-FIA-MS/MS | 129 metabolites | European | 6 |
| Feofanova, E. | 29610217 | 1,552/1,872 | GC-MS, LC-MS | 102 circulating lipid-related metabolites | European, African-American | 7 |
| Gieger, C. | 19043545 | 284 | ESI-MS/MS | 363 metabolites | Southern German | 8 |
| Harshfield, E. | 10.1101/2020.10.16.20213520 | 5,662/13,814 | DI-HRMS | 360 lipids | Pakistani, European | 9 |
| Hartiala, J. | 26822151 | 1,985/1,895 | ESI-MS/MS | plasma betaine levels | European | 10 |
| Hicks, A.A. | 19798445 | 4,400 | ESI-MS/MS | 33 sphingolipids, and 43 matched metabolite ratios | European | 11 |
| Hong, M.G. | 23281178 | 402/489 | UHPLC-MS | 6,138 unique molecular features | Swedish | 12 |
| Hu, Y. | 28298293 | 3,521/12,020 | TLC-GC | Unclear number of Monounsaturated FA | Chinese, European | 13 |
| Illig, T. | 20037589 | 1,809/422 | HPLC-ESI-MS/MS | 163 metabolic traits | European | 14 |
| Inouye, M. | 22916037 | 6,600 | NMR | 130 metabolite measures | European | 15 |
| Kalsbeek, A. | 29652918 | 2,400 | GC | 22 FA and 15 FA ratios | Framingham Heart Study Offspring Cohort | 16 |
| Kettunen, J. | 22286219 | 8,330 | NMR | 216 serum metabolic traits | Finnish | 17 |
| Kettunen, J. | 27005778 | 24,925 | NMR | 123 metabolic traits | European | 18 |
| Krumsiek, J. | 23093944 | 1,768 | UHPLC-MS/MS, GC-MS/MS | 486 metabolic traits (including 213 unknowns) | German | 19 |
| Long, T. | 28263315 | 1,960 | UHPLC–MS/MS | 644 metabolites | European | 20 |
| Lotta, L. | 27898682 | 16,596 | MS/MS, GC-MS, UHPLC-MS/MS | Branched-chain AA | European | 21 |
| Lotta, L. | 33414548 | 8,569 - 86,507 | FIA-ESI-MS/MS, UHPLC-ESI-MS, NMR | 174 metabolites | European | 22 |
| Mittelstrass, K. | 21852955 | 3,381 | HPLC-MS/MS | 131 metabolites | South German | 23 |
| Nicholson, G. | 21931564 | 211 | NMR, FIA-ESI-MS/MS | 526 metabolite peaks | European | 24 |
| Raffler, J. | 26352407 | 3,861/1,691 | NMR | 15,379 targeted and untargeted metabolic traits | European | 25 |
| Rhee, E. | 23823483 | 2,076 | HPLC-ESI-MS/MS | 217 metabolites | European | 26 |
| Shin, S. | 24816252 | 7,824 | HPLC-MS/MS, GC-MS/MS | 486 metabolites (including 177 unknown) | European | 27 |
| SNIPA | 25431330 | - | - | - | Various | 28 |

| | | | | | | |
|---|---|---|---|---|---|---|
| Suhre, K. | 21886157 | 1,768/1,052 | UHPLC-MS/MS, GC-MS/MS | 276 metabolites | European | 29 |
| Tabassum, R. | 31551469 | 2,181 | DI-HRMS | 141 lipids | European | 30 |
| Teslovich, T. | 29481666 | 8,545 | NMR | 9 AA | Finnish | 31 |
| Tukiainen, T. | 22156771 | 8,330 | NMR | 117 metabolites | Finnish | 32 |
| Xie, W. | 23378610 | 957/341 | Unclear | 14 metabolites | European | 33 |
| Yet, I. | 27073872 | 1,001 | MS | 605 metabolites | European | 34 |
| Yu, B. | 27884205 | 1,872/1,552 | GC-MS, LC-MS | 70 AA | African American, European American | 35 |

Mass-spectrometry, MS; Tandem mass-spectrometry, MS/MS; High-resolution mass-spectrometry, HRMS; Nuclear magnetic resonance, NMR; Electrospray ionization, ESI; Gas chromatography, GC; (Ultra-)High performance liquid chromatography, (U)HPLC; Thin-layer chromatography, TLC; Flow-injection/infusion analysis, FIA; Direct infusion, DI; Amino acids, AA; Acyl-carnitines, AC; Fatty acids, FA;

**Supplementary Note 2: Regional association plots for lipid loci that colocalize with coronary artery disease.** We identified 47 putative shared causal variants from colocalization analysis of lipid loci (Supplementary Table 16). Shown is the regional association plot of coronary artery disease[36] and up to the top four lipid species that show evidence of colocalization (H3+H4 > 0.8; H4/H3 > 10). Regions were isolated from the 737 lead SNPs, with a window of 400 KB centred on the lead SNP. The indicated candidate SNP was the genetic variant with the largest posterior probability. Marker colours indicate linkage disequilibrium with the candidate SNP (obtained from the 1000 Genome European dataset). Colocalization sensitivity analysis was performed for the top colocalization, assessing the posterior probability of H0-H4 for different values of prior $p_{12}$. Shaded regions indicate the value of $p_{12}$ which produces posterior probabilities of H3+H4 > 0.8 and H4/H3 > 10.

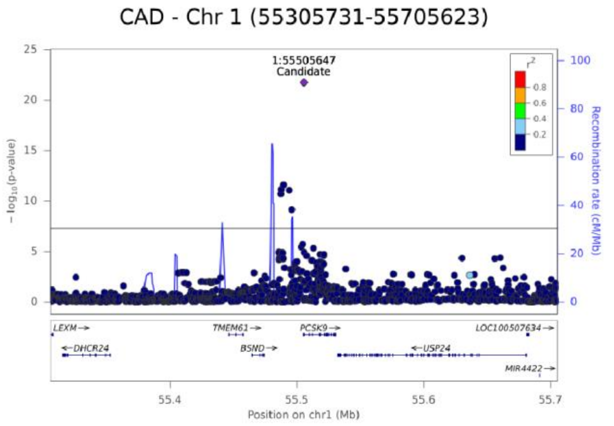
CAD - Chr 1 (55305731-55705623)
1:55505647
Candidate
- log10(p-value)
Recombination rate (cM/Mb)
r2
LEXM
DHCR24
TMEM61
BSND
PCSK9
USP24
LOC100507634
MIR4422
Position on chr1 (Mb)

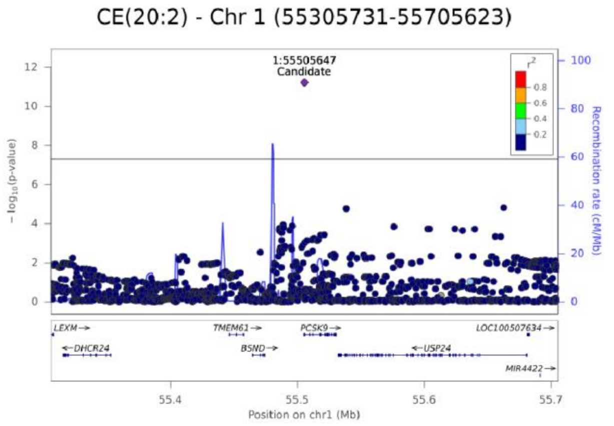
CE(20:2) - Chr 1 (55305731-55705623)
1:55505647
Candidate
- log10(p-value)
Recombination rate (cM/Mb)
r2
LEXM
DHCR24
TMEM61
BSND
PCSK9
USP24
LOC100507634
MIR4422
Position on chr1 (Mb)

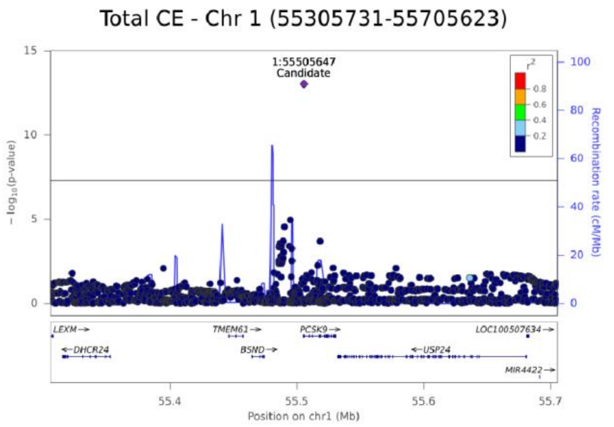
Total CE - Chr 1 (55305731-55705623)
1:55505647
Candidate
- log10(p-value)
Recombination rate (cM/Mb)
r2
LEXM
DHCR24
TMEM61
BSND
PCSK9
USP24
LOC100507634
MIR4422
Position on chr1 (Mb)

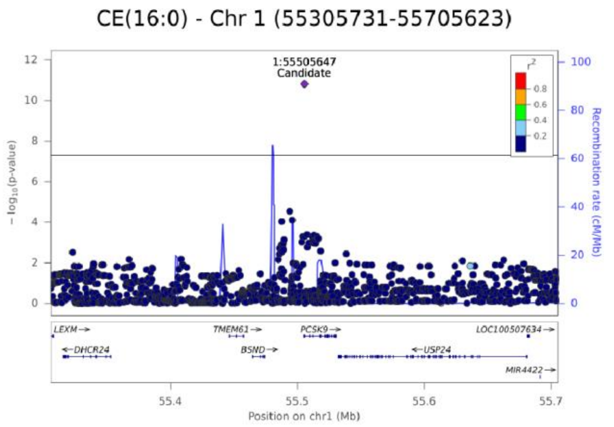
CE(16:0) - Chr 1 (55305731-55705623)
1:55505647
Candidate
- log10(p-value)
Recombination rate (cM/Mb)
r2
LEXM
DHCR24
TMEM61
BSND
PCSK9
USP24
LOC100507634
MIR4422
Position on chr1 (Mb)

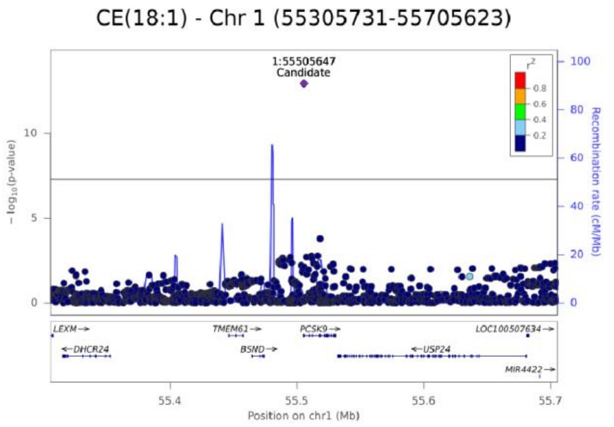
CE(18:1) - Chr 1 (55305731-55705623)
1:55505647
Candidate
- log10(p-value)
Recombination rate (cM/Mb)
r2
LEXM
DHCR24
TMEM61
BSND
PCSK9
USP24
LOC100507634
MIR4422
Position on chr1 (Mb)

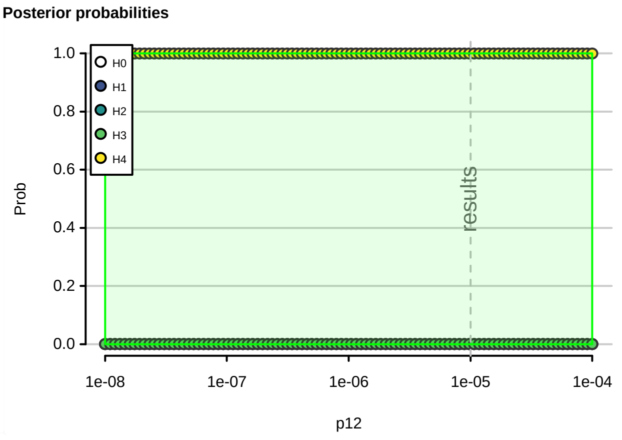
Posterior probabilities
Prob
H0
H1
H2
H3
H4
results
p12

CAD - Chr 1 (109621861-110020621)

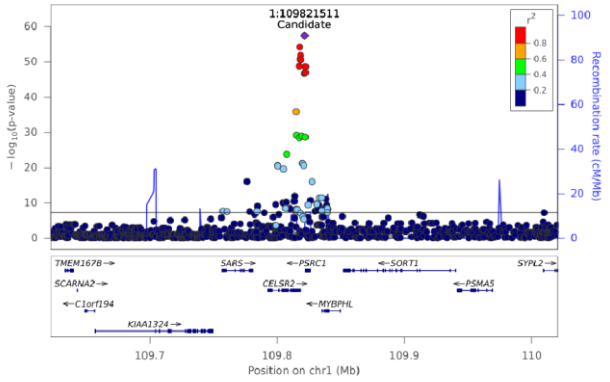


Posterior probabilities

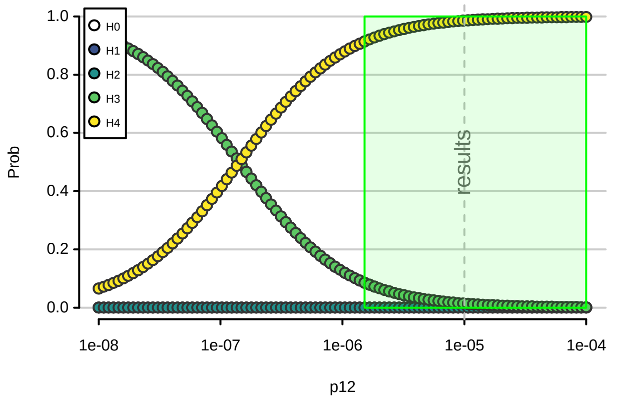


Total HexCer - Chr 1 (109621861-110020621)

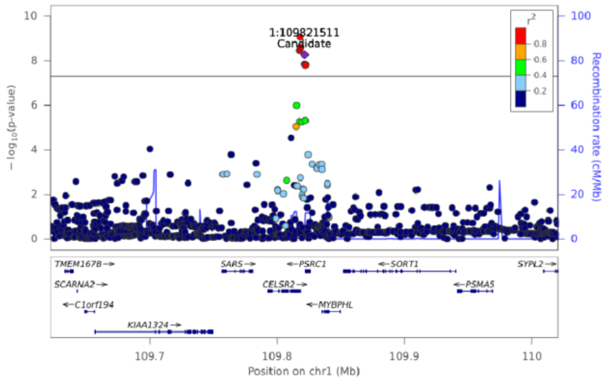


HexCer(d18:1/24:1) - Chr 1 (109621861-110020621)

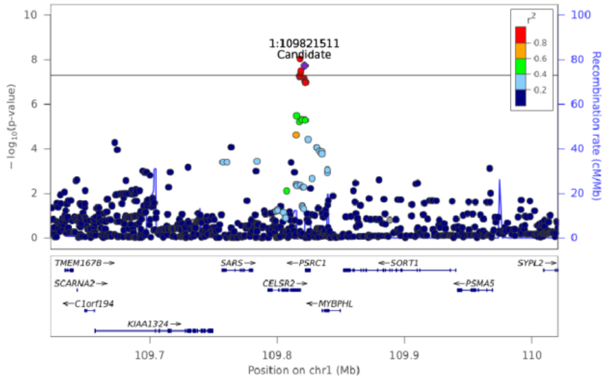

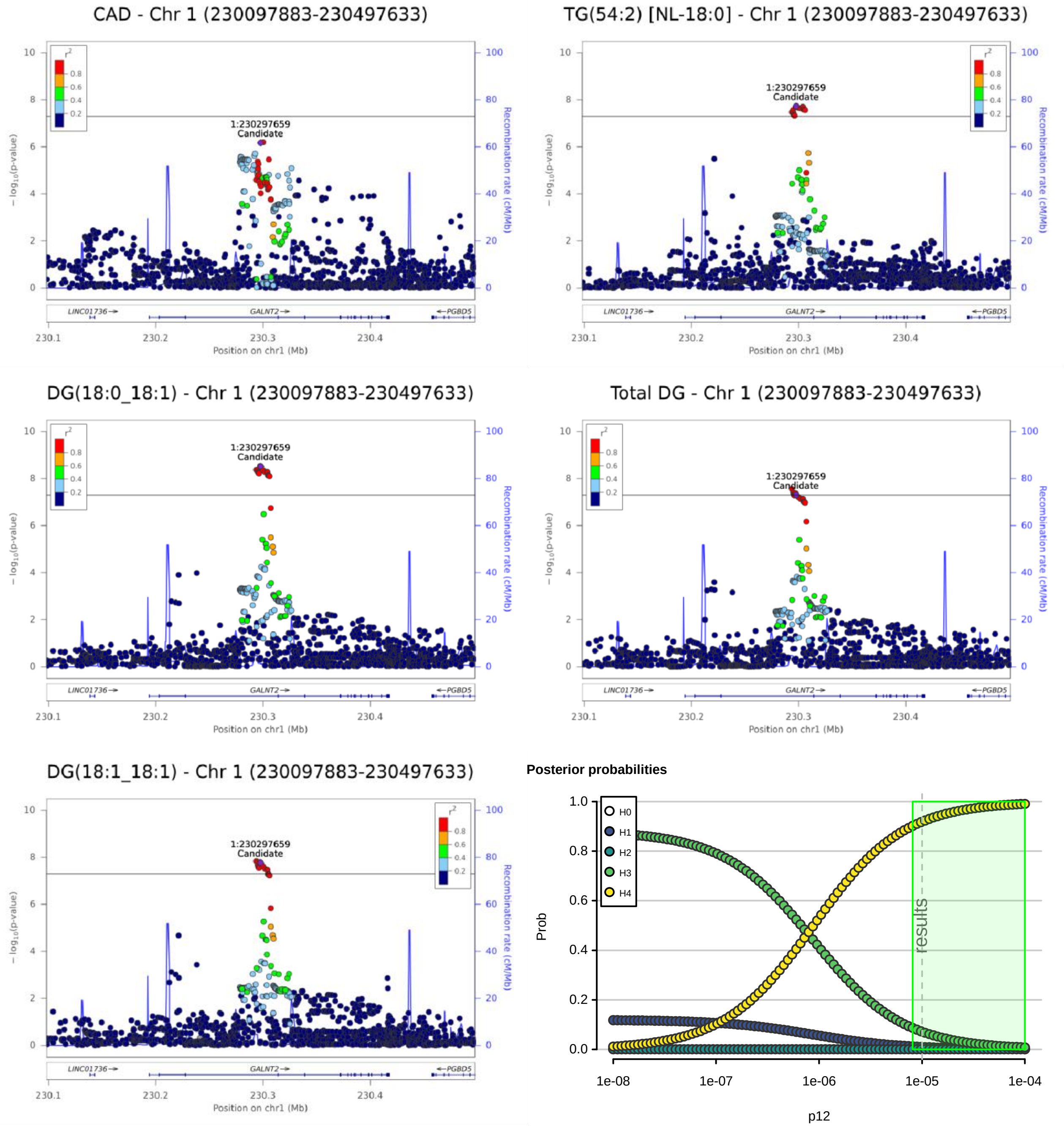
CAD - Chr 1 (230097883-230497633)
TG(54:2) [NL-18:0] - Chr 1 (230097883-230497633)
DG(18:0_18:1) - Chr 1 (230097883-230497633)
Total DG - Chr 1 (230097883-230497633)
DG(18:1_18:1) - Chr 1 (230097883-230497633)
1:230297659
Candidate
Recombination rate (cM/Mb)
LINC01736
GALNT2
PGBD5
Position on chr1 (Mb)
Posterior probabilities
H0
H1
H2
H3
H4
Prob
results
p12

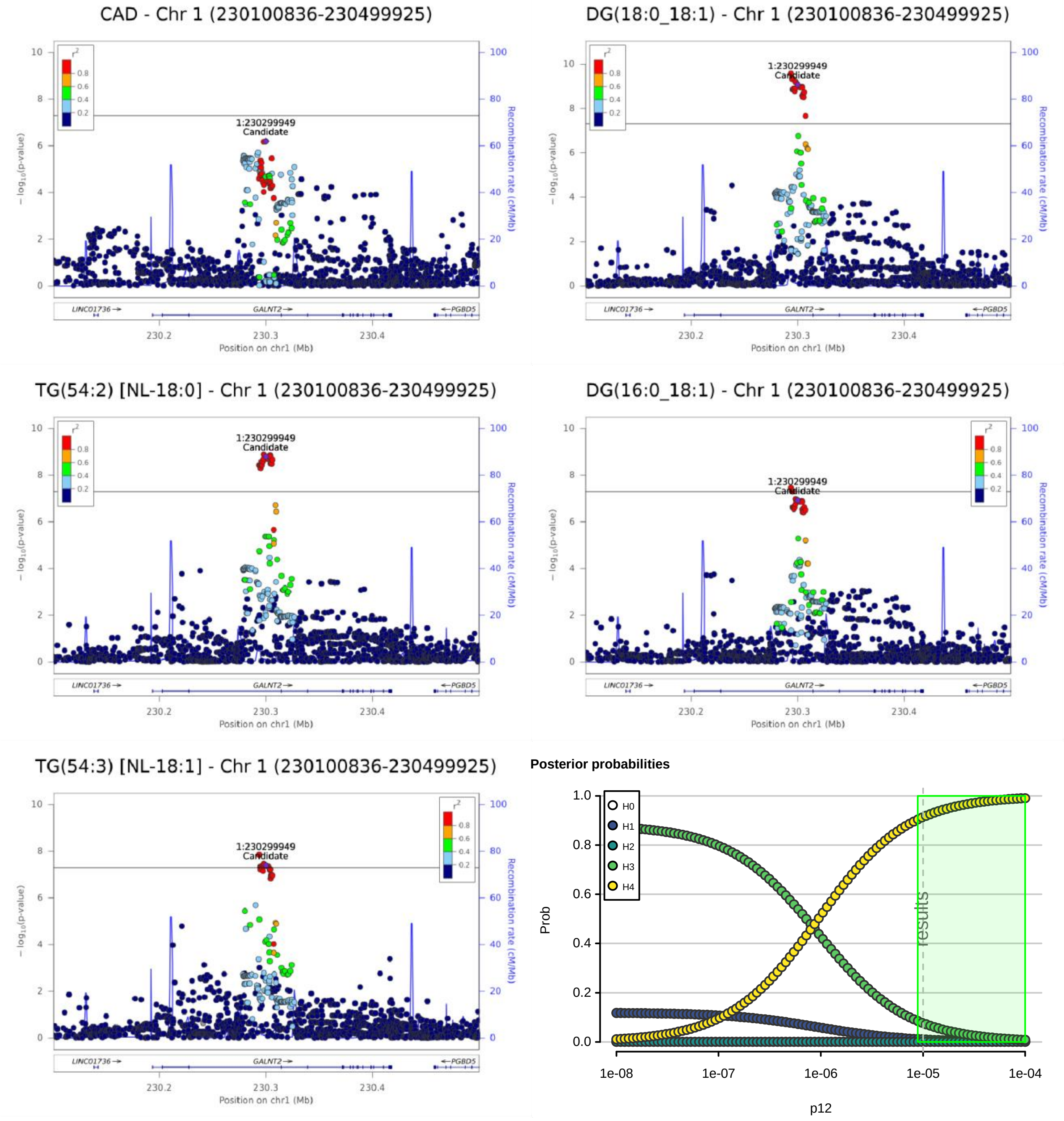
CAD - Chr 1 (230100836-230499925)
DG(18:0_18:1) - Chr 1 (230100836-230499925)
TG(54:2) [NL-18:0] - Chr 1 (230100836-230499925)
DG(16:0_18:1) - Chr 1 (230100836-230499925)
TG(54:3) [NL-18:1] - Chr 1 (230100836-230499925)
1:230299949
Candidate
r2
0.8
0.6
0.4
0.2
− log10(p-value)
Recombination rate (cM/Mb)
LINC01736→
GALNT2→
←PGBD5
230.2
230.3
230.4
Position on chr1 (Mb)
Posterior probabilities
H0
H1
H2
H3
H4
Prob
results
1e-08
1e-07
1e-06
1e-05
1e-04
p12

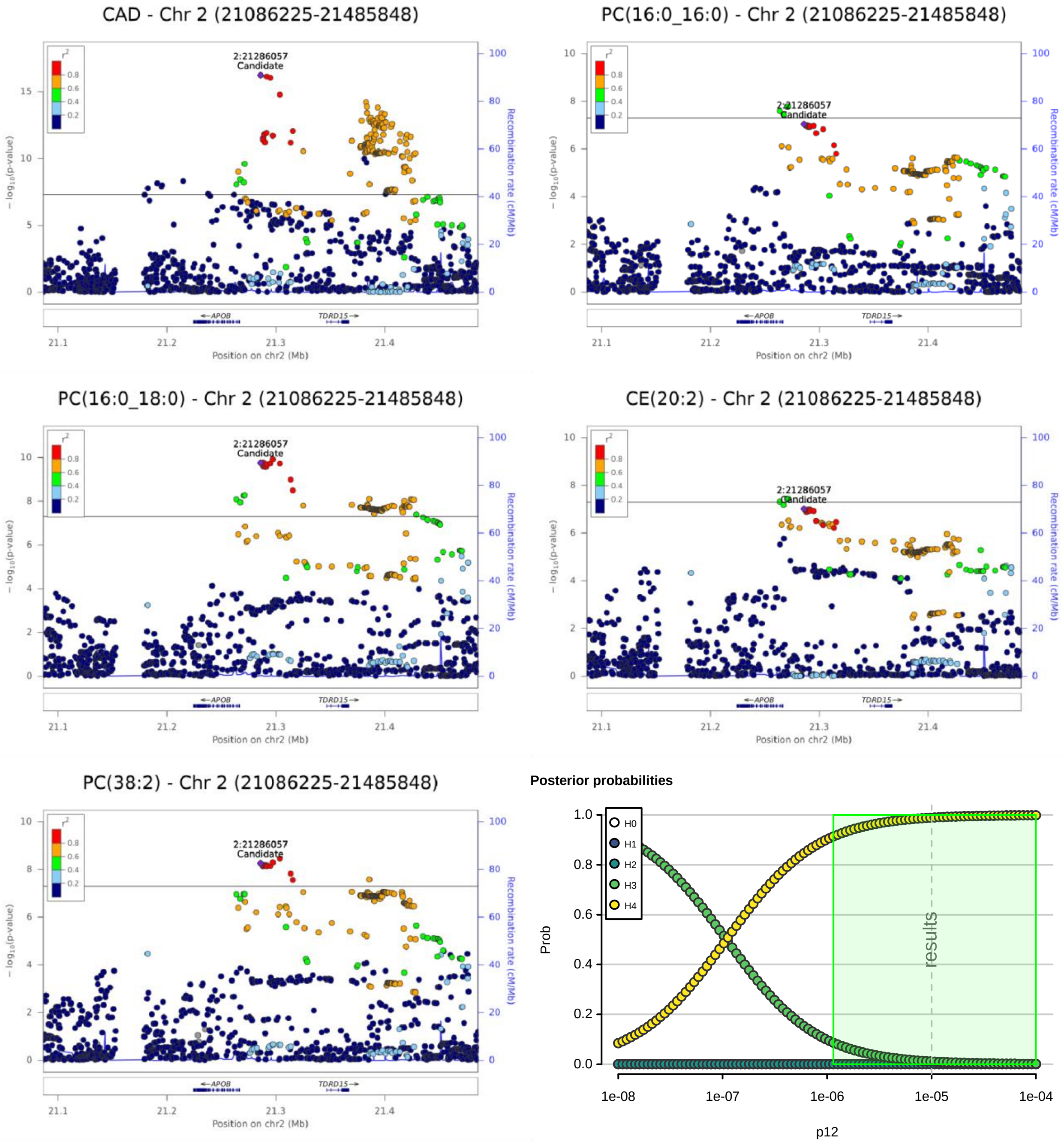
CAD - Chr 2 (21086225-21485848)
PC(16:0_16:0) - Chr 2 (21086225-21485848)
PC(16:0_18:0) - Chr 2 (21086225-21485848)
CE(20:2) - Chr 2 (21086225-21485848)
PC(38:2) - Chr 2 (21086225-21485848)
2:21286057
Candidate
$r^2$
0.8
0.6
0.4
0.2
$-\log_{10}$(p-value)
Recombination rate (cM/Mb)
APOB
TDRD15
21.1
21.2
21.3
21.4
Position on chr2 (Mb)
Posterior probabilities
Prob
H0
H1
H2
H3
H4
results
1e-08
1e-07
1e-06
1e-05
1e-04
p12

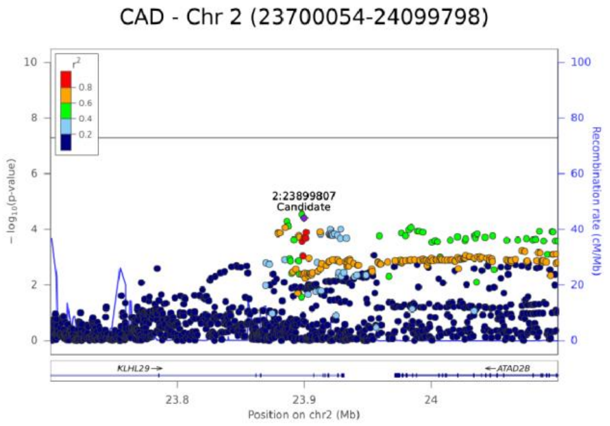
CAD - Chr 2 (23700054-24099798)
2:23899807
Candidate
− log10(p-value)
Recombination rate (cM/Mb)
KLHL29
ATAD2B
Position on chr2 (Mb)

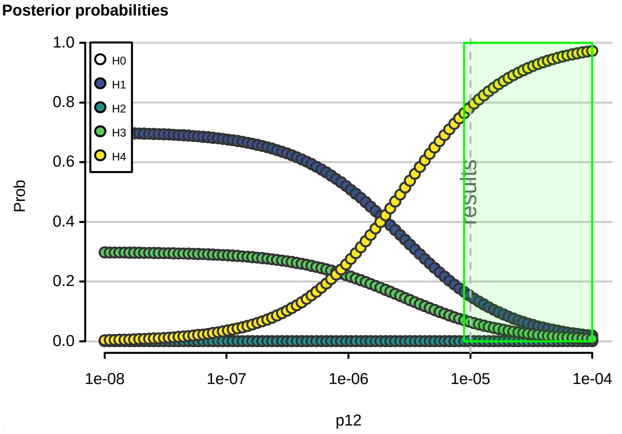
Posterior probabilities
H0
H1
H2
H3
H4
Prob
p12
results

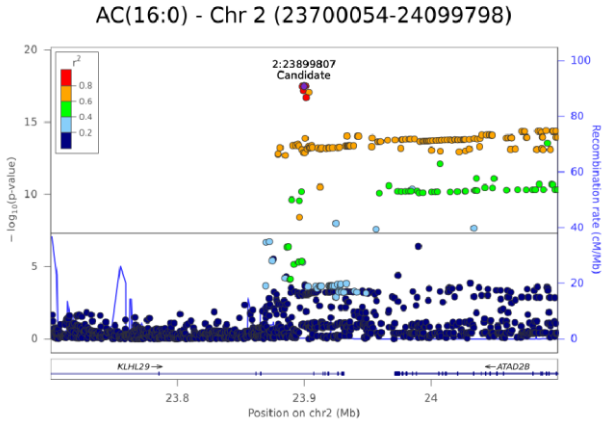
AC(16:0) - Chr 2 (23700054-24099798)
2:23899807
Candidate
− log10(p-value)
Recombination rate (cM/Mb)
KLHL29
ATAD2B
Position on chr2 (Mb)

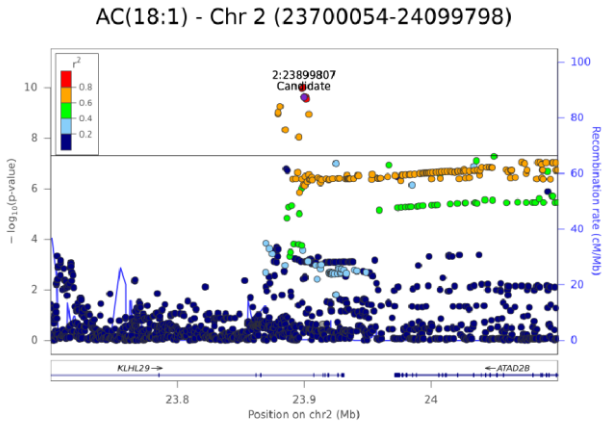
AC(18:1) - Chr 2 (23700054-24099798)
2:23899807
Candidate
− log10(p-value)
Recombination rate (cM/Mb)
KLHL29
ATAD2B
Position on chr2 (Mb)

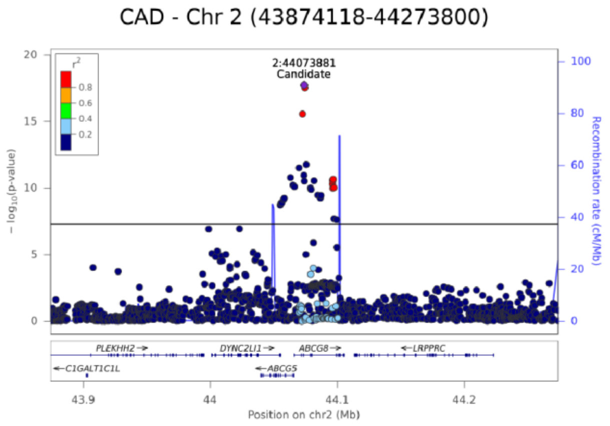
CAD - Chr 2 (43874118-44273800)
2:44073881
Candidate
− log10(p-value)
Recombination rate (cM/Mb)
PLEKHH2
DYNC2LI1
ABCG8
LRPPRC
C1GALT1C1L
ABCG5
Position on chr2 (Mb)

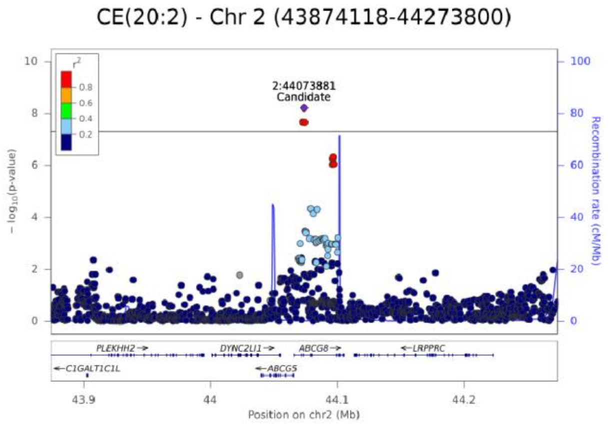
CE(20:2) - Chr 2 (43874118-44273800)
2:44073881
Candidate
− log10(p-value)
Recombination rate (cM/Mb)
PLEKHH2
DYNC2LI1
ABCG8
LRPPRC
C1GALT1C1L
ABCG5
Position on chr2 (Mb)

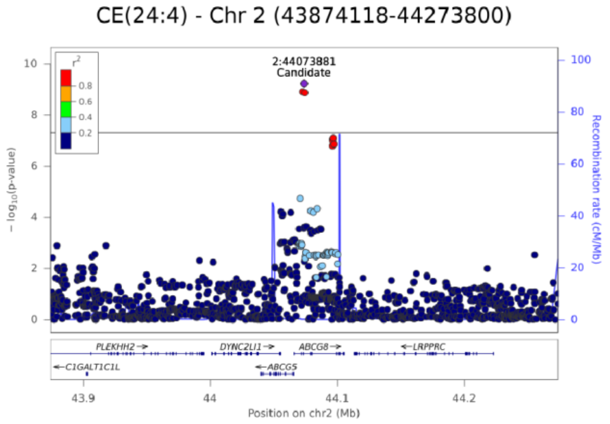
CE(24:4) - Chr 2 (43874118-44273800)
2:44073881
Candidate
− log10(p-value)
Recombination rate (cM/Mb)
PLEKHH2
DYNC2LI1
ABCG8
LRPPRC
C1GALT1C1L
ABCG5
Position on chr2 (Mb)

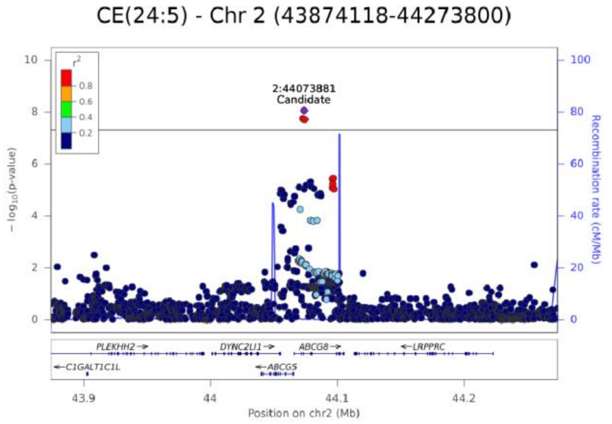
CE(24:5) - Chr 2 (43874118-44273800)
2:44073881
Candidate
− log10(p-value)
Recombination rate (cM/Mb)
PLEKHH2
DYNC2LI1
ABCG8
LRPPRC
C1GALT1C1L
ABCG5
Position on chr2 (Mb)

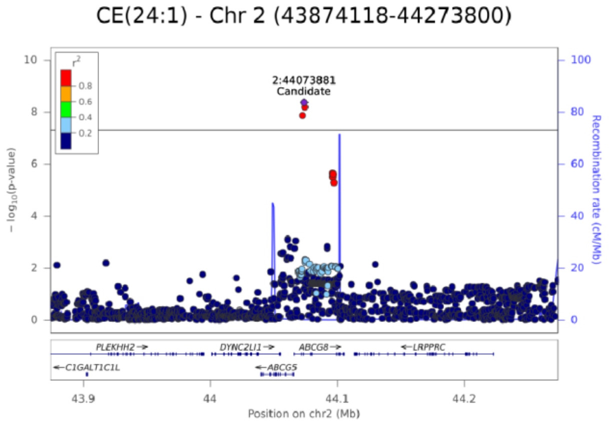
CE(24:1) - Chr 2 (43874118-44273800)
2:44073881
Candidate
− log10(p-value)
Recombination rate (cM/Mb)
PLEKHH2
DYNC2LI1
ABCG8
LRPPRC
C1GALT1C1L
ABCG5
Position on chr2 (Mb)

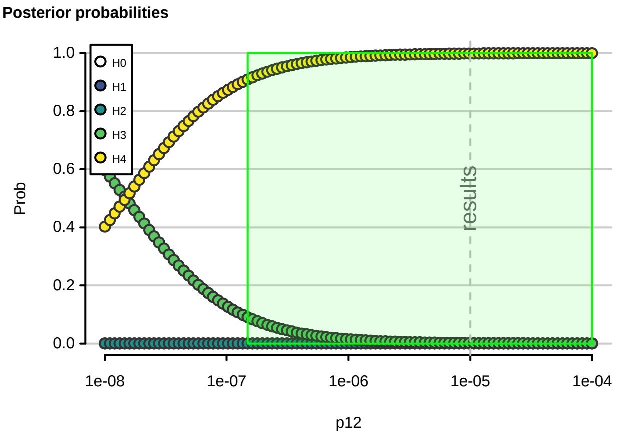
Posterior probabilities
H0
H1
H2
H3
H4
Prob
results
p12

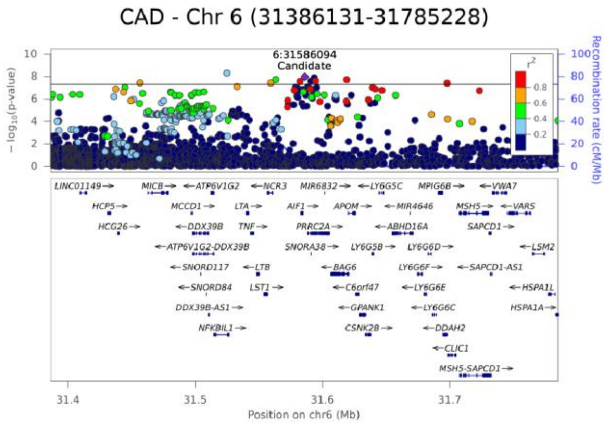
CAD - Chr 6 (31386131-31785228)
6:31586094
Candidate
− log10(p-value)
Recombination rate (cM/Mb)
Position on chr6 (Mb)

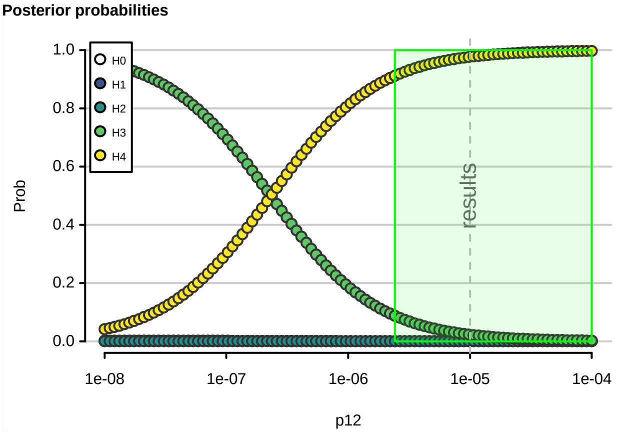
Posterior probabilities
H0
H1
H2
H3
H4
Prob
results
p12

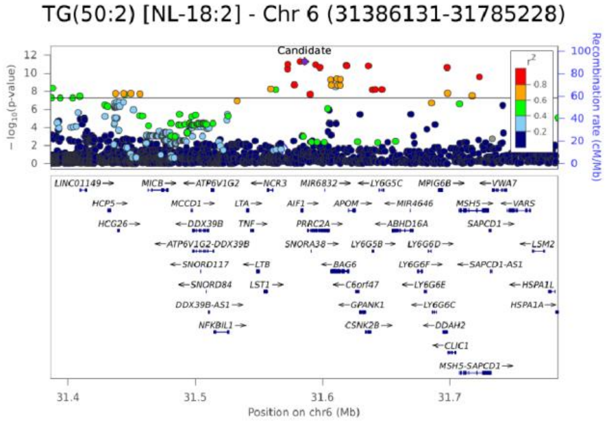
TG(50:2) [NL-18:2] - Chr 6 (31386131-31785228)
Candidate
− log10(p-value)
Recombination rate (cM/Mb)
Position on chr6 (Mb)

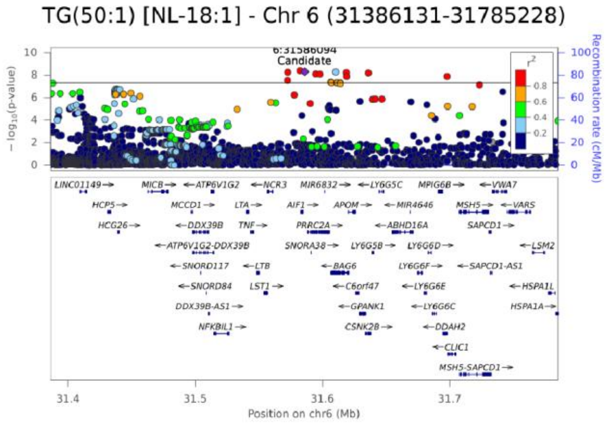
TG(50:1) [NL-18:1] - Chr 6 (31386131-31785228)
6:31586094
Candidate
− log10(p-value)
Recombination rate (cM/Mb)
Position on chr6 (Mb)

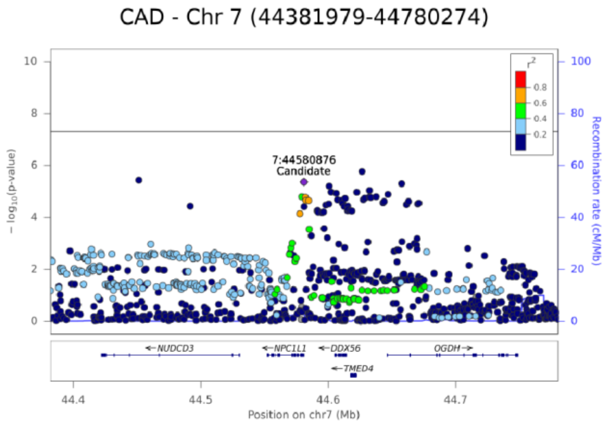


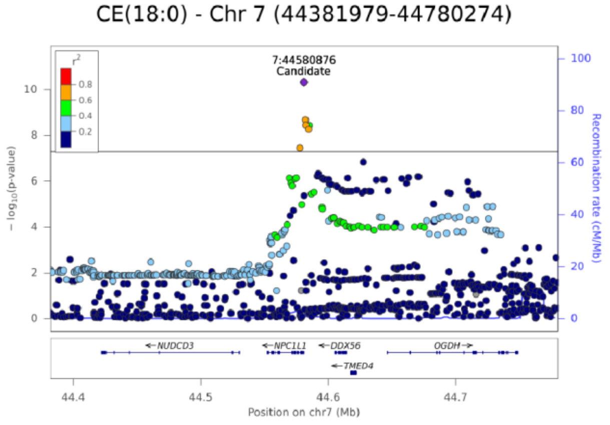


**Posterior probabilities**

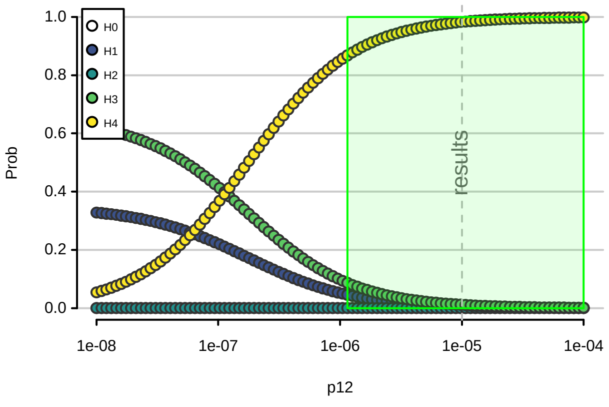

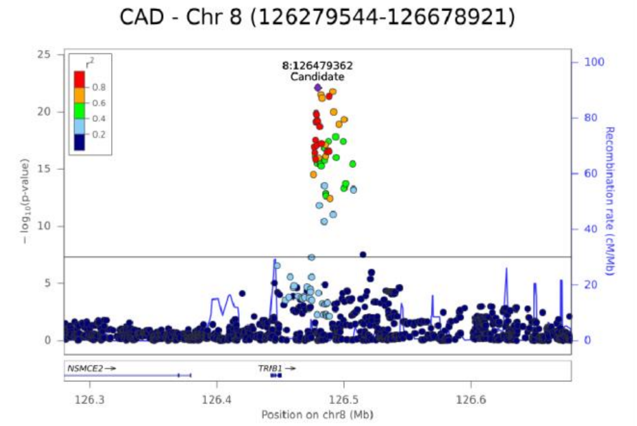


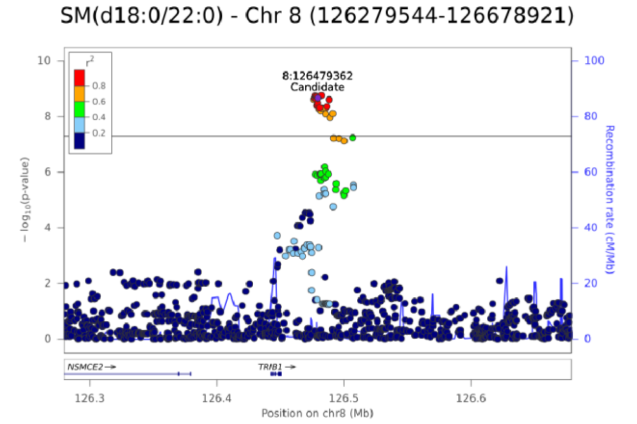


**Posterior probabilities**

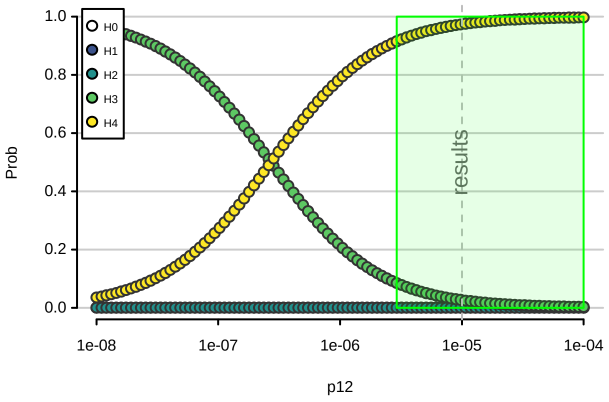

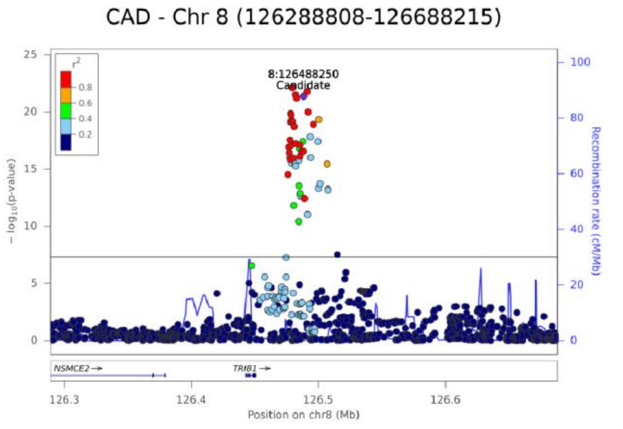


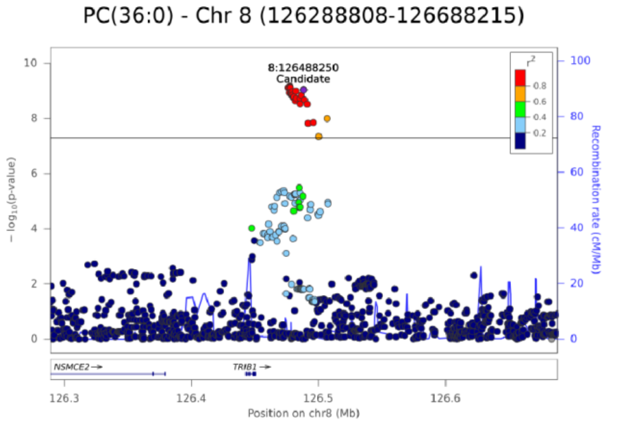


**Posterior probabilities**

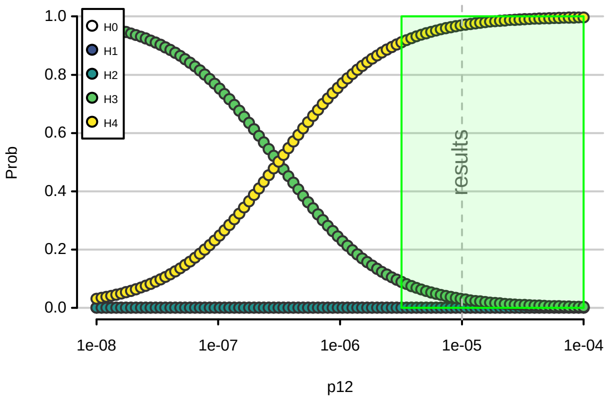

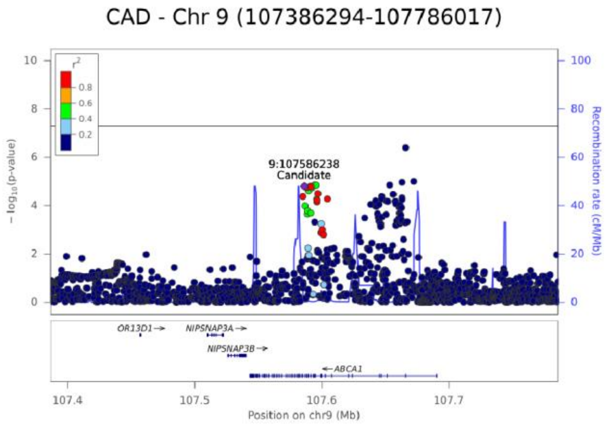
CAD - Chr 9 (107386294-107786017)
9:107586238
Candidate
− log10(p-value)
Recombination rate (cM/Mb)
OR13D1
NIPSNAP3A
NIPSNAP3B
ABCA1
Position on chr9 (Mb)

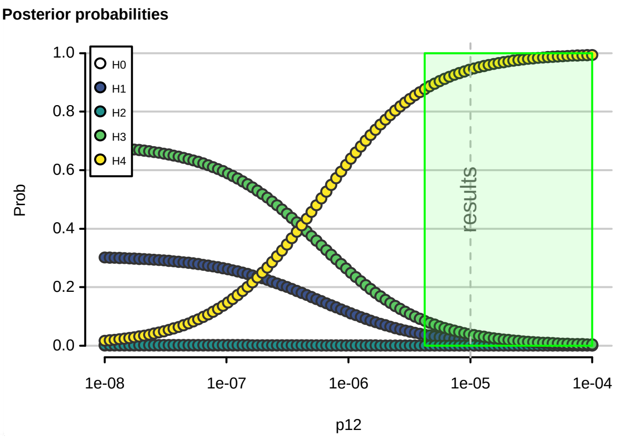
Posterior probabilities
H0
H1
H2
H3
H4
Prob
results
p12

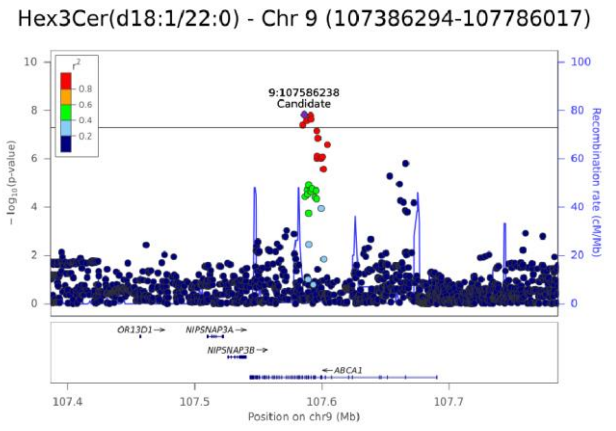
Hex3Cer(d18:1/22:0) - Chr 9 (107386294-107786017)
9:107586238
Candidate
− log10(p-value)
Recombination rate (cM/Mb)
OR13D1
NIPSNAP3A
NIPSNAP3B
ABCA1
Position on chr9 (Mb)

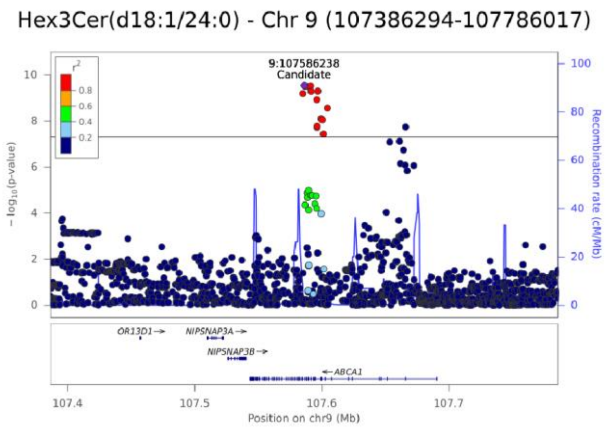
Hex3Cer(d18:1/24:0) - Chr 9 (107386294-107786017)
9:107586238
Candidate
− log10(p-value)
Recombination rate (cM/Mb)
OR13D1
NIPSNAP3A
NIPSNAP3B
ABCA1
Position on chr9 (Mb)

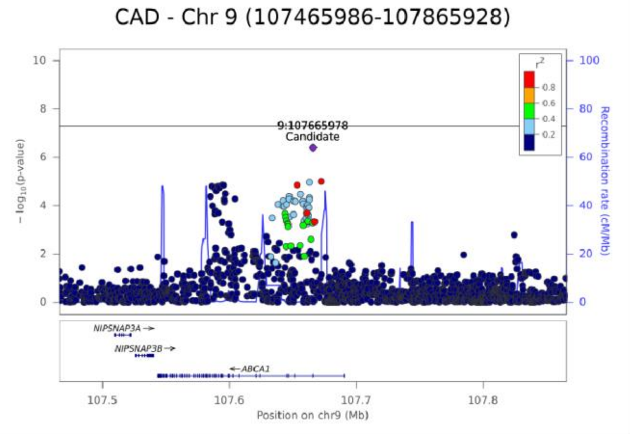


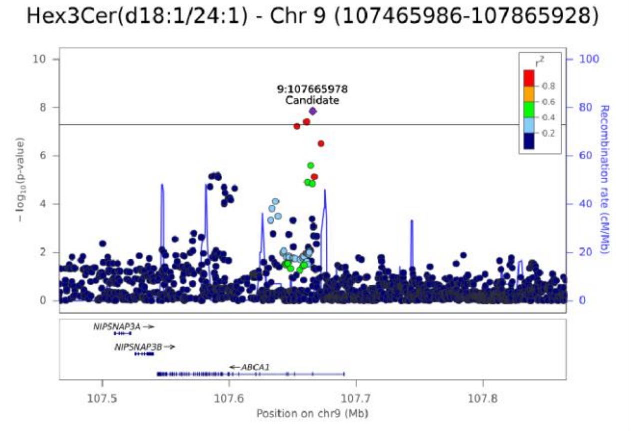


**Posterior probabilities**

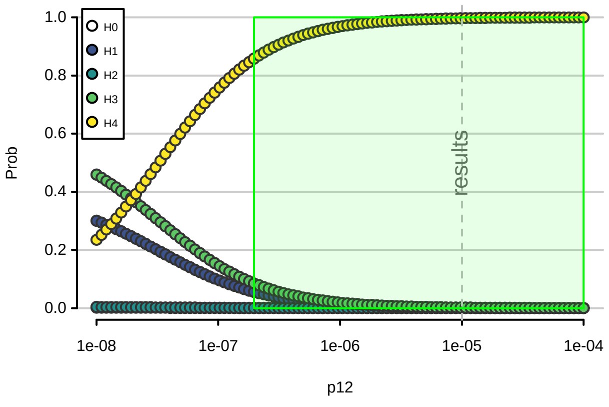

CAD - Chr 9 (135941932-136341776)

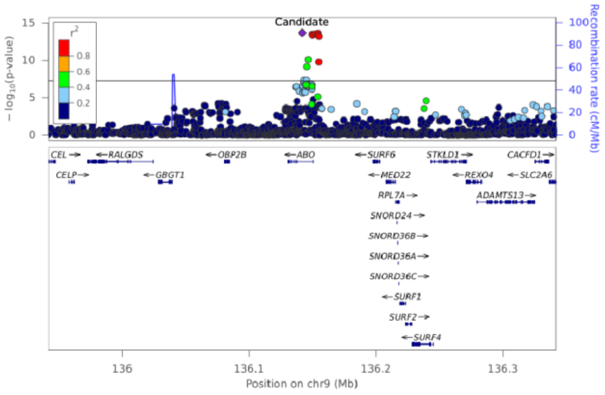


CE(18:0) - Chr 9 (135941932-136341776)

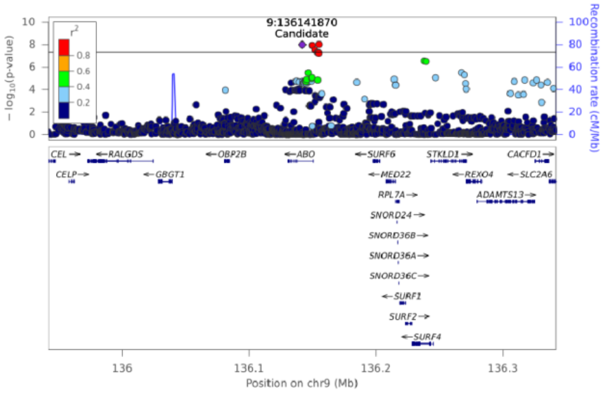


**Posterior probabilities**

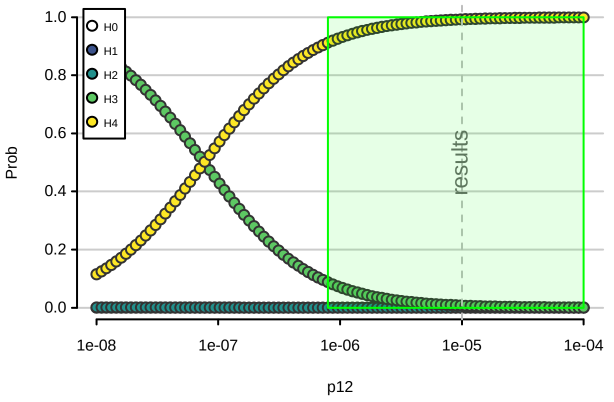

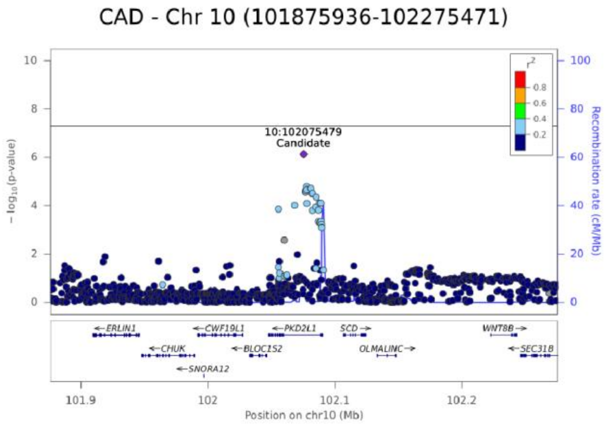
CAD - Chr 10 (101875936-102275471)
10:102075479
Candidate
−log10(p-value)
Recombination rate (cM/Mb)
ERLIN1
CWF19L1
PKD2L1
SCD
WNT8B
CHUK
BLOC1S2
OLMALINC
SEC31B
SNORA12
Position on chr10 (Mb)

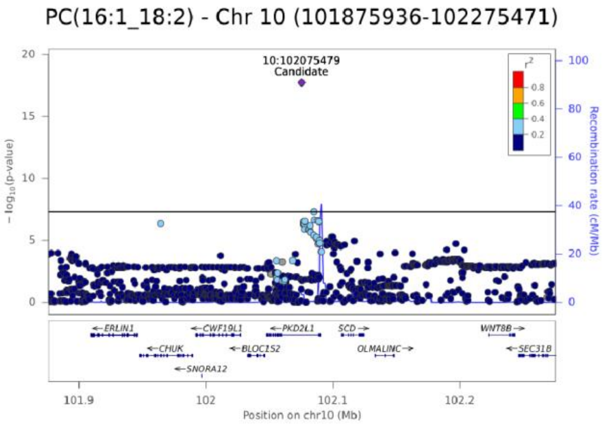
PC(16:1_18:2) - Chr 10 (101875936-102275471)
10:102075479
Candidate
−log10(p-value)
Recombination rate (cM/Mb)
Position on chr10 (Mb)

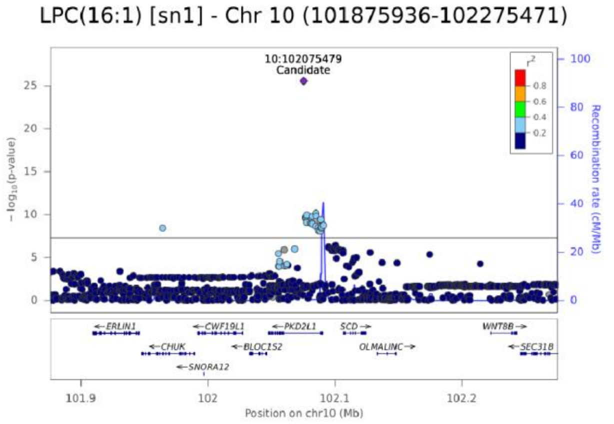
LPC(16:1) [sn1] - Chr 10 (101875936-102275471)
10:102075479
Candidate
−log10(p-value)
Recombination rate (cM/Mb)
Position on chr10 (Mb)

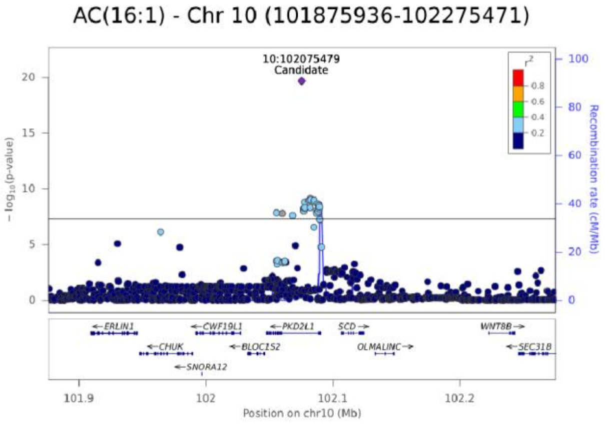
AC(16:1) - Chr 10 (101875936-102275471)
10:102075479
Candidate
−log10(p-value)
Recombination rate (cM/Mb)
Position on chr10 (Mb)

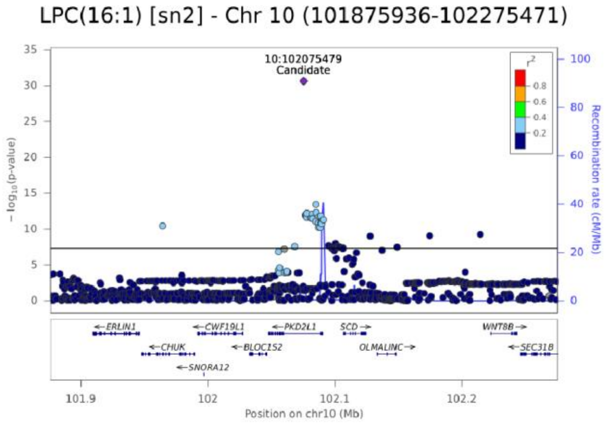
LPC(16:1) [sn2] - Chr 10 (101875936-102275471)
10:102075479
Candidate
−log10(p-value)
Recombination rate (cM/Mb)
Position on chr10 (Mb)

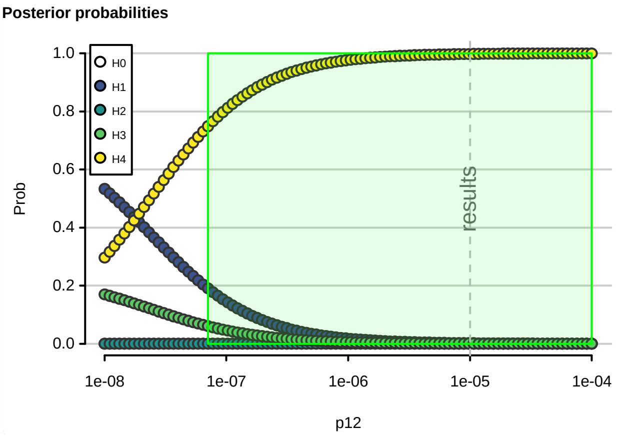
Posterior probabilities
H0
H1
H2
H3
H4
Prob
results
p12

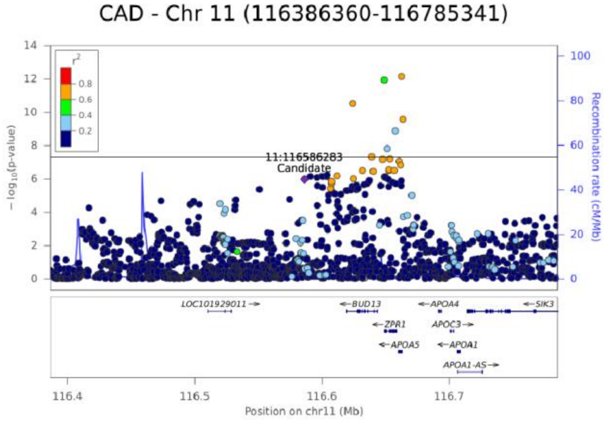

CAD - Chr 11 (116386360-116785341)
11:116586283
Candidate
LOC101929011
BUD13
ZPR1
APOA5
APOA4
APOC3
APOA1
APOA1-AS
SIK3
Position on chr11 (Mb)
Recombination rate (cM/Mb)


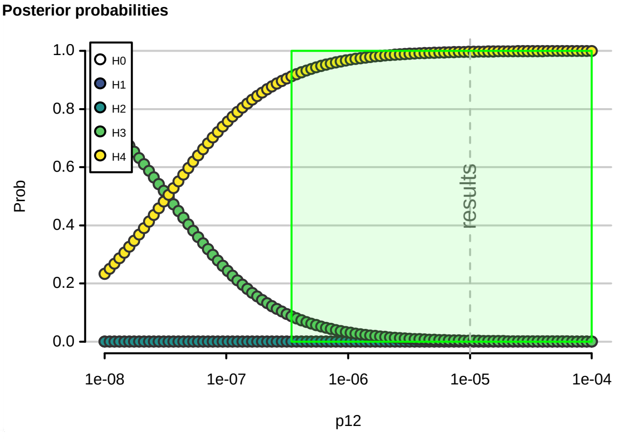

Posterior probabilities
H0
H1
H2
H3
H4
Prob
results
p12


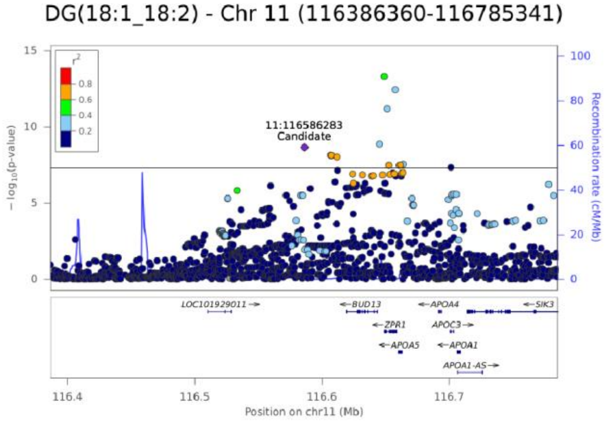

DG(18:1_18:2) - Chr 11 (116386360-116785341)
11:116586283
Candidate
Position on chr11 (Mb)
Recombination rate (cM/Mb)


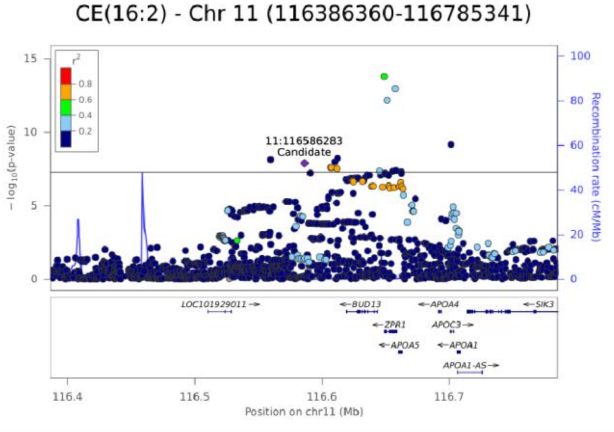

CE(16:2) - Chr 11 (116386360-116785341)
11:116586283
Candidate
Position on chr11 (Mb)
Recombination rate (cM/Mb)

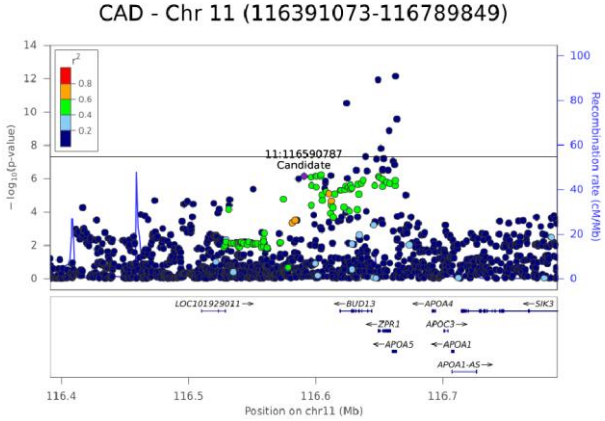
CAD - Chr 11 (116391073-116789849)
11:116590787
Candidate
− log10(p-value)
Recombination rate (cM/Mb)
LOC101929011
BUD13
ZPR1
APOA5
APOA4
APOC3
APOA1
APOA1-AS
SIK3
Position on chr11 (Mb)

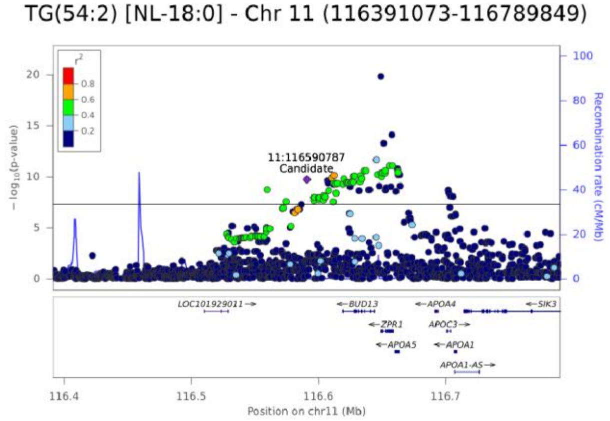
TG(54:2) [NL-18:0] - Chr 11 (116391073-116789849)
11:116590787
Candidate
− log10(p-value)
Recombination rate (cM/Mb)
Position on chr11 (Mb)

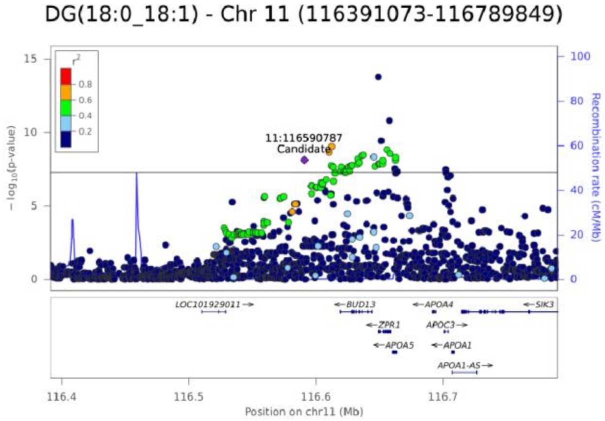
DG(18:0_18:1) - Chr 11 (116391073-116789849)
11:116590787
Candidate
− log10(p-value)
Recombination rate (cM/Mb)
Position on chr11 (Mb)

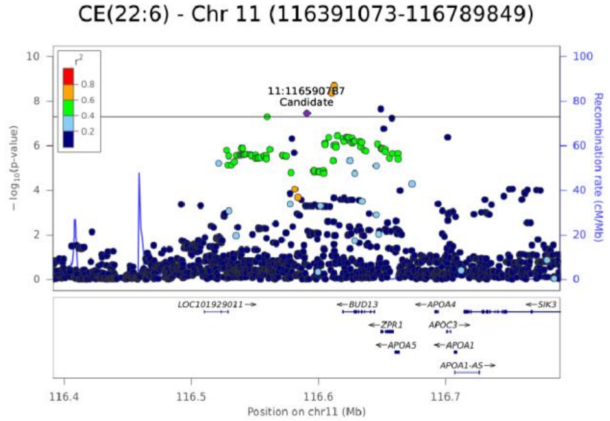
CE(22:6) - Chr 11 (116391073-116789849)
11:116590787
Candidate
− log10(p-value)
Recombination rate (cM/Mb)
Position on chr11 (Mb)

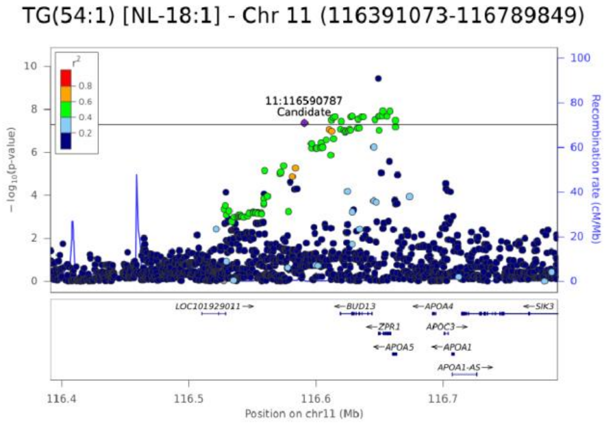
TG(54:1) [NL-18:1] - Chr 11 (116391073-116789849)
11:116590787
Candidate
− log10(p-value)
Recombination rate (cM/Mb)
Position on chr11 (Mb)

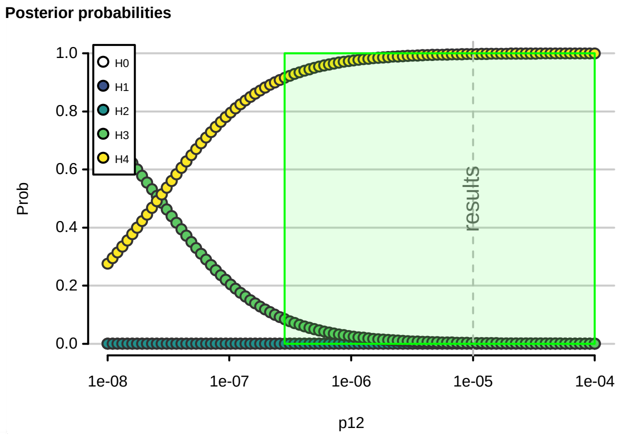
Posterior probabilities
H0
H1
H2
H3
H4
Prob
results
p12

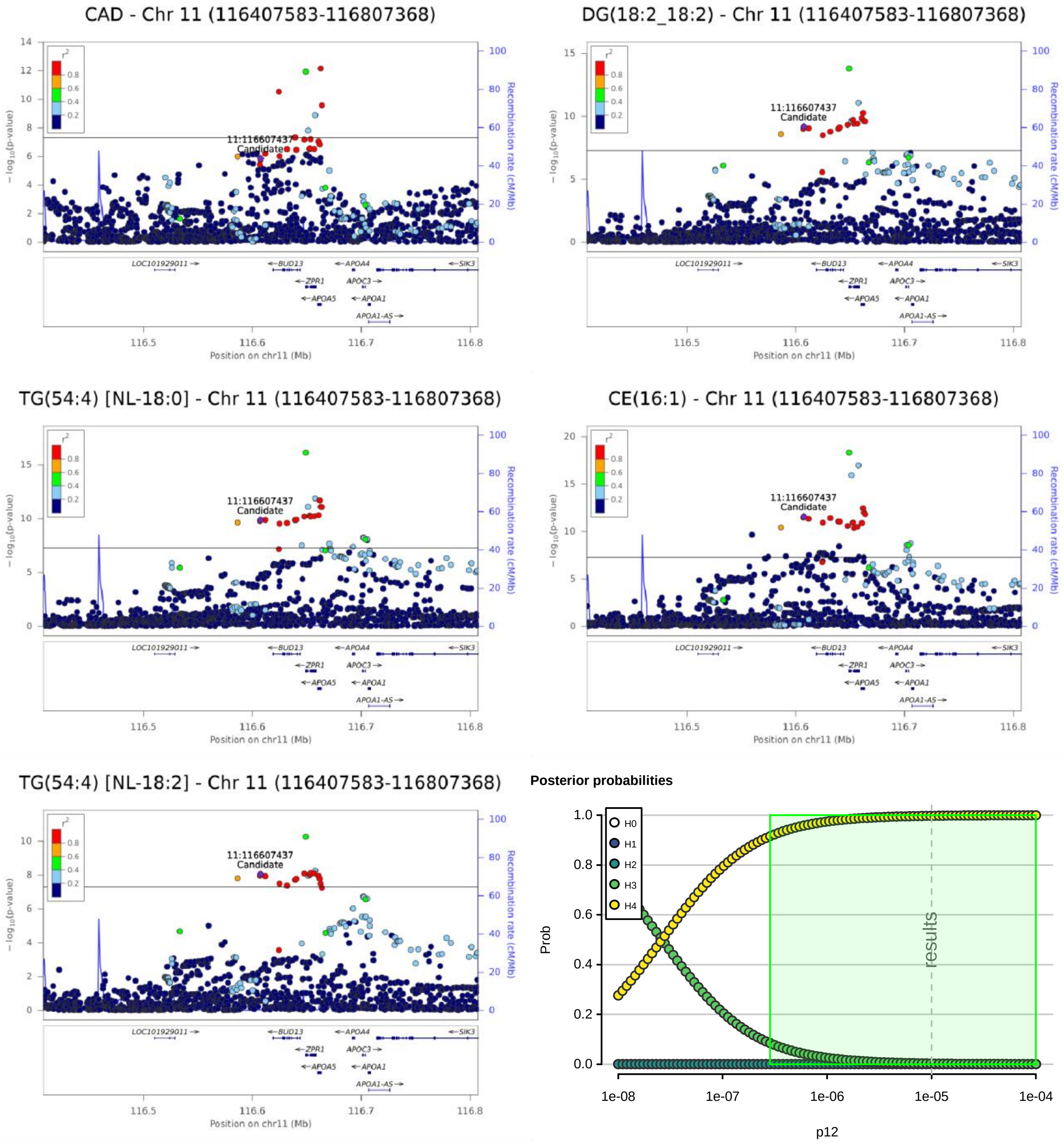
CAD - Chr 11 (116407583-116807368)
DG(18:2_18:2) - Chr 11 (116407583-116807368)
TG(54:4) [NL-18:0] - Chr 11 (116407583-116807368)
CE(16:1) - Chr 11 (116407583-116807368)
TG(54:4) [NL-18:2] - Chr 11 (116407583-116807368)
11:116607437
Candidate
Recombination rate (cM/Mb)
Position on chr11 (Mb)
LOC101929011
BUD13
ZPR1
APOA5
APOA4
APOC3
APOA1
APOA1-AS
SIK3
Posterior probabilities
Prob
p12
H0
H1
H2
H3
H4
results

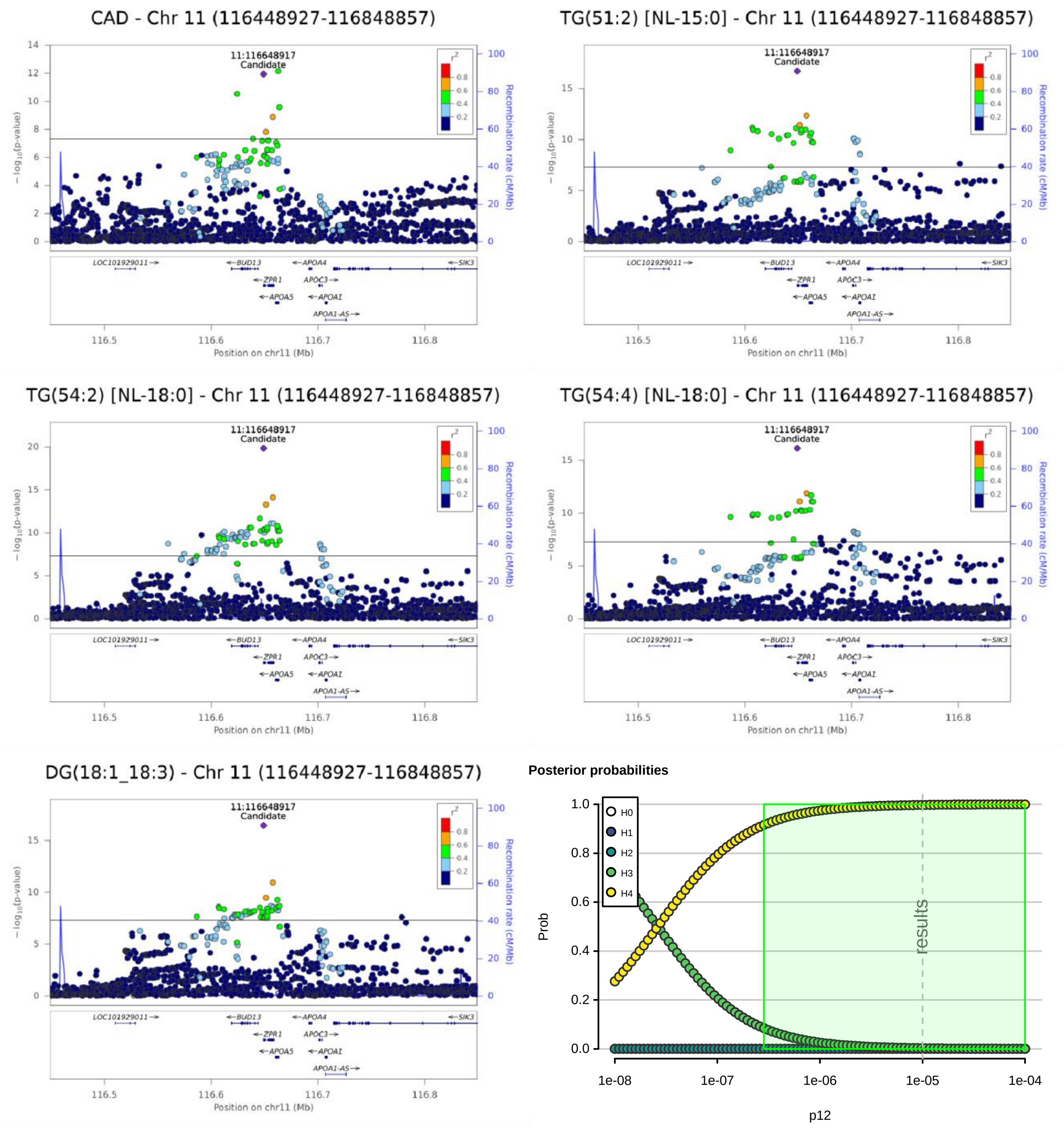
CAD - Chr 11 (116448927-116848857)
TG(51:2) [NL-15:0] - Chr 11 (116448927-116848857)
TG(54:2) [NL-18:0] - Chr 11 (116448927-116848857)
TG(54:4) [NL-18:0] - Chr 11 (116448927-116848857)
DG(18:1_18:3) - Chr 11 (116448927-116848857)
11:116648917
Candidate
Recombination rate (cM/Mb)
Position on chr11 (Mb)
LOC101929011
BUD13
ZPR1
APOA5
APOA4
APOC3
APOA1
APOA1-AS
SIK3
Posterior probabilities
H0
H1
H2
H3
H4
Prob
results
p12

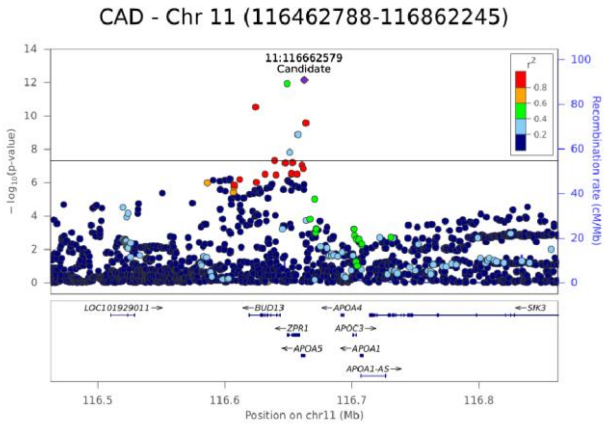
CAD - Chr 11 (116462788-116862245)
11:116662579
Candidate
− log10(p-value)
Recombination rate (cM/Mb)
LOC101929011
BUD13
ZPR1
APOA5
APOA4
APOC3
APOA1
APOA1-AS
SIK3
Position on chr11 (Mb)

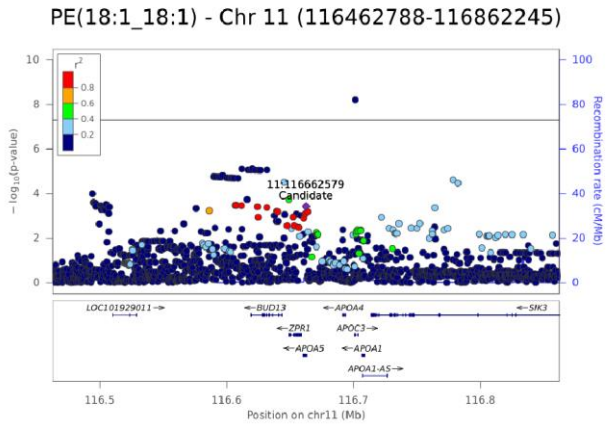
PE(18:1_18:1) - Chr 11 (116462788-116862245)
11:116662579
Candidate
− log10(p-value)
Recombination rate (cM/Mb)
LOC101929011
BUD13
ZPR1
APOA5
APOA4
APOC3
APOA1
APOA1-AS
SIK3
Position on chr11 (Mb)

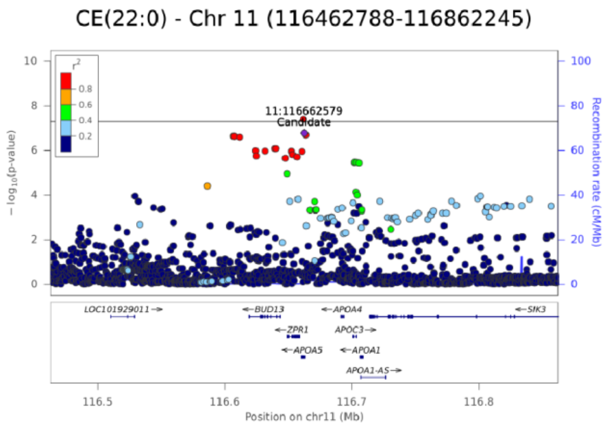
CE(22:0) - Chr 11 (116462788-116862245)
11:116662579
Candidate
− log10(p-value)
Recombination rate (cM/Mb)
LOC101929011
BUD13
ZPR1
APOA5
APOA4
APOC3
APOA1
APOA1-AS
SIK3
Position on chr11 (Mb)

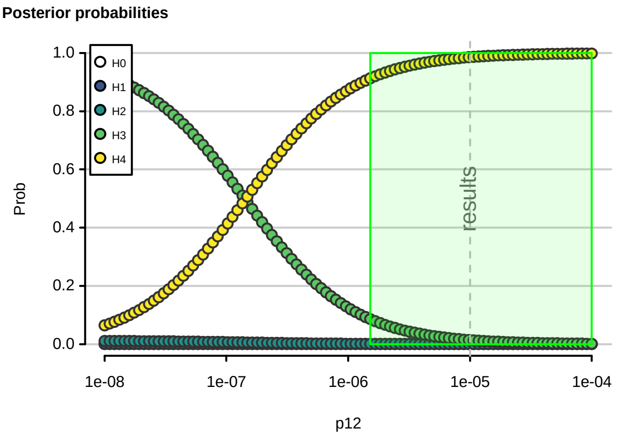
Posterior probabilities
Prob
H0
H1
H2
H3
H4
results
p12

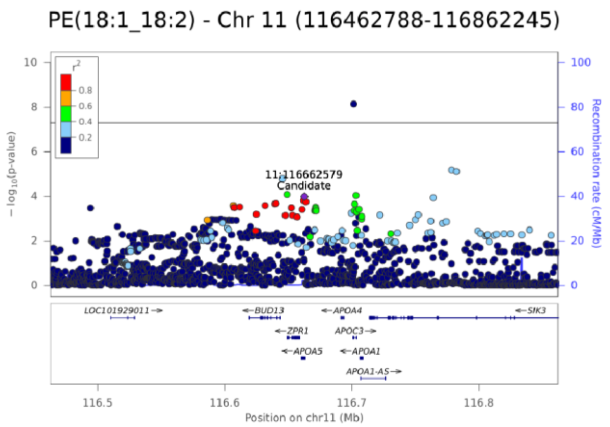
PE(18:1_18:2) - Chr 11 (116462788-116862245)
11:116662579
Candidate
− log10(p-value)
Recombination rate (cM/Mb)
LOC101929011
BUD13
ZPR1
APOA5
APOA4
APOC3
APOA1
APOA1-AS
SIK3
Position on chr11 (Mb)

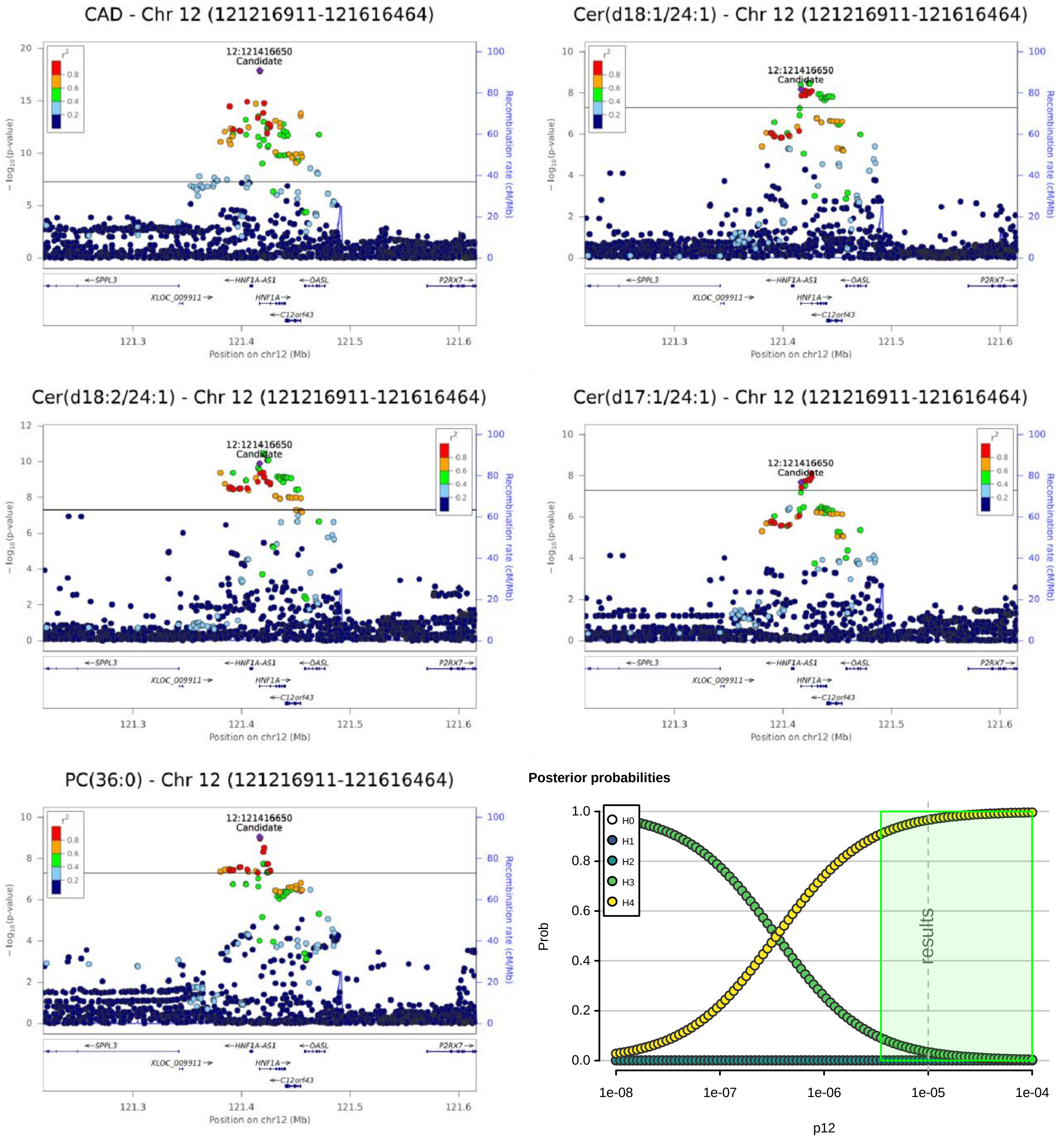
CAD - Chr 12 (121216911-121616464)
Cer(d18:1/24:1) - Chr 12 (121216911-121616464)
Cer(d18:2/24:1) - Chr 12 (121216911-121616464)
Cer(d17:1/24:1) - Chr 12 (121216911-121616464)
PC(36:0) - Chr 12 (121216911-121616464)
12:121416650
Candidate
− log10(p-value)
Recombination rate (cM/Mb)
SPPL3
HNF1A-AS1
OASL
P2RX7
XLOC_009911
HNF1A
C12orf43
121.3
121.4
121.5
121.6
Position on chr12 (Mb)
Posterior probabilities
H0
H1
H2
H3
H4
Prob
results
1e-08
1e-07
1e-06
1e-05
1e-04
p12

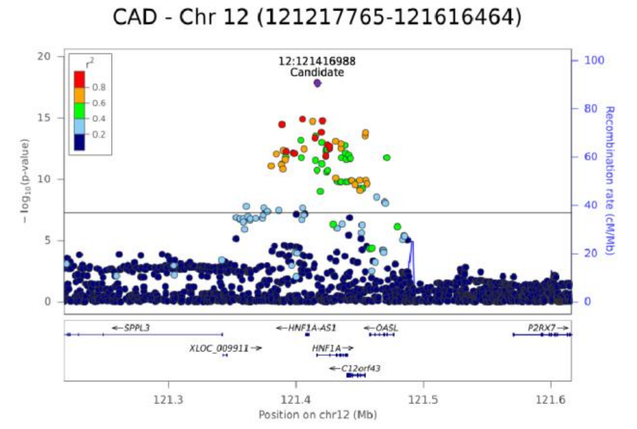


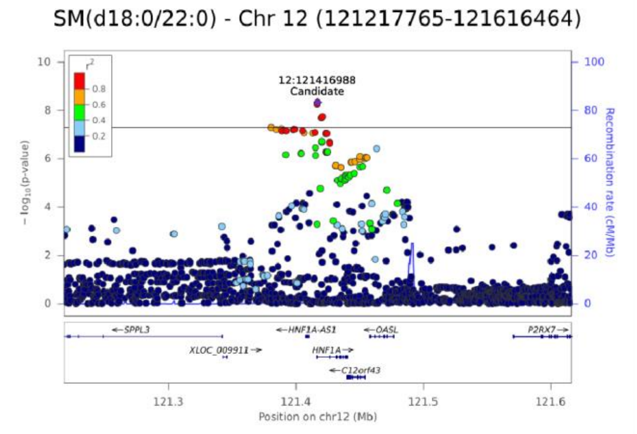


**Posterior probabilities**

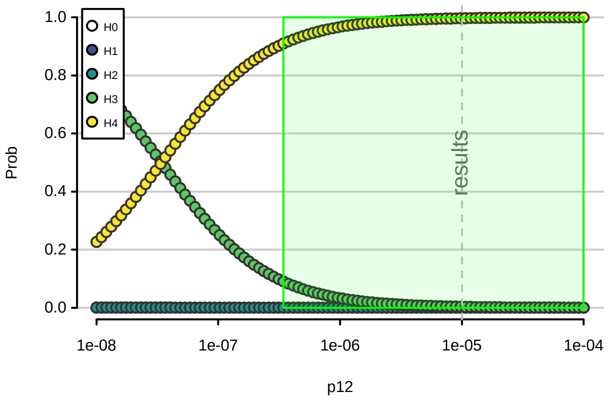

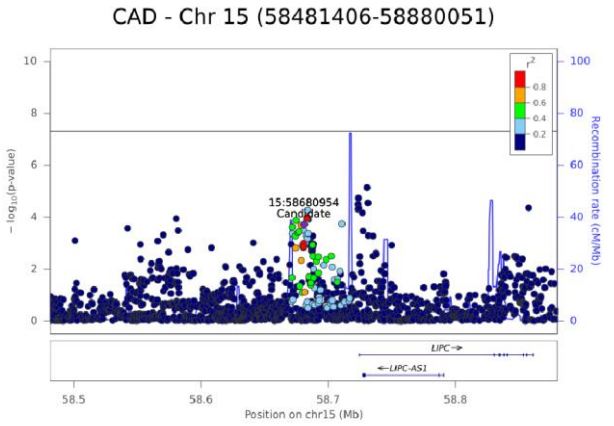
CAD - Chr 15 (58481406-58880051)
15:58680954
Candidate
r²
0.8
0.6
0.4
0.2
− log10(p-value)
Recombination rate (cM/Mb)
LIPC
LIPC-AS1
58.5
58.6
58.7
58.8
Position on chr15 (Mb)

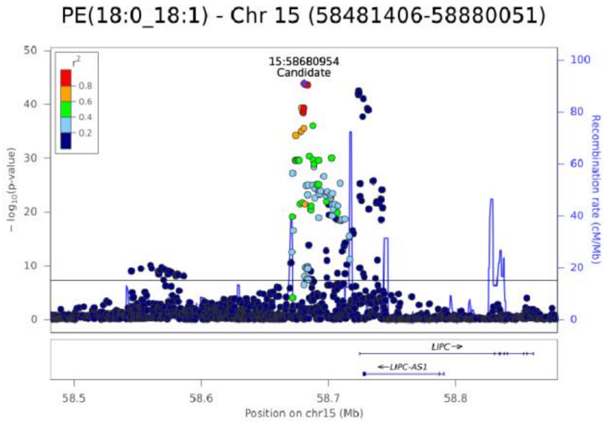
PE(18:0_18:1) - Chr 15 (58481406-58880051)
15:58680954
Candidate
r²
0.8
0.6
0.4
0.2
− log10(p-value)
Recombination rate (cM/Mb)
LIPC
LIPC-AS1
58.5
58.6
58.7
58.8
Position on chr15 (Mb)

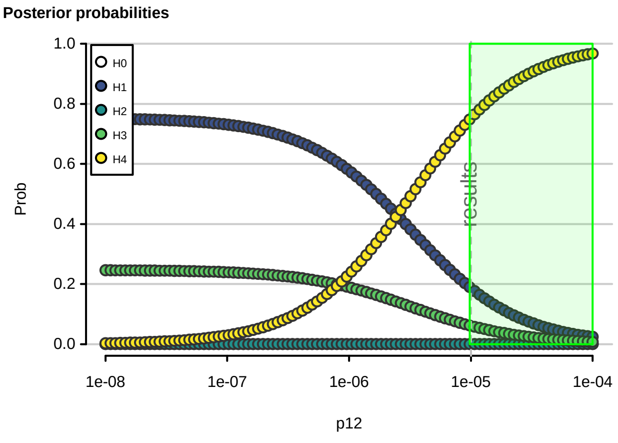
Posterior probabilities
H0
H1
H2
H3
H4
Prob
results
1e-08
1e-07
1e-06
1e-05
1e-04
p12

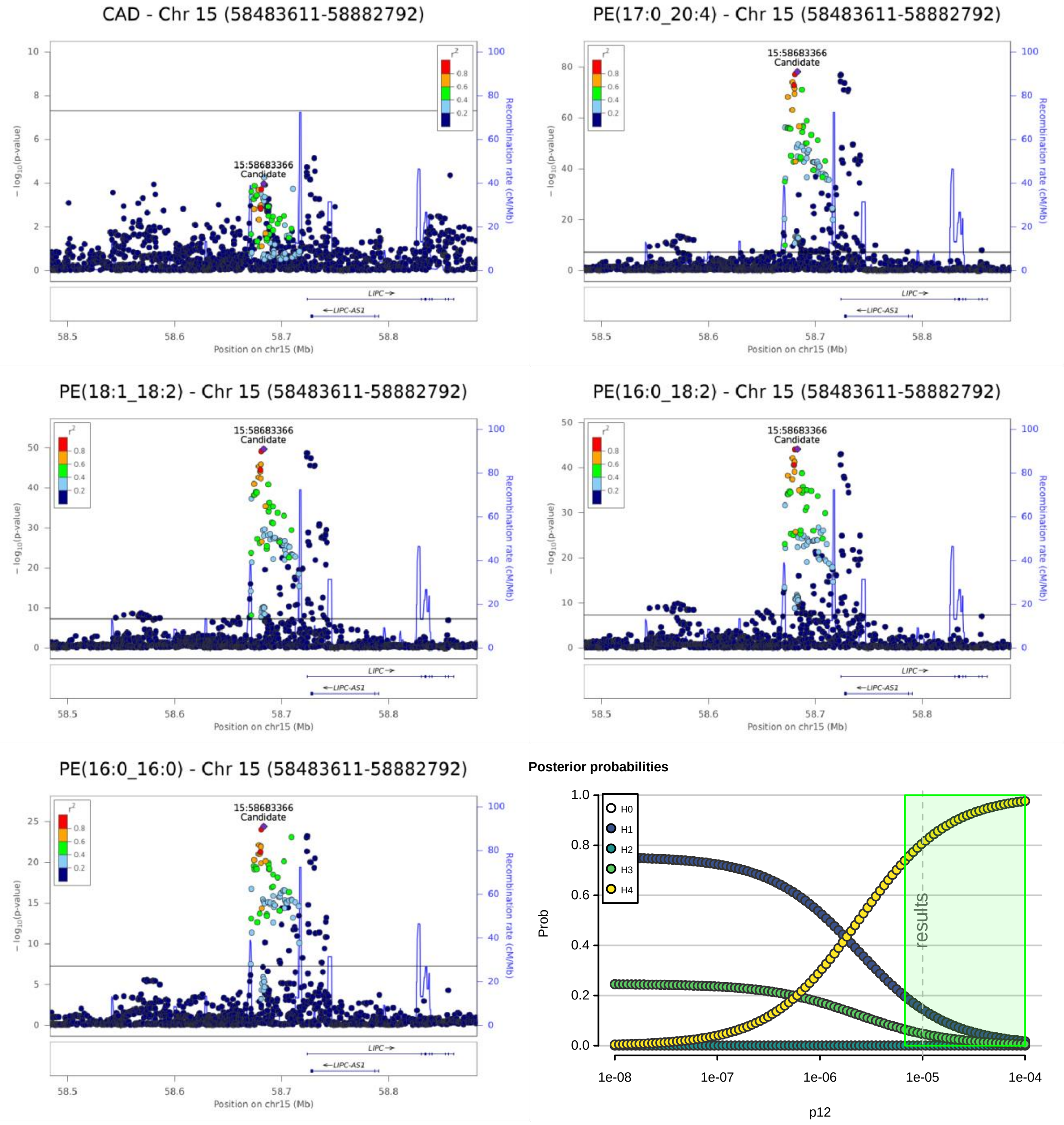
CAD - Chr 15 (58483611-58882792)
PE(17:0_20:4) - Chr 15 (58483611-58882792)
PE(18:1_18:2) - Chr 15 (58483611-58882792)
PE(16:0_18:2) - Chr 15 (58483611-58882792)
PE(16:0_16:0) - Chr 15 (58483611-58882792)
15:58683366
Candidate
$r^2$
0.8
0.6
0.4
0.2
$-\log_{10}$(p-value)
Recombination rate (cM/Mb)
LIPC→
←LIPC-AS1
Position on chr15 (Mb)
58.5
58.6
58.7
58.8
Posterior probabilities
H0
H1
H2
H3
H4
Prob
results
p12
1e-08
1e-07
1e-06
1e-05
1e-04

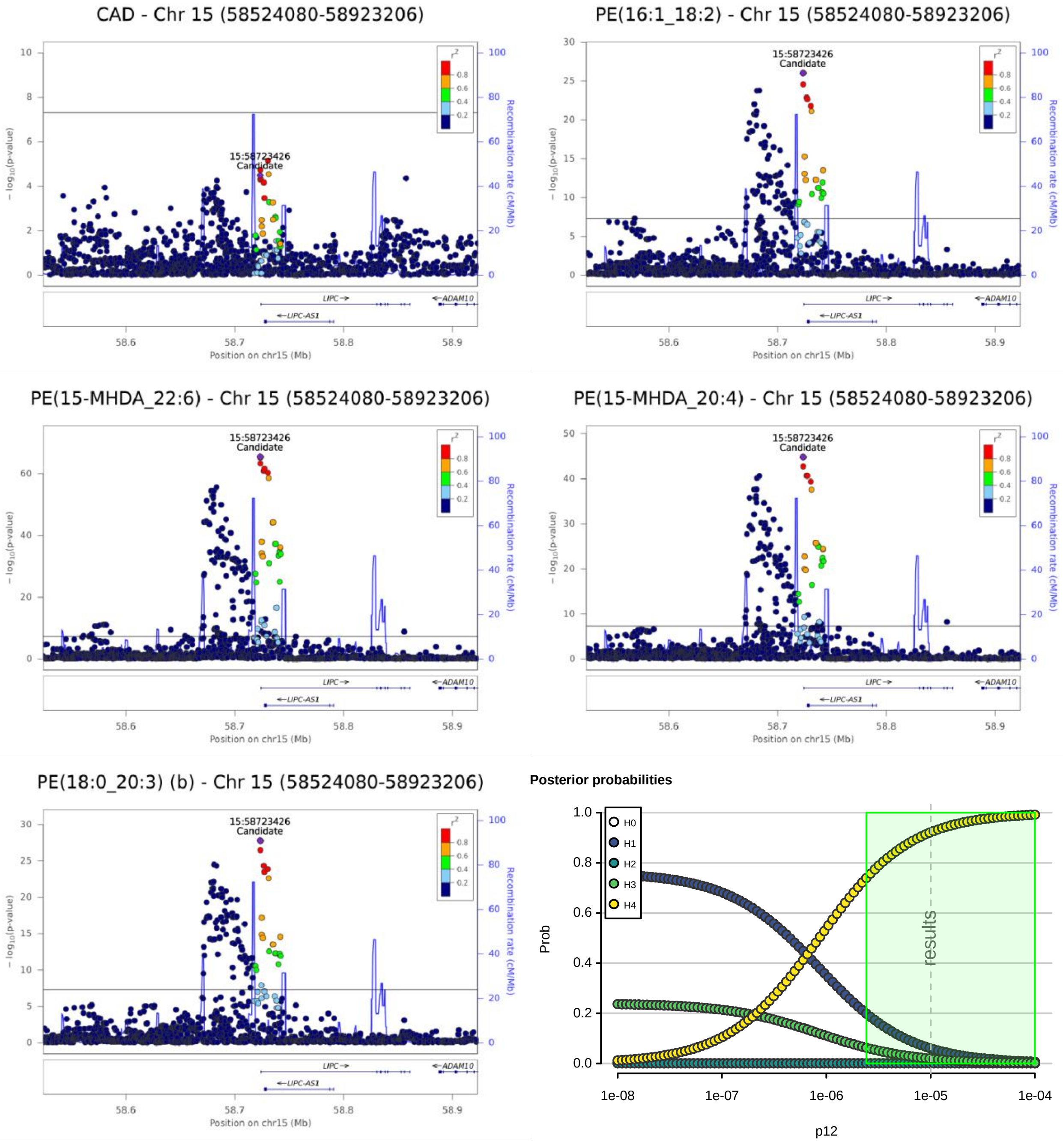
CAD - Chr 15 (58524080-58923206)
PE(16:1_18:2) - Chr 15 (58524080-58923206)
PE(15-MHDA_22:6) - Chr 15 (58524080-58923206)
PE(15-MHDA_20:4) - Chr 15 (58524080-58923206)
PE(18:0_20:3) (b) - Chr 15 (58524080-58923206)
15:58723426
Candidate
− log10(p-value)
Recombination rate (cM/Mb)
Position on chr15 (Mb)
LIPC
LIPC-AS1
ADAM10
Posterior probabilities
Prob
p12
results
H0
H1
H2
H3
H4

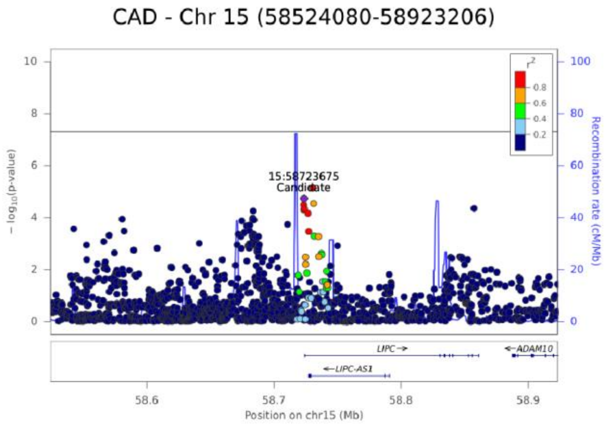
CAD - Chr 15 (58524080-58923206)
15:58723675
Candidate
− log10(p-value)
Recombination rate (cM/Mb)
LIPC
ADAM10
LIPC-AS1
Position on chr15 (Mb)

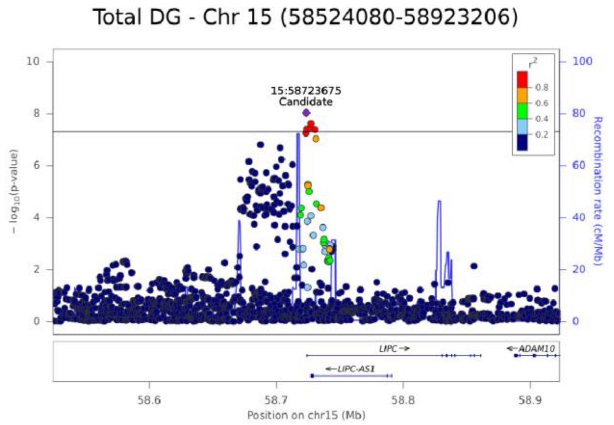
Total DG - Chr 15 (58524080-58923206)
15:58723675
Candidate
− log10(p-value)
Recombination rate (cM/Mb)
LIPC
ADAM10
LIPC-AS1
Position on chr15 (Mb)

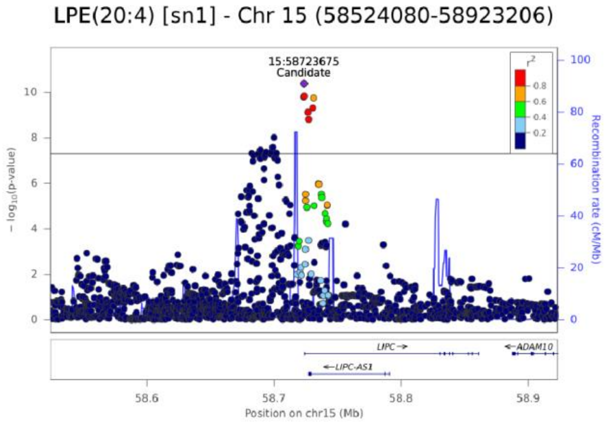
LPE(20:4) [sn1] - Chr 15 (58524080-58923206)
15:58723675
Candidate
− log10(p-value)
Recombination rate (cM/Mb)
LIPC
ADAM10
LIPC-AS1
Position on chr15 (Mb)

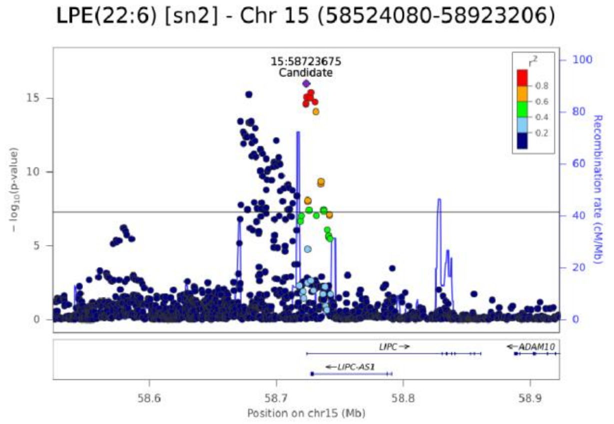
LPE(22:6) [sn2] - Chr 15 (58524080-58923206)
15:58723675
Candidate
− log10(p-value)
Recombination rate (cM/Mb)
LIPC
ADAM10
LIPC-AS1
Position on chr15 (Mb)

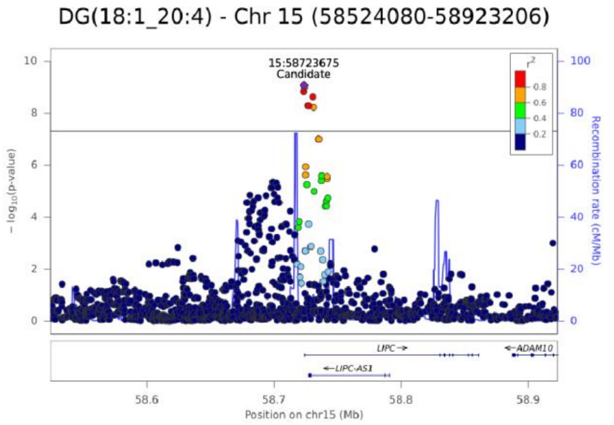
DG(18:1_20:4) - Chr 15 (58524080-58923206)
15:58723675
Candidate
− log10(p-value)
Recombination rate (cM/Mb)
LIPC
ADAM10
LIPC-AS1
Position on chr15 (Mb)

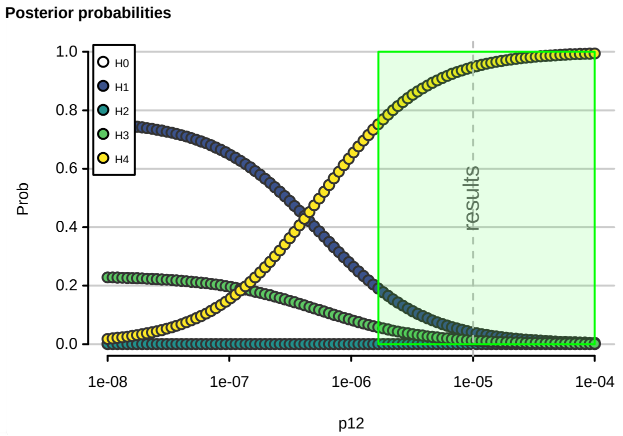
Posterior probabilities
H0
H1
H2
H3
H4
Prob
results
1e-08
1e-07
1e-06
1e-05
1e-04
p12

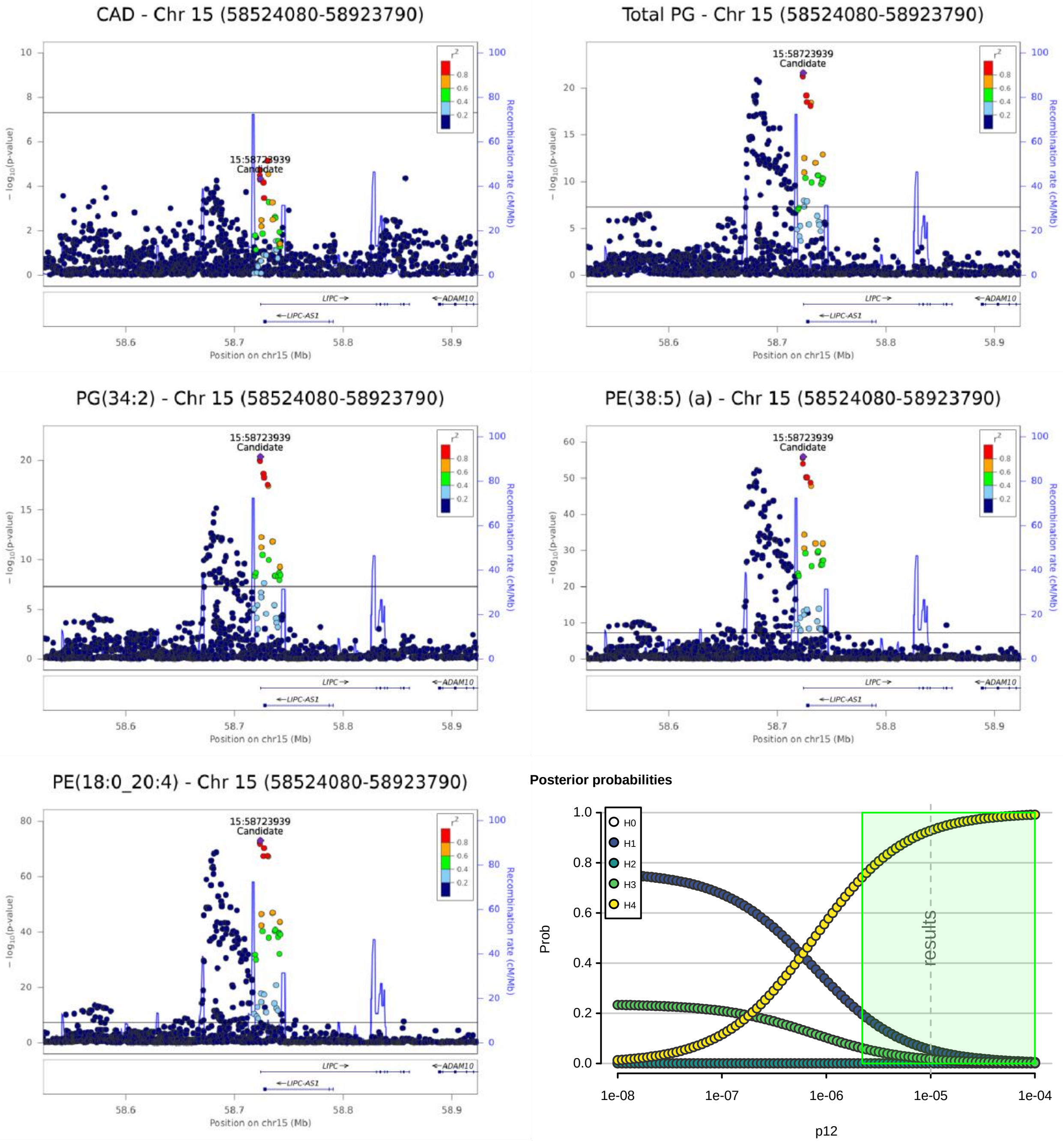
CAD - Chr 15 (58524080-58923790)
Total PG - Chr 15 (58524080-58923790)
PG(34:2) - Chr 15 (58524080-58923790)
PE(38:5) (a) - Chr 15 (58524080-58923790)
PE(18:0_20:4) - Chr 15 (58524080-58923790)
15:58723939
Candidate
− log10(p-value)
Recombination rate (cM/Mb)
LIPC
ADAM10
LIPC-AS1
Position on chr15 (Mb)
Posterior probabilities
H0
H1
H2
H3
H4
Prob
results
p12

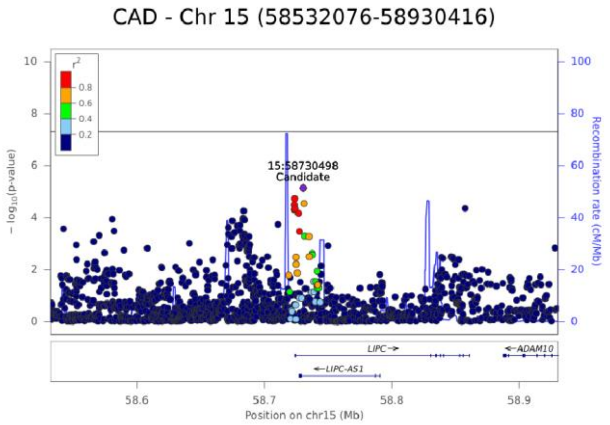
CAD - Chr 15 (58532076-58930416)
15:58730498
Candidate
− log10(p-value)
Recombination rate (cM/Mb)
r2
LIPC→
←ADAM10
←LIPC-AS1
58.6
58.7
58.8
58.9
Position on chr15 (Mb)

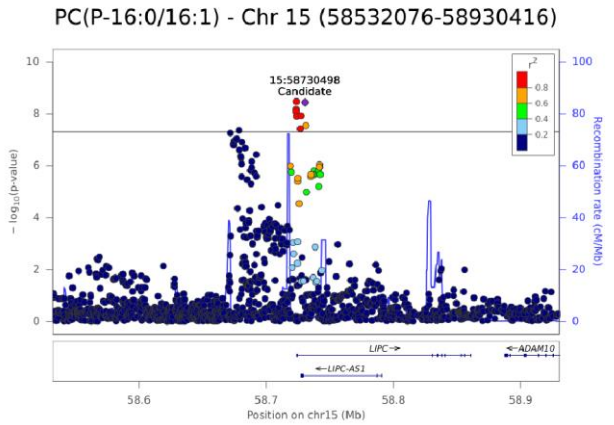
PC(P-16:0/16:1) - Chr 15 (58532076-58930416)
15:58730498
Candidate
− log10(p-value)
Recombination rate (cM/Mb)
r2
LIPC→
←ADAM10
←LIPC-AS1
58.6
58.7
58.8
58.9
Position on chr15 (Mb)

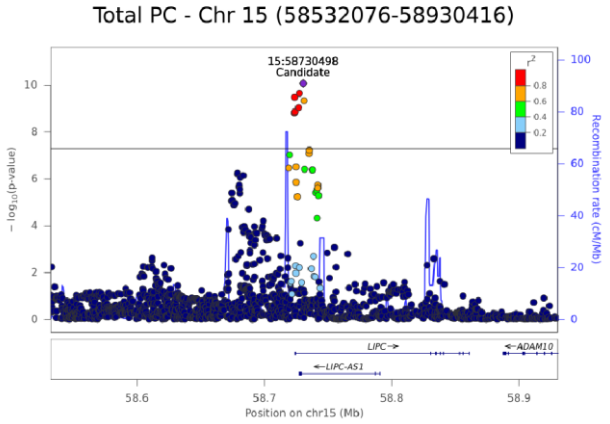
Total PC - Chr 15 (58532076-58930416)
15:58730498
Candidate
− log10(p-value)
Recombination rate (cM/Mb)
r2
LIPC→
←ADAM10
←LIPC-AS1
58.6
58.7
58.8
58.9
Position on chr15 (Mb)

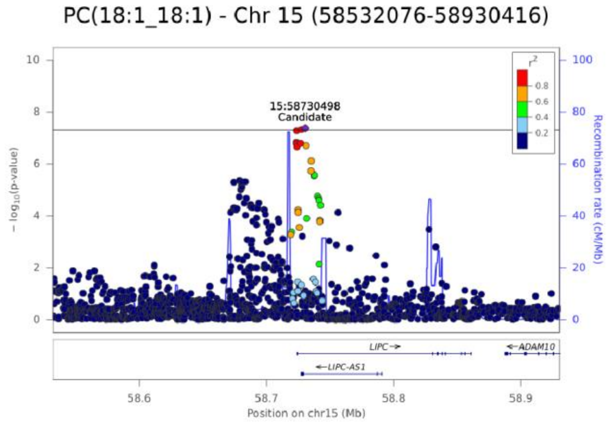
PC(18:1_18:1) - Chr 15 (58532076-58930416)
15:58730498
Candidate
− log10(p-value)
Recombination rate (cM/Mb)
r2
LIPC→
←ADAM10
←LIPC-AS1
58.6
58.7
58.8
58.9
Position on chr15 (Mb)

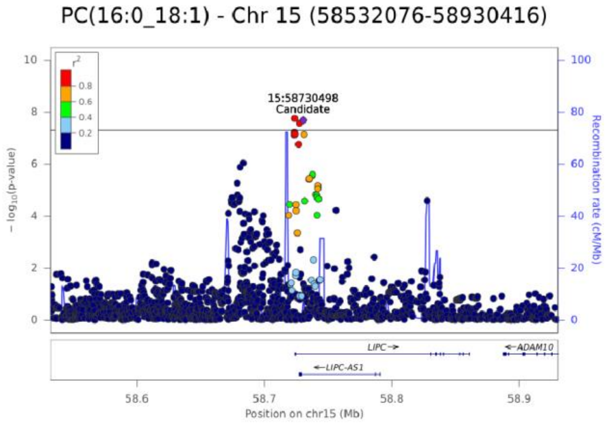
PC(16:0_18:1) - Chr 15 (58532076-58930416)
15:58730498
Candidate
− log10(p-value)
Recombination rate (cM/Mb)
r2
LIPC→
←ADAM10
←LIPC-AS1
58.6
58.7
58.8
58.9
Position on chr15 (Mb)

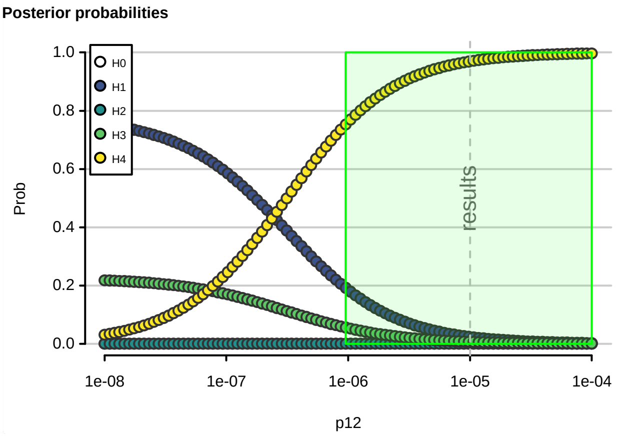
Posterior probabilities
H0
H1
H2
H3
H4
Prob
results
1e-08
1e-07
1e-06
1e-05
1e-04
p12

CAD - Chr 15 (58532076-58931039)

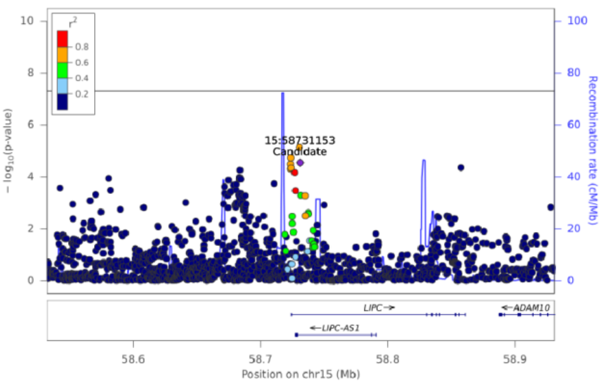


LPE(20:4) [sn2] - Chr 15 (58532076-58931039)

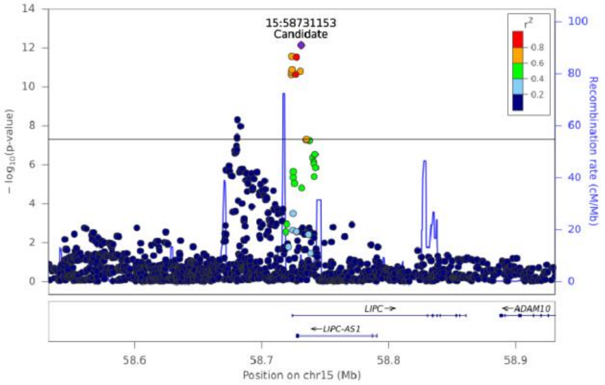


LPE(20:4) [sn1] - Chr 15 (58532076-58931039)

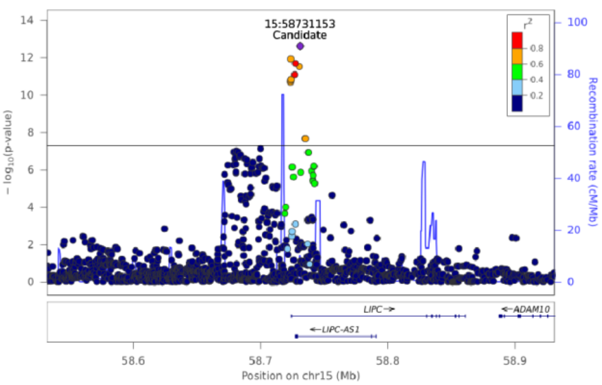


**Posterior probabilities**

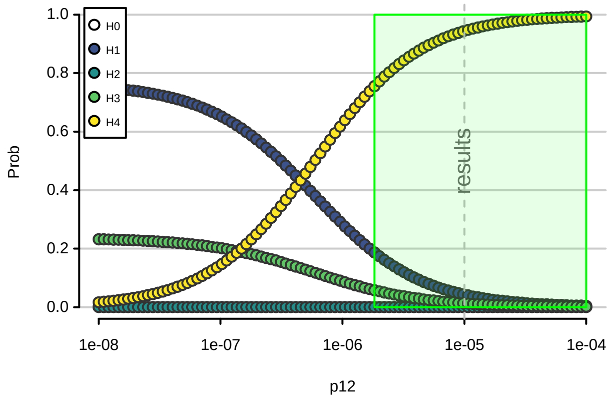


TG(56:6) [NL-20:4] - Chr 15 (58532076-58931039)

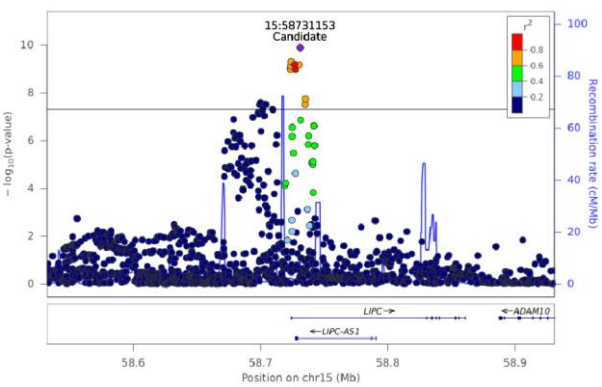

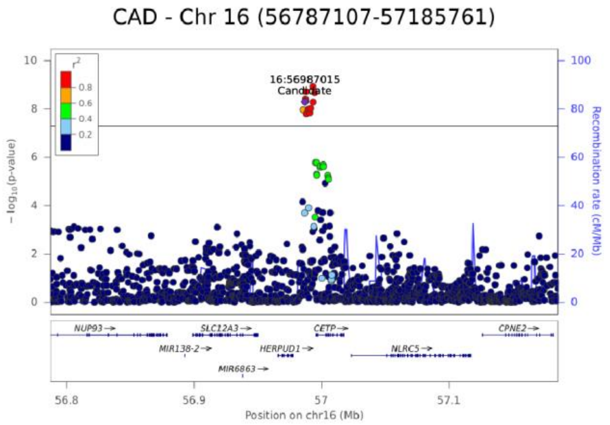
CAD - Chr 16 (56787107-57185761)
16:56987015
Candidate
− log10(p-value)
Recombination rate (cM/Mb)
NUP93
SLC12A3
CETP
CPNE2
MIR138-2
HERPUD1
NLRC5
MIR6863
Position on chr16 (Mb)

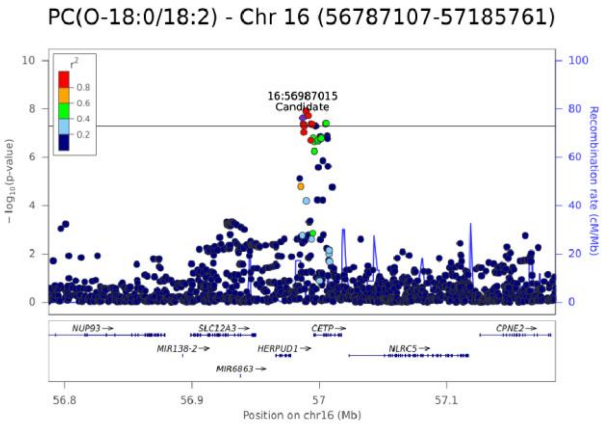
PC(O-18:0/18:2) - Chr 16 (56787107-57185761)
16:56987015
Candidate
− log10(p-value)
Recombination rate (cM/Mb)
NUP93
SLC12A3
CETP
CPNE2
MIR138-2
HERPUD1
NLRC5
MIR6863
Position on chr16 (Mb)

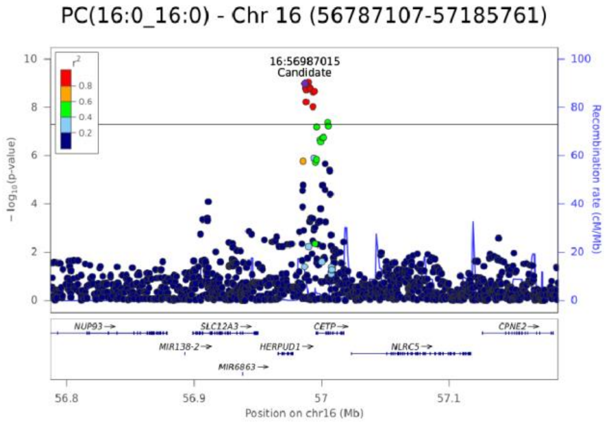
PC(16:0_16:0) - Chr 16 (56787107-57185761)
16:56987015
Candidate
− log10(p-value)
Recombination rate (cM/Mb)
NUP93
SLC12A3
CETP
CPNE2
MIR138-2
HERPUD1
NLRC5
MIR6863
Position on chr16 (Mb)

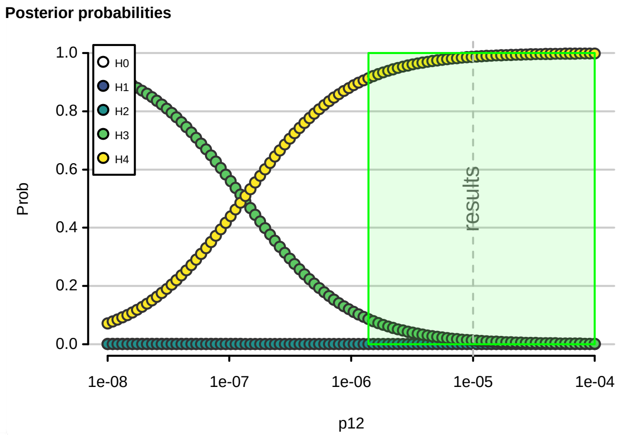
Posterior probabilities
H0
H1
H2
H3
H4
Prob
results
p12

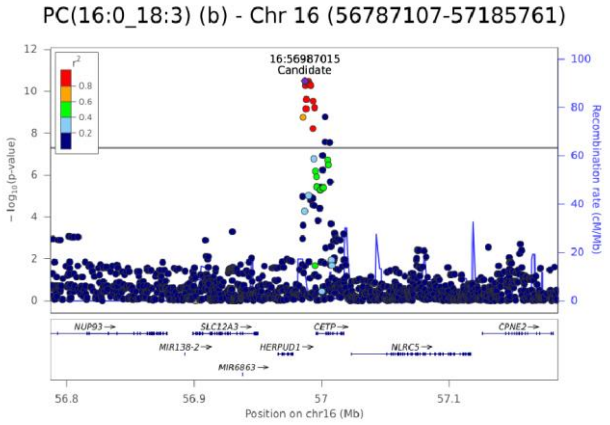
PC(16:0_18:3) (b) - Chr 16 (56787107-57185761)
16:56987015
Candidate
− log10(p-value)
Recombination rate (cM/Mb)
NUP93
SLC12A3
CETP
CPNE2
MIR138-2
HERPUD1
NLRC5
MIR6863
Position on chr16 (Mb)

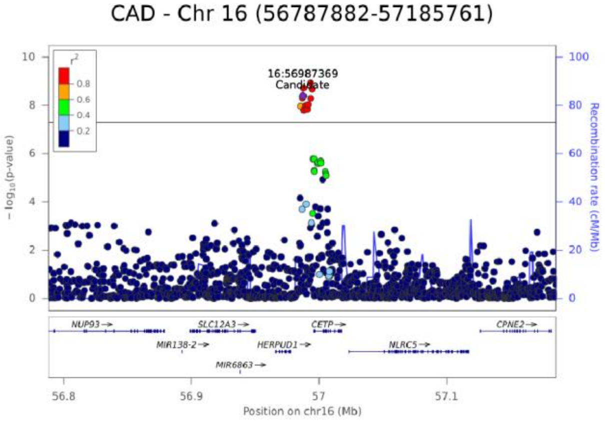
CAD - Chr 16 (56787882-57185761)
16:56987369
Candidate
r²
− log10(p-value)
Recombination rate (cM/Mb)
NUP93
SLC12A3
CETP
CPNE2
MIR138-2
HERPUD1
NLRC5
MIR6863
Position on chr16 (Mb)

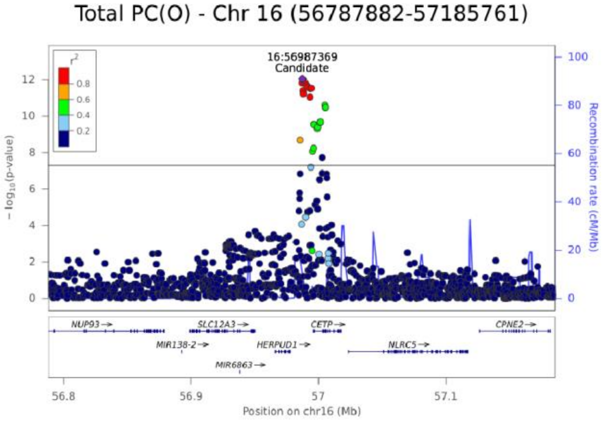
Total PC(O) - Chr 16 (56787882-57185761)
16:56987369
Candidate
r²
− log10(p-value)
Recombination rate (cM/Mb)
NUP93
SLC12A3
CETP
CPNE2
MIR138-2
HERPUD1
NLRC5
MIR6863
Position on chr16 (Mb)

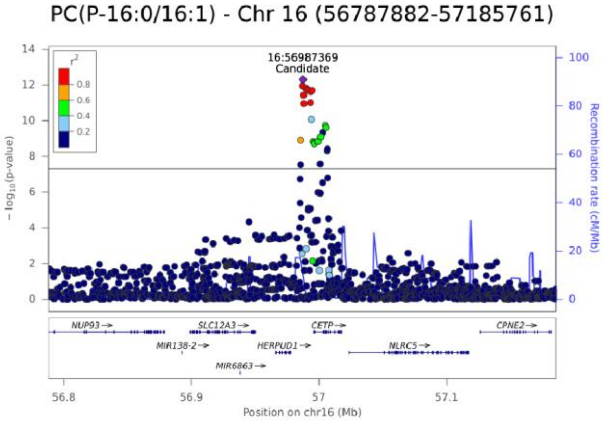
PC(P-16:0/16:1) - Chr 16 (56787882-57185761)
16:56987369
Candidate
r²
− log10(p-value)
Recombination rate (cM/Mb)
NUP93
SLC12A3
CETP
CPNE2
MIR138-2
HERPUD1
NLRC5
MIR6863
Position on chr16 (Mb)

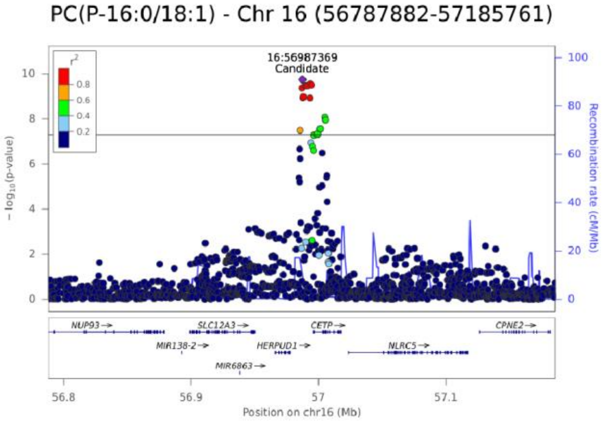
PC(P-16:0/18:1) - Chr 16 (56787882-57185761)
16:56987369
Candidate
r²
− log10(p-value)
Recombination rate (cM/Mb)
NUP93
SLC12A3
CETP
CPNE2
MIR138-2
HERPUD1
NLRC5
MIR6863
Position on chr16 (Mb)

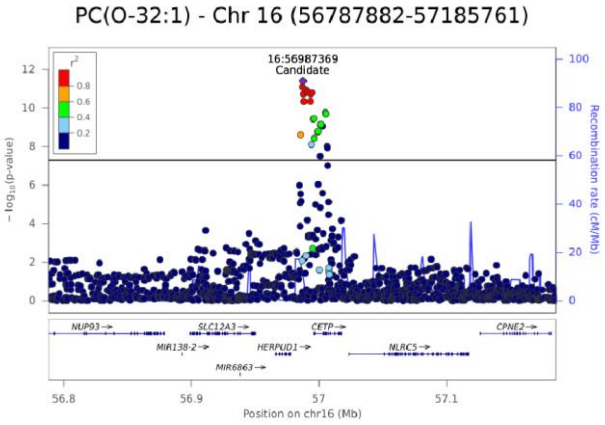
PC(O-32:1) - Chr 16 (56787882-57185761)
16:56987369
Candidate
r²
− log10(p-value)
Recombination rate (cM/Mb)
NUP93
SLC12A3
CETP
CPNE2
MIR138-2
HERPUD1
NLRC5
MIR6863
Position on chr16 (Mb)

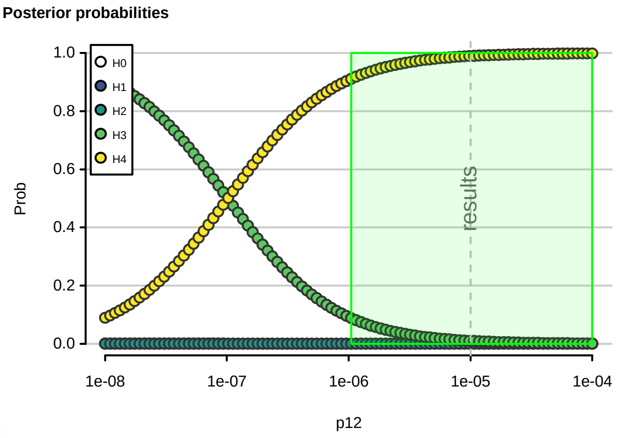
Posterior probabilities
Prob
H0
H1
H2
H3
H4
results
p12

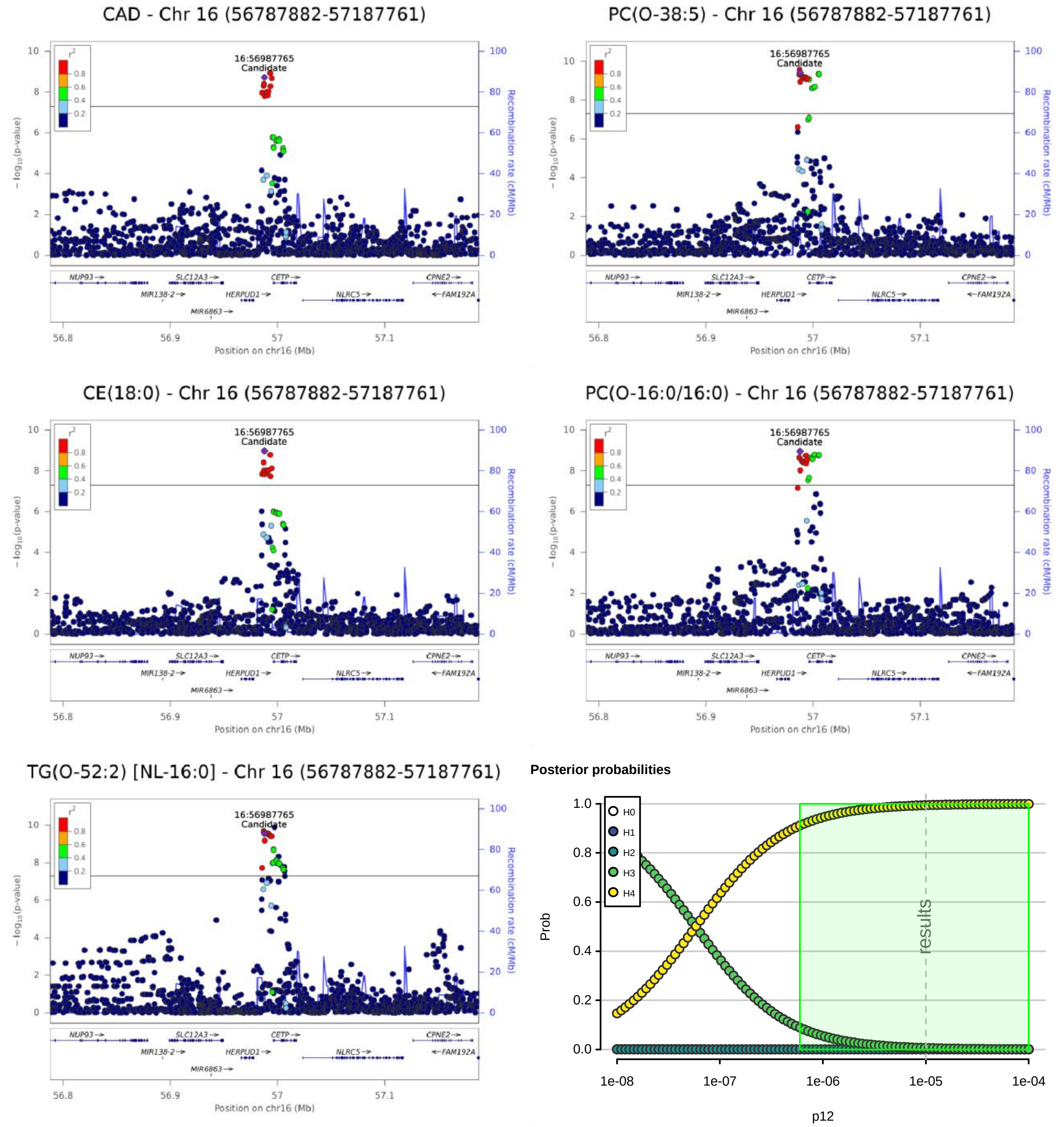
CAD - Chr 16 (56787882-57187761)
PC(O-38:5) - Chr 16 (56787882-57187761)
CE(18:0) - Chr 16 (56787882-57187761)
PC(O-16:0/16:0) - Chr 16 (56787882-57187761)
TG(O-52:2) [NL-16:0] - Chr 16 (56787882-57187761)
16:56987765
Candidate
− log10 (p-value)
Recombination rate (cM/Mb)
Position on chr16 (Mb)
NUP93
SLC12A3
CETP
CPNE2
MIR138-2
HERPUD1
NLRC5
FAM192A
MIR6863
Posterior probabilities
Prob
p12
H0
H1
H2
H3
H4
results

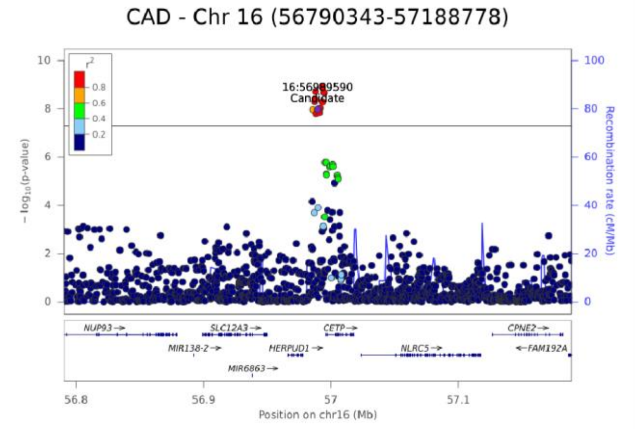


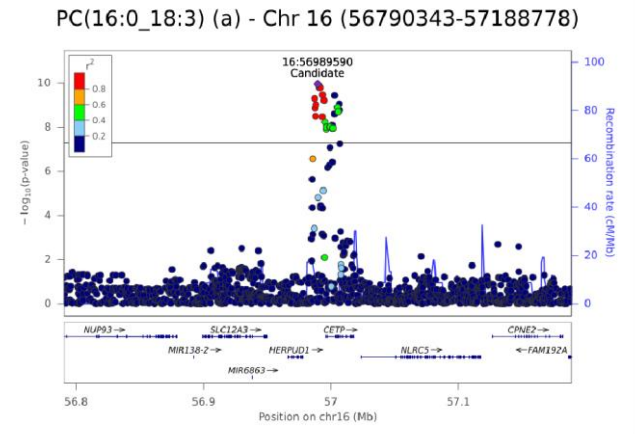


**Posterior probabilities**

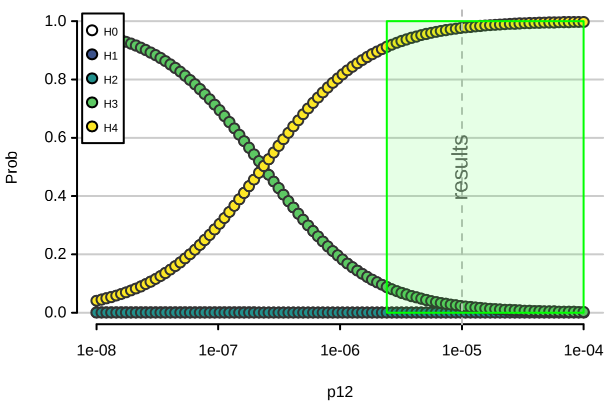

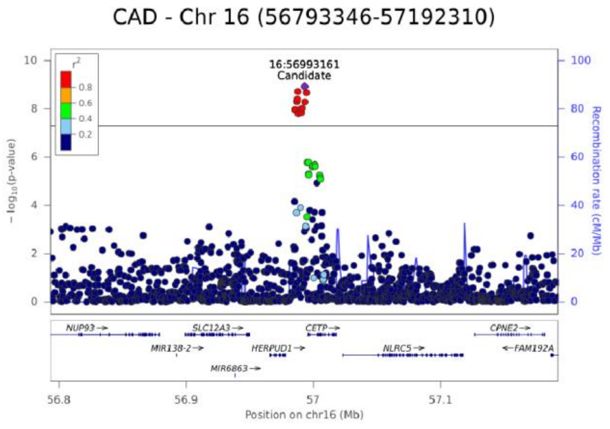
CAD - Chr 16 (56793346-57192310)
16:56993161
Candidate
r2
0.8
0.6
0.4
0.2
- log10(p-value)
Recombination rate (cM/Mb)
NUP93
SLC12A3
CETP
CPNE2
MIR138-2
HERPUD1
NLRC5
FAM192A
MIR6863
56.8
56.9
57
57.1
Position on chr16 (Mb)

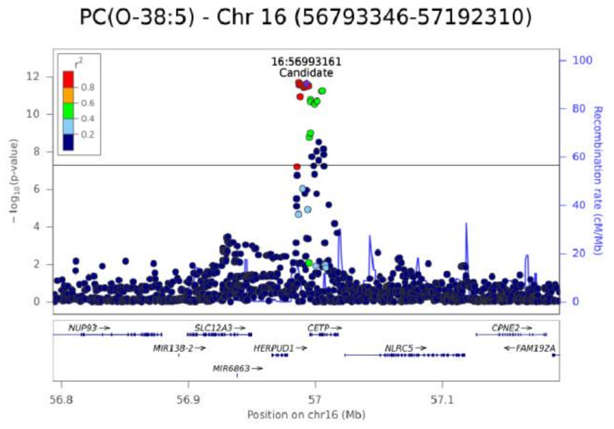
PC(O-38:5) - Chr 16 (56793346-57192310)
16:56993161
Candidate
r2
0.8
0.6
0.4
0.2
- log10(p-value)
Recombination rate (cM/Mb)
NUP93
SLC12A3
CETP
CPNE2
MIR138-2
HERPUD1
NLRC5
FAM192A
MIR6863
56.8
56.9
57
57.1
Position on chr16 (Mb)

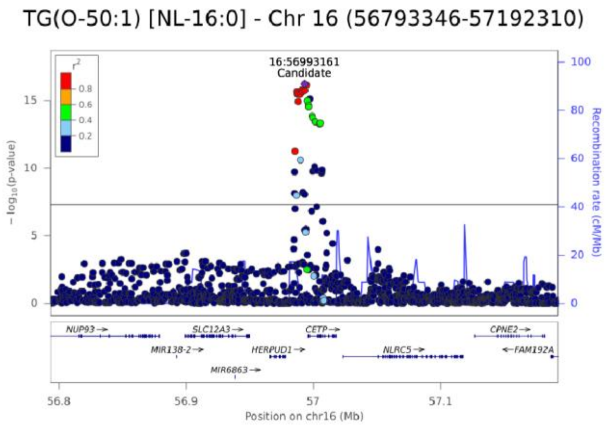
TG(O-50:1) [NL-16:0] - Chr 16 (56793346-57192310)
16:56993161
Candidate
r2
0.8
0.6
0.4
0.2
- log10(p-value)
Recombination rate (cM/Mb)
NUP93
SLC12A3
CETP
CPNE2
MIR138-2
HERPUD1
NLRC5
FAM192A
MIR6863
56.8
56.9
57
57.1
Position on chr16 (Mb)

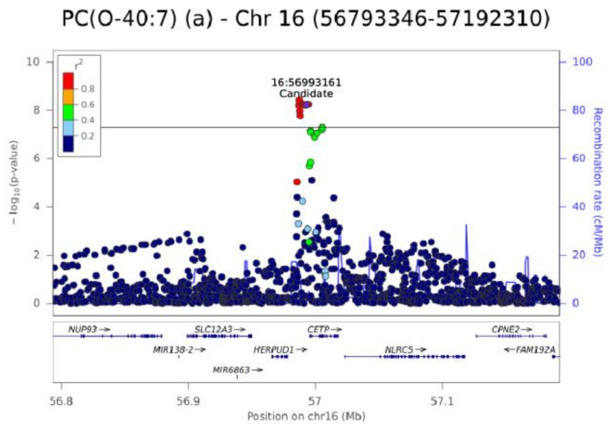
PC(O-40:7) (a) - Chr 16 (56793346-57192310)
16:56993161
Candidate
r2
0.8
0.6
0.4
0.2
- log10(p-value)
Recombination rate (cM/Mb)
NUP93
SLC12A3
CETP
CPNE2
MIR138-2
HERPUD1
NLRC5
FAM192A
MIR6863
56.8
56.9
57
57.1
Position on chr16 (Mb)

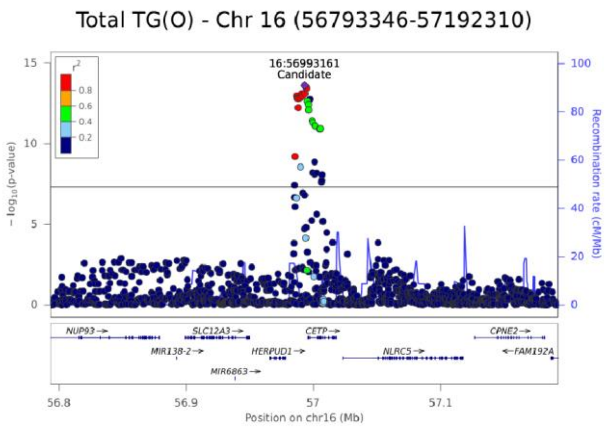
Total TG(O) - Chr 16 (56793346-57192310)
16:56993161
Candidate
r2
0.8
0.6
0.4
0.2
- log10(p-value)
Recombination rate (cM/Mb)
NUP93
SLC12A3
CETP
CPNE2
MIR138-2
HERPUD1
NLRC5
FAM192A
MIR6863
56.8
56.9
57
57.1
Position on chr16 (Mb)

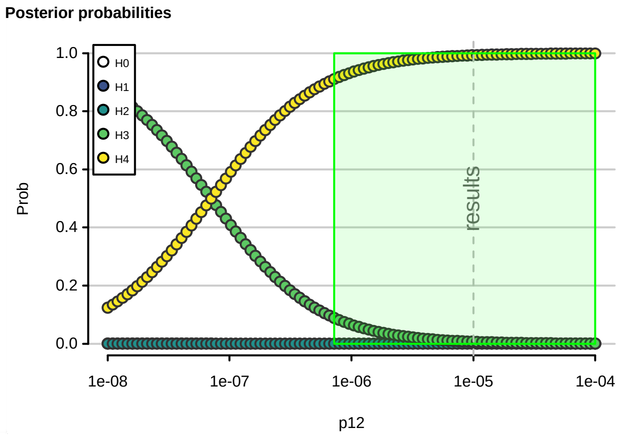
Posterior probabilities
H0
H1
H2
H3
H4
Prob
results
1e-08
1e-07
1e-06
1e-05
1e-04
p12

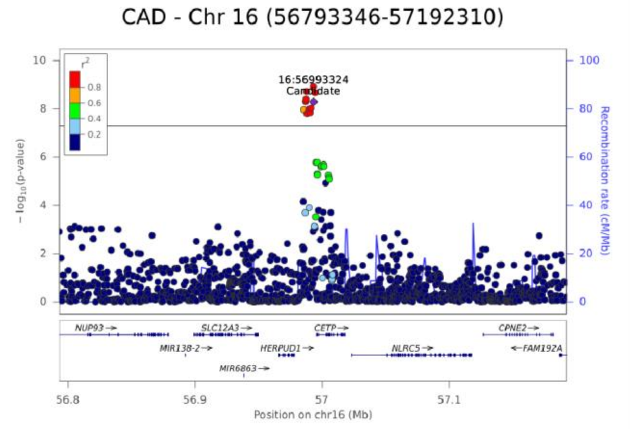


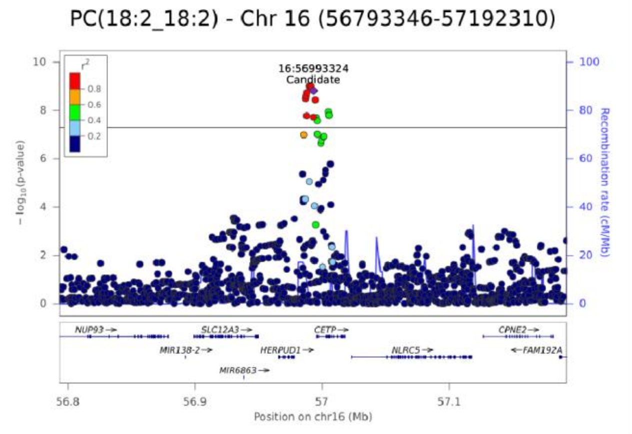


**Posterior probabilities**

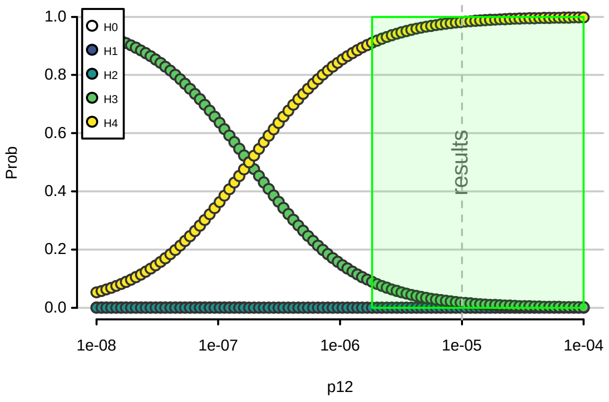

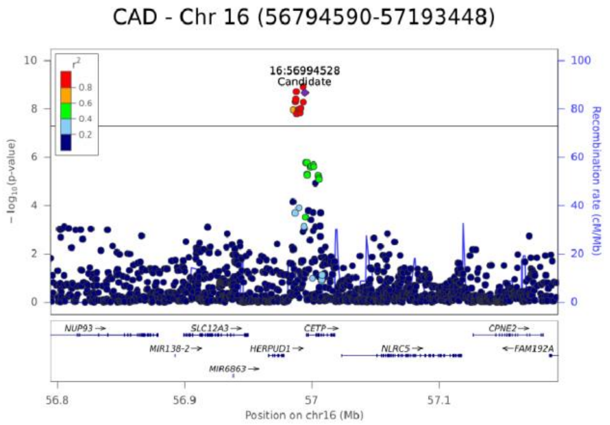
CAD - Chr 16 (56794590-57193448)
16:56994528
Candidate
− log10(p-value)
Recombination rate (cM/Mb)
NUP93
SLC12A3
CETP
CPNE2
MIR138-2
HERPUD1
NLRC5
FAM192A
MIR6863
Position on chr16 (Mb)

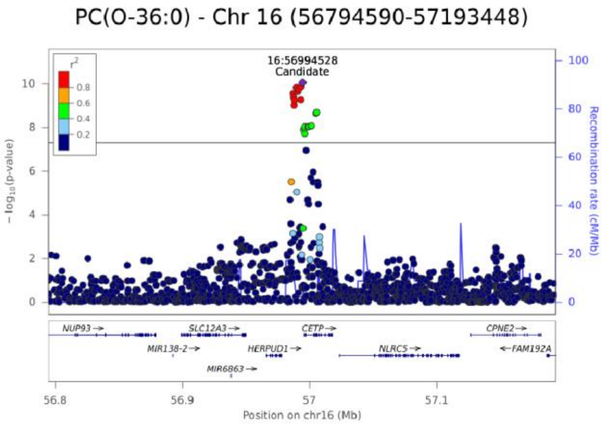
PC(O-36:0) - Chr 16 (56794590-57193448)
16:56994528
Candidate
− log10(p-value)
Recombination rate (cM/Mb)
NUP93
SLC12A3
CETP
CPNE2
MIR138-2
HERPUD1
NLRC5
FAM192A
MIR6863
Position on chr16 (Mb)

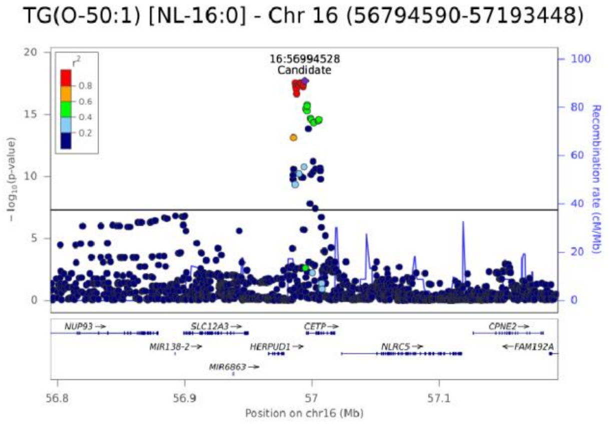
TG(O-50:1) [NL-16:0] - Chr 16 (56794590-57193448)
16:56994528
Candidate
− log10(p-value)
Recombination rate (cM/Mb)
NUP93
SLC12A3
CETP
CPNE2
MIR138-2
HERPUD1
NLRC5
FAM192A
MIR6863
Position on chr16 (Mb)

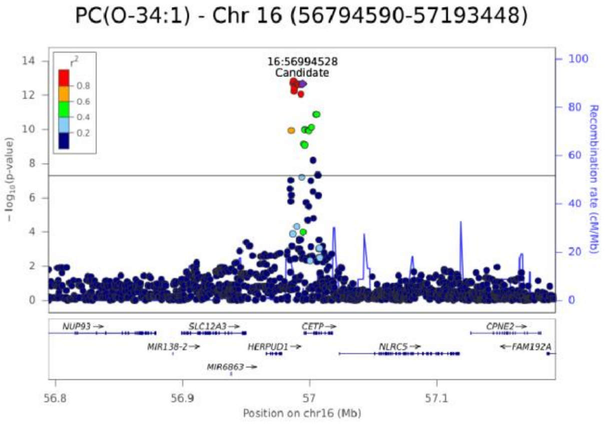
PC(O-34:1) - Chr 16 (56794590-57193448)
16:56994528
Candidate
− log10(p-value)
Recombination rate (cM/Mb)
NUP93
SLC12A3
CETP
CPNE2
MIR138-2
HERPUD1
NLRC5
FAM192A
MIR6863
Position on chr16 (Mb)

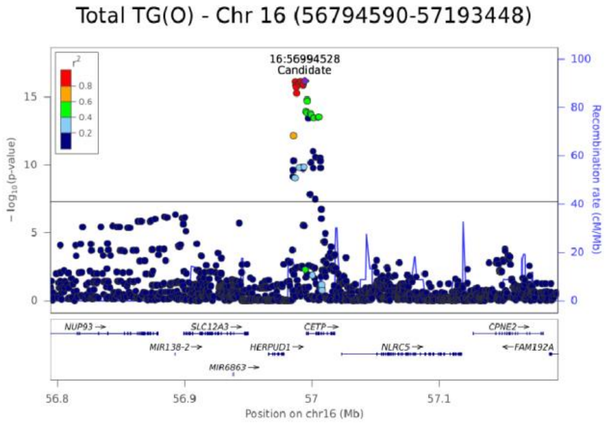
Total TG(O) - Chr 16 (56794590-57193448)
16:56994528
Candidate
− log10(p-value)
Recombination rate (cM/Mb)
NUP93
SLC12A3
CETP
CPNE2
MIR138-2
HERPUD1
NLRC5
FAM192A
MIR6863
Position on chr16 (Mb)

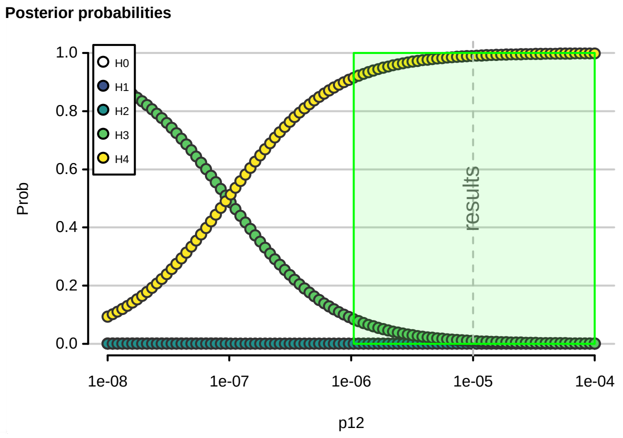
Posterior probabilities
H0
H1
H2
H3
H4
Prob
results
p12

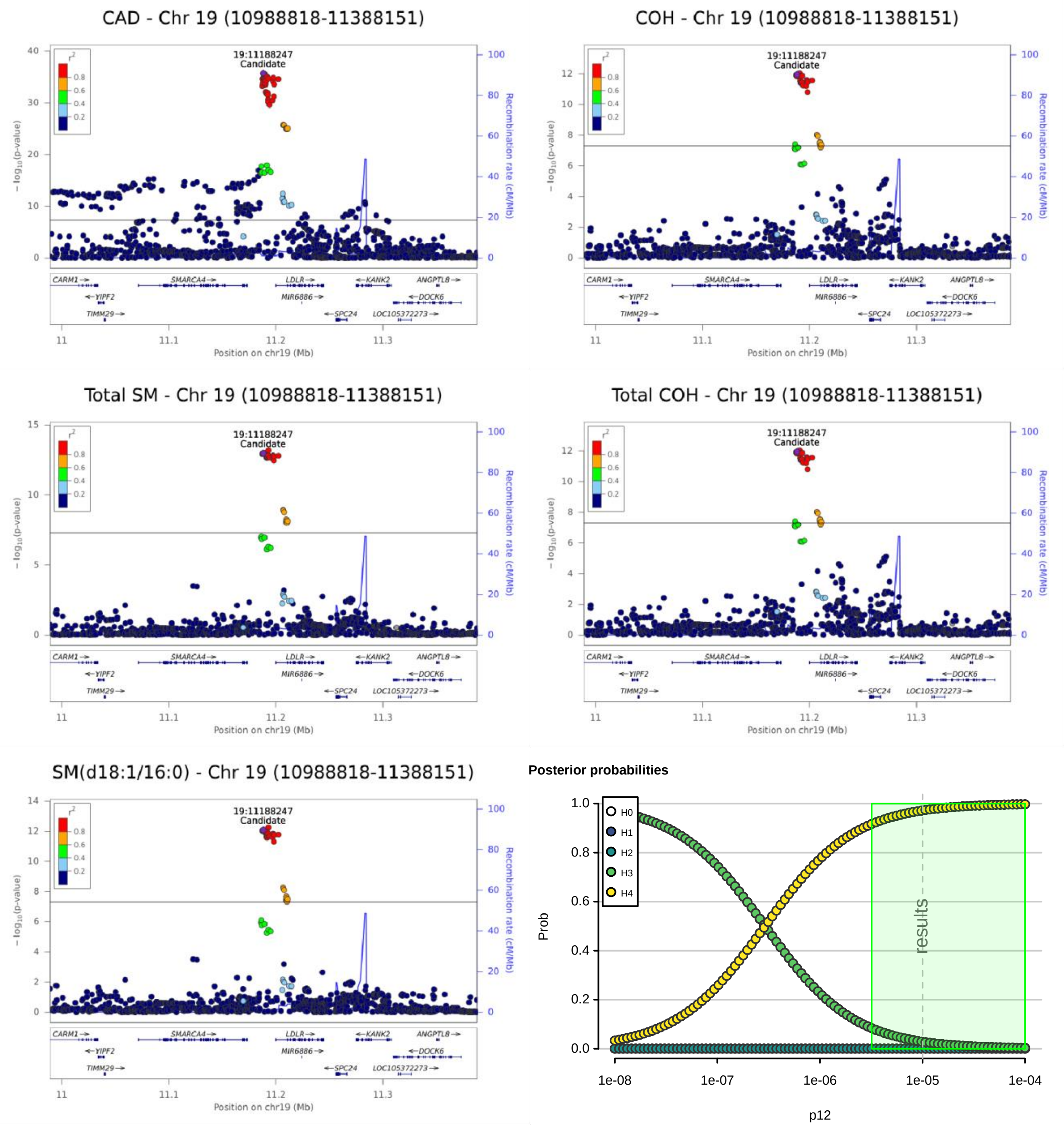
CAD - Chr 19 (10988818-11388151)
COH - Chr 19 (10988818-11388151)
Total SM - Chr 19 (10988818-11388151)
Total COH - Chr 19 (10988818-11388151)
SM(d18:1/16:0) - Chr 19 (10988818-11388151)
19:11188247
Candidate
− log10(p-value)
Recombination rate (cM/Mb)
Position on chr19 (Mb)
CARM1
SMARCA4
LDLR
KANK2
ANGPTL8
YIPF2
MIR6886
DOCK6
TIMM29
SPC24
LOC105372273
Posterior probabilities
H0
H1
H2
H3
H4
Prob
results
p12

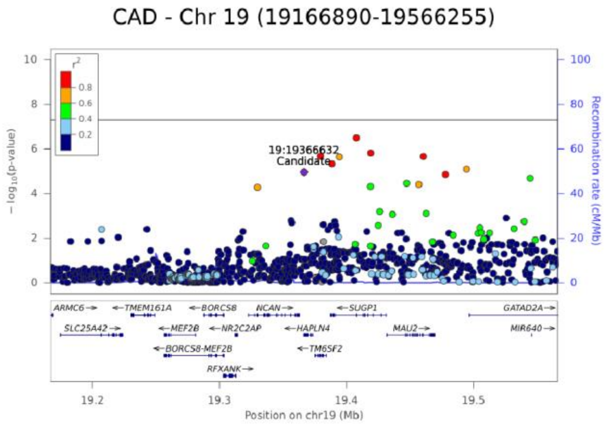


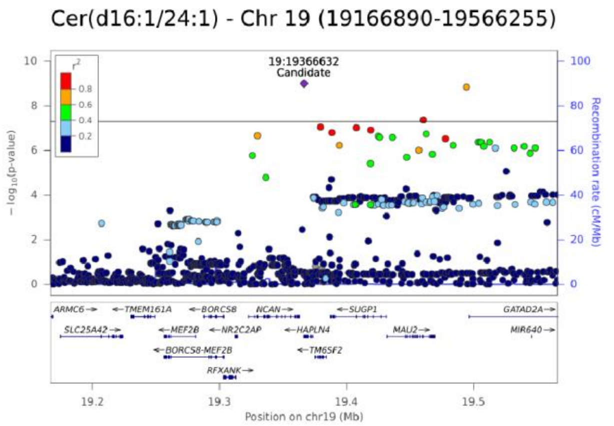


**Posterior probabilities**

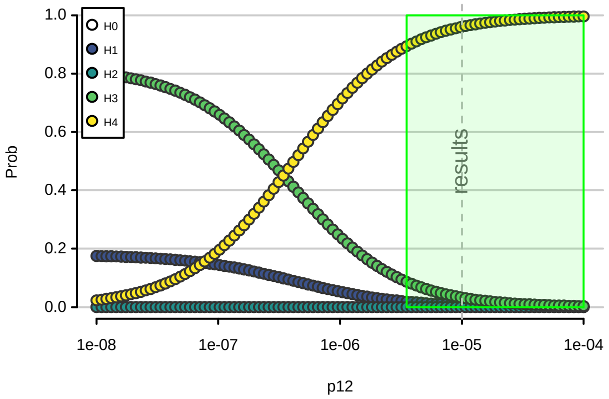

CAD - Chr 19 (19179819-19579046)

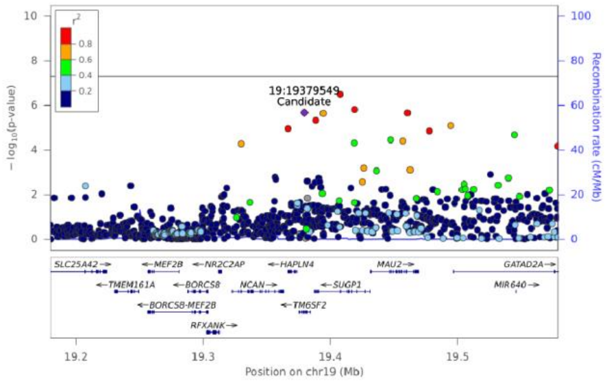


Posterior probabilities

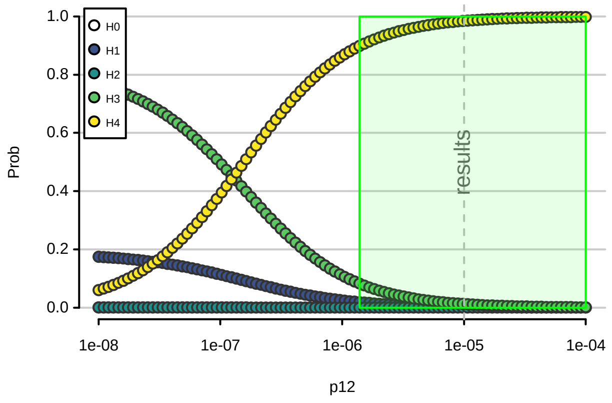


LPC(20:3) [sn1] - Chr 19 (19179819-19579046)

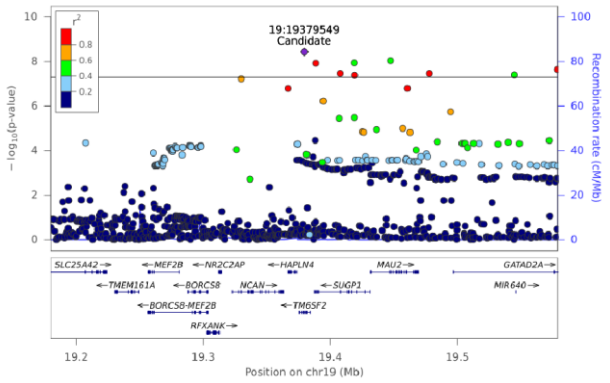


PC(18:1_20:3) - Chr 19 (19179819-19579046)

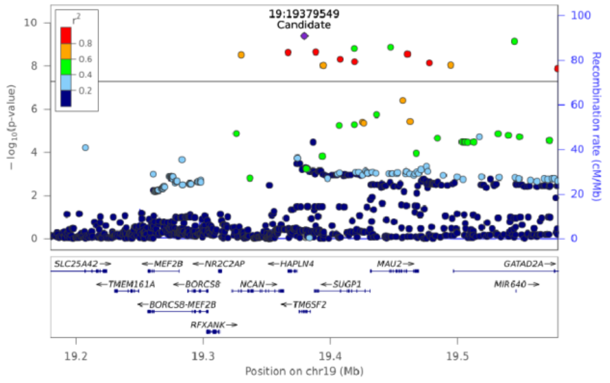

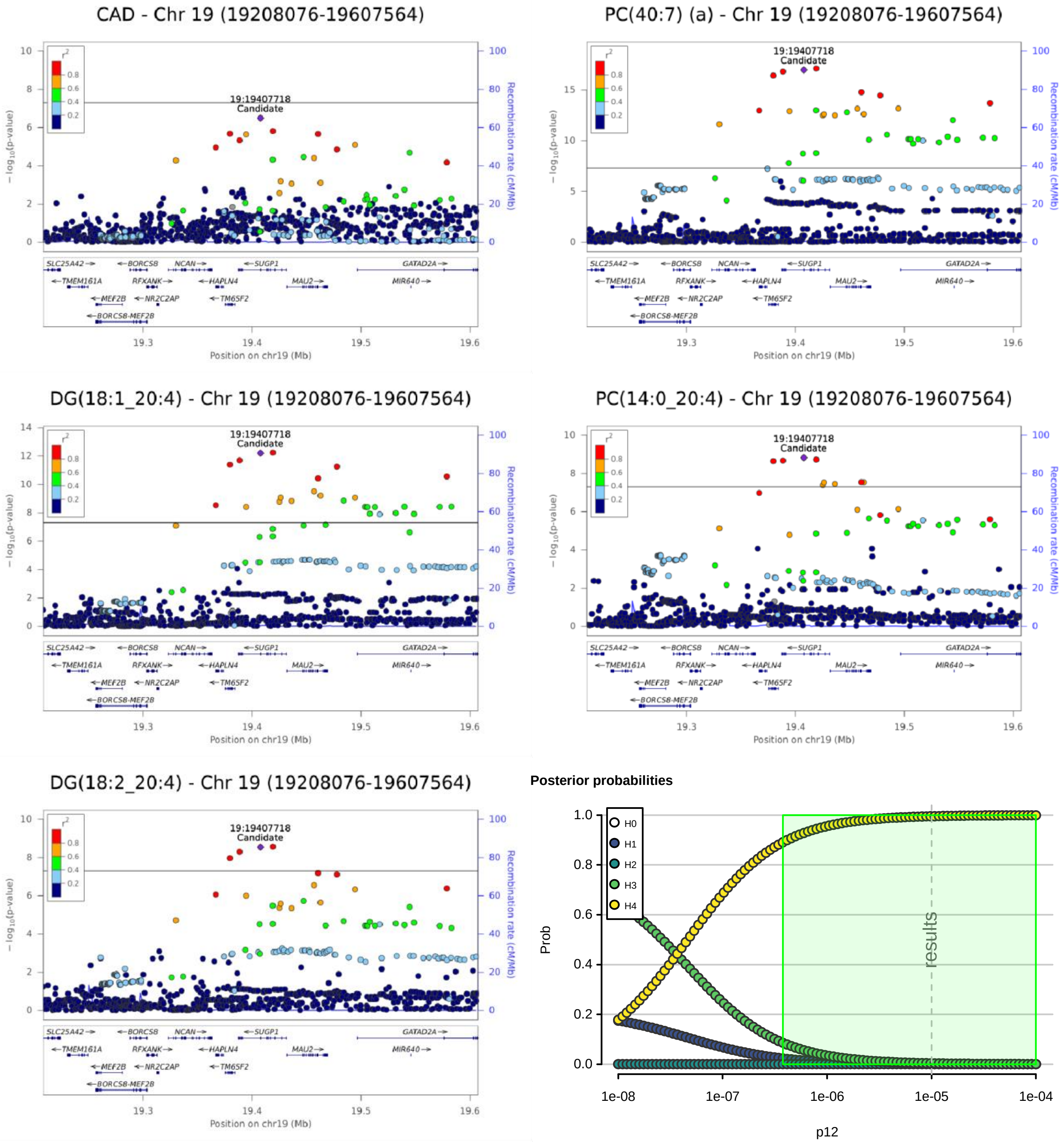

CAD - Chr 19 (19208076-19607564)
PC(40:7) (a) - Chr 19 (19208076-19607564)
DG(18:1_20:4) - Chr 19 (19208076-19607564)
PC(14:0_20:4) - Chr 19 (19208076-19607564)
DG(18:2_20:4) - Chr 19 (19208076-19607564)
19:19407718
Candidate
− log10(p-value)
Recombination rate (cM/Mb)
Position on chr19 (Mb)
SLC25A42 BORCS8 NCAN SUGP1 GATAD2A
TMEM161A RFXANK HAPLN4 MAU2 MIR640
MEF2B NR2C2AP TM6SF2
BORCS8-MEF2B
Posterior probabilities
H0
H1
H2
H3
H4
Prob
results
p12

CAD - Chr 19 (19260686-19660523)

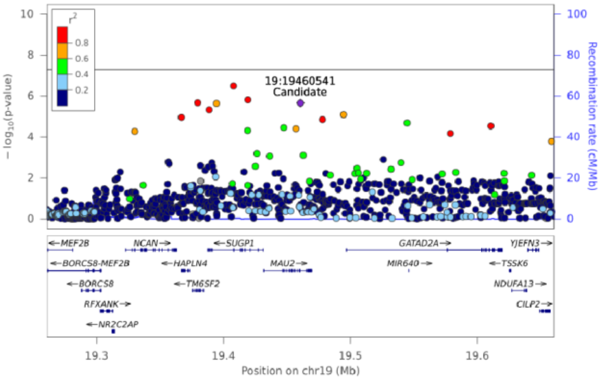


TG(50:3) [NL-16:1] - Chr 19 (19260686-19660523)

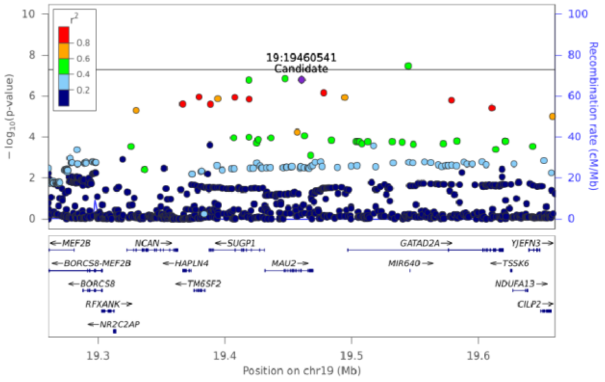


Cer(d18:1/24:0) - Chr 19 (19260686-19660523)

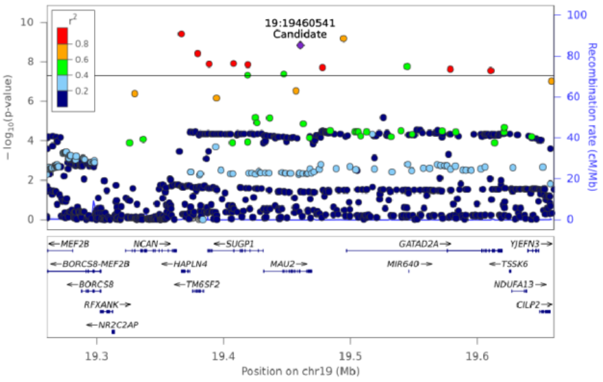


**Posterior probabilities**

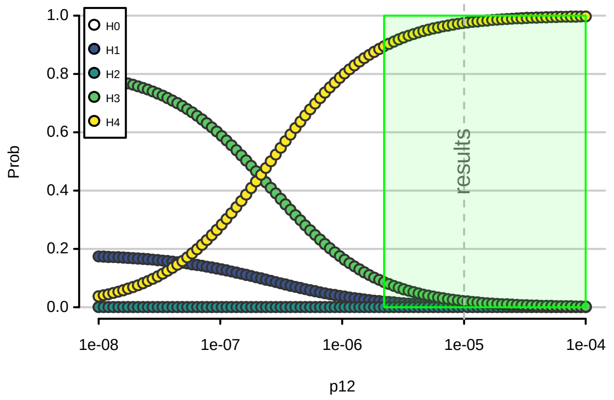


TG(50:2) [NL-18:1] - Chr 19 (19260686-19660523)

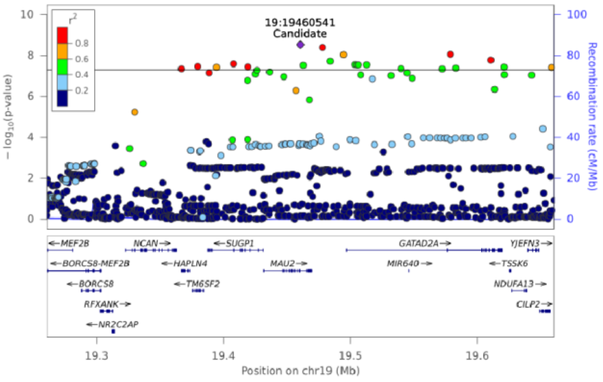

CAD - Chr 19 (19295556-19693426)

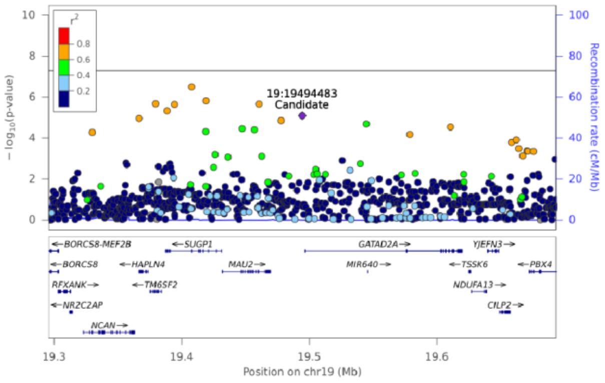


Cer(d17:1/24:1) - Chr 19 (19295556-19693426)

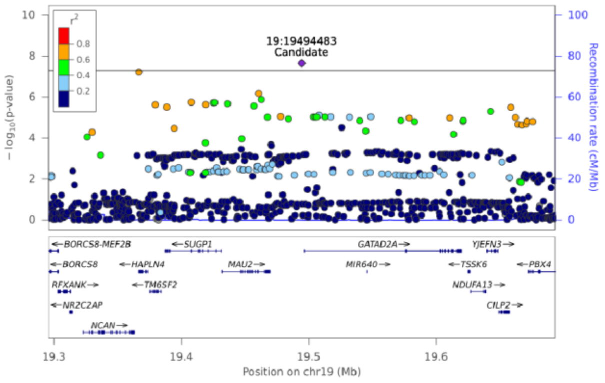


Total Cer - Chr 19 (19295556-19693426)

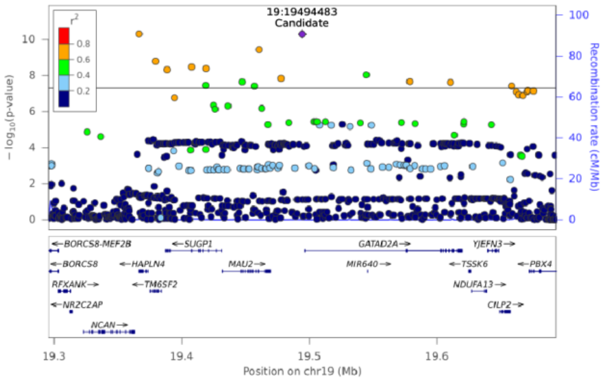


**Posterior probabilities**

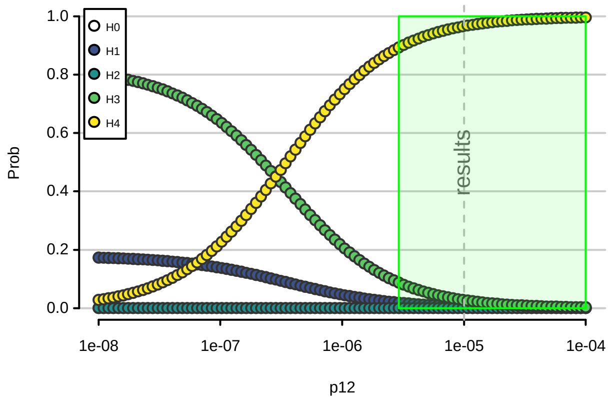


Cer(d18:1/24:1) - Chr 19 (19295556-19693426)

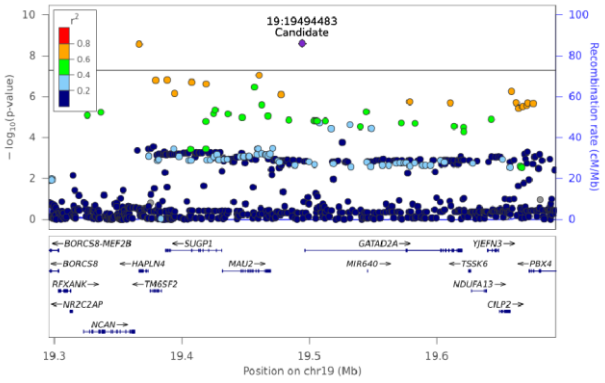

CAD - Chr 19 (45212381-45610846)

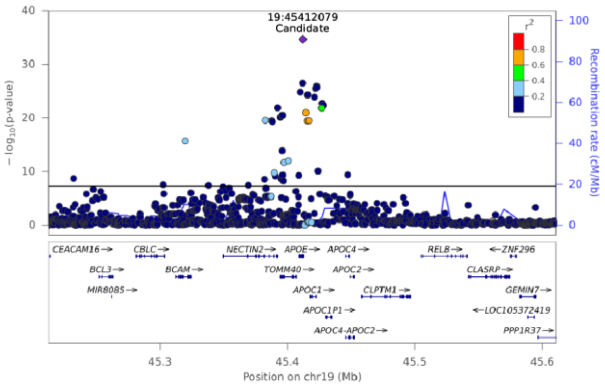


Total CE - Chr 19 (45212381-45610846)

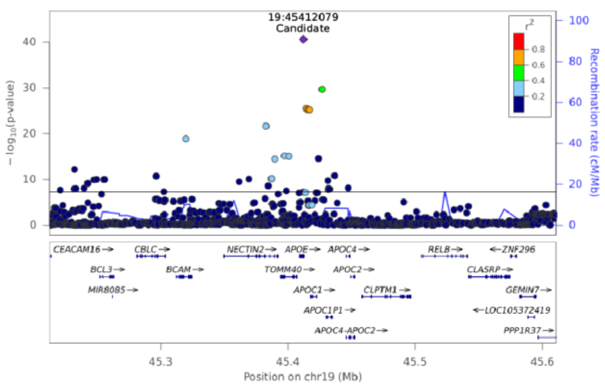


CE(16:0) - Chr 19 (45212381-45610846)

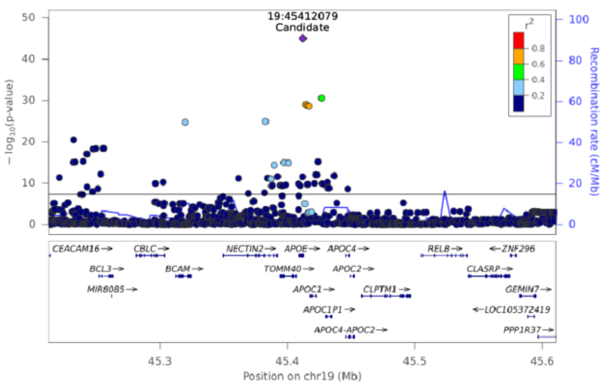


CE(18:1) - Chr 19 (45212381-45610846)

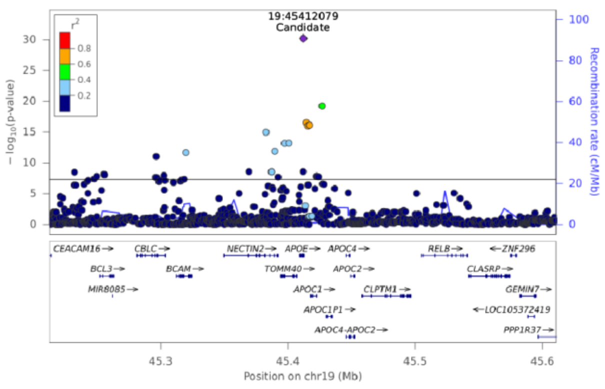


CE(18:2) - Chr 19 (45212381-45610846)

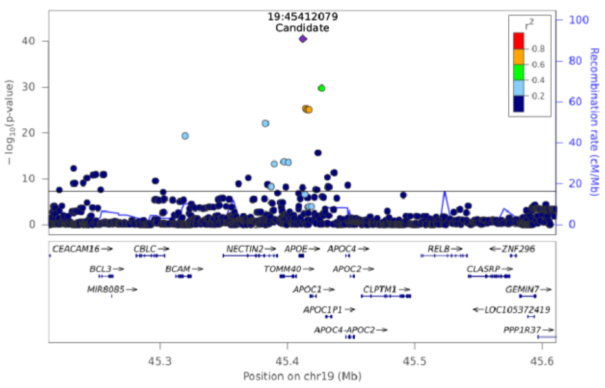


**Posterior probabilities**

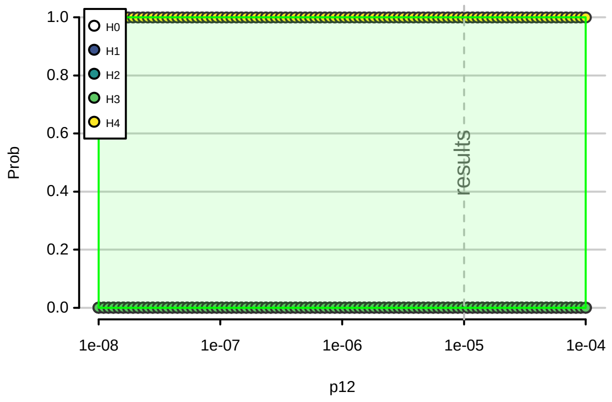

**Supplementary Note 3: Comparison of estimated lipidomic effect sizes**. To assess the impact of adjusted for clinical lipid traits or to assess the difference between adjusted and mtCOJO GWAS results, we calculated t-statistics for the difference in beta-coefficients.

$$t = \frac{\beta_{STD} - \beta_{ADJ}}{\sqrt{SE(\beta_{STD})^2 + SE\left(\beta_{ADJ}\right)^2}}$$

Where $\beta_{ADJ}$ and $\beta_{STD}$ are the estimated genetic effects from models with and without adjustment for clinical lipid traits, respectively. SE(β) is the estimated standard error of the estimates.
